# A prospective study of time-dependent childhood adversity and DNA methylation across childhood and adolescence

**DOI:** 10.1101/2021.06.28.21259423

**Authors:** Alexandre A. Lussier, Yiwen Zhu, Brooke J. Smith, Janine Cerutti, Andrew J. Simpkin, Andrew D.A.C. Smith, Matthew J. Suderman, Esther Walton, Caroline L Relton, Kerry J. Ressler, Erin C. Dunn

## Abstract

**Background:** Children exposed to adversity often have epigenetic profiles, including DNA methylation (DNAm) signatures, that differ from children without adversity histories. These signatures may be more common among children exposed during sensitive periods in development. However, it remains unclear if adversity has persistent (versus transient) effects on the epigenome across childhood and adolescence. Thus, we examined the relationship between time-varying adversity and genome-wide DNAm, measured three times from birth to adolescence using prospective data from the Avon Longitudinal Study of Parents and Children.

**Methods:** We first investigated the timing of exposure to seven types of adversity (measured 5-8 times between ages 0-11) and DNAm at age 15 using a structured life course modeling approach (SLCMA). We also assessed the persistence of adversity-DNAm associations identified from age 7 DNAm into adolescence and the influence of adversity on DNAm trajectories from ages 0-15.

**Results:** Adversity exposure was associated with differences in age 15 DNAm at 41 loci (R^2^≥0.035; p<1×10^−5^; 22 at FDR<0.05). Most loci were associated with adversities (i.e., physical, sexual, or emotional abuse; one-adult households) occurring between ages 3-5. DNAm differences present at age 7 resolved by adolescence; age 15 differences were not apparent in childhood. We also identified six distinct DNAm trajectories that highlighted both immediate and latent effects of adversity. Associations were robust in internal validation analyses using nonparametric bootstrapping.

**Conclusions:** These findings highlight the immediate and latent effects of childhood adversity on DNAm, providing a potential biological mechanism linking adversity to physical and mental health outcomes across development.

## INTRODUCTION

Children exposed to adversity, such as abuse or maltreatment^1^, family disruption or dysfunction^2^, or poverty^3^, frequently have poorer physical and mental health outcomes later in life^4-6^. Epigenetic processes, including DNA methylation (DNAm), are increasingly recognized as potential underlying mechanisms for these associations, as DNAm is responsive to life experiences^7^ and may mediate the link between environmental exposures and chronic disease^8^. Several large-scale population-based studies, systematic reviews, and meta-analyses have shown links between childhood adversity, DNAm, and adverse health outcomes across the life course^9-14^. However, prior studies investigating the epigenome of children exposed to adversity have not yet explored two key dimensions of the adversity-DNAm relationship, which are critical to understand the biological risk posed by childhood adversity, identify children at risk for poor health, and improve intervention targets for health promotion and disease prevention.

First, it remains unclear how the *timing* of childhood adversity might shape DNAm. Recent human and animal studies suggest there may be *sensitive periods* for epigenetic programming when physiological and neurobiological systems are primed for external influences, allowing life experiences to impart more enduring effects^15-18^. Few studies have investigated the specific developmental periods when adversity may have greater effects on DNAm^13,19^, with none investigating sensitive-period effects on epigenetic patterns in adolescence.

Second, little is known about how the epigenome of children exposed to adversity varies across development and how DNAm variation may shape health. In a recent article, Oh and Petronis^20^ argued that the dynamic nature of epigenetic mechanisms is best examined through longitudinal studies that model chrono-epigenetic patterns, or the temporal dynamics of epigenetic processes. Although previous studies have shown the epigenome is dynamic across development^21-30^, no study has determined how childhood adversity might influence DNAm trajectories.

To address these gaps, we examined the longitudinal relationship between early-life adversity and genome-wide DNAm across childhood and adolescence, using data collected over two decades from a subsample of youth enrolled in the Avon Longitudinal Study of Parents and Children (ALSPAC) cohort. We examined the associations between exposure to seven types of childhood adversity, assessed repeatedly between birth and age 11, and DNAm at age 15. Given the unique availability of three waves of DNAm in ALSPAC (birth, age 7, age 15), we also examined DNAm trajectories from birth to adolescence.

Our aims were to: 1) determine whether childhood adversity has time-dependent effects on adolescent DNAm; 2) characterize the developmental trajectories of DNAm linked to adversity; and 3) evaluate the persistence of previously-identified associations between adversity and DNAm in childhood^19^. To our knowledge, this study is the first to investigate the time-varying influences of childhood adversity on adolescent DNAm and DNAm trajectories from childhood to adolescence. These analyses are a novel extension of our prior work, which for one of the first times revealed sensitive periods for the effect of childhood adversity on epigenetic alterations at age 7 in ALSPAC^13,19^.

## MATERIALS AND METHODS

### Sample

ALSPAC is a large population-based birth cohort from Avon, UK of 14,451 children followed from before birth through early adulthood^31,32^. Blood-based DNAm profiles were generated for a subsample of ALSPAC mother-child pairs with cord blood at birth (n=905), whole blood at age 7 (n=970), and peripheral blood leukocytes at age 15 (n=966)^33^ (full ALSPAC details in **Supplemental materials**). Of note, ALSPAC is one of the few longitudinal studies with both repeated measures of childhood adversity and multiple waves of DNAm data.

### Measures of childhood adversity

We examined the effect of seven types of childhood adversity previously associated with DNAm^34-38^: 1) caregiver physical or emotional abuse; 2) sexual or physical abuse (by anyone); 3) maternal psychopathology; 4) one-adult households; 5) family instability; 6) financial hardship; and 7) neighborhood disadvantage. These adversities were generated from maternal reports via mailed questionnaires, collected 5-8 times between birth and age 11 (**Figure 1**; details in **Table S1**).

**Figure 1.**
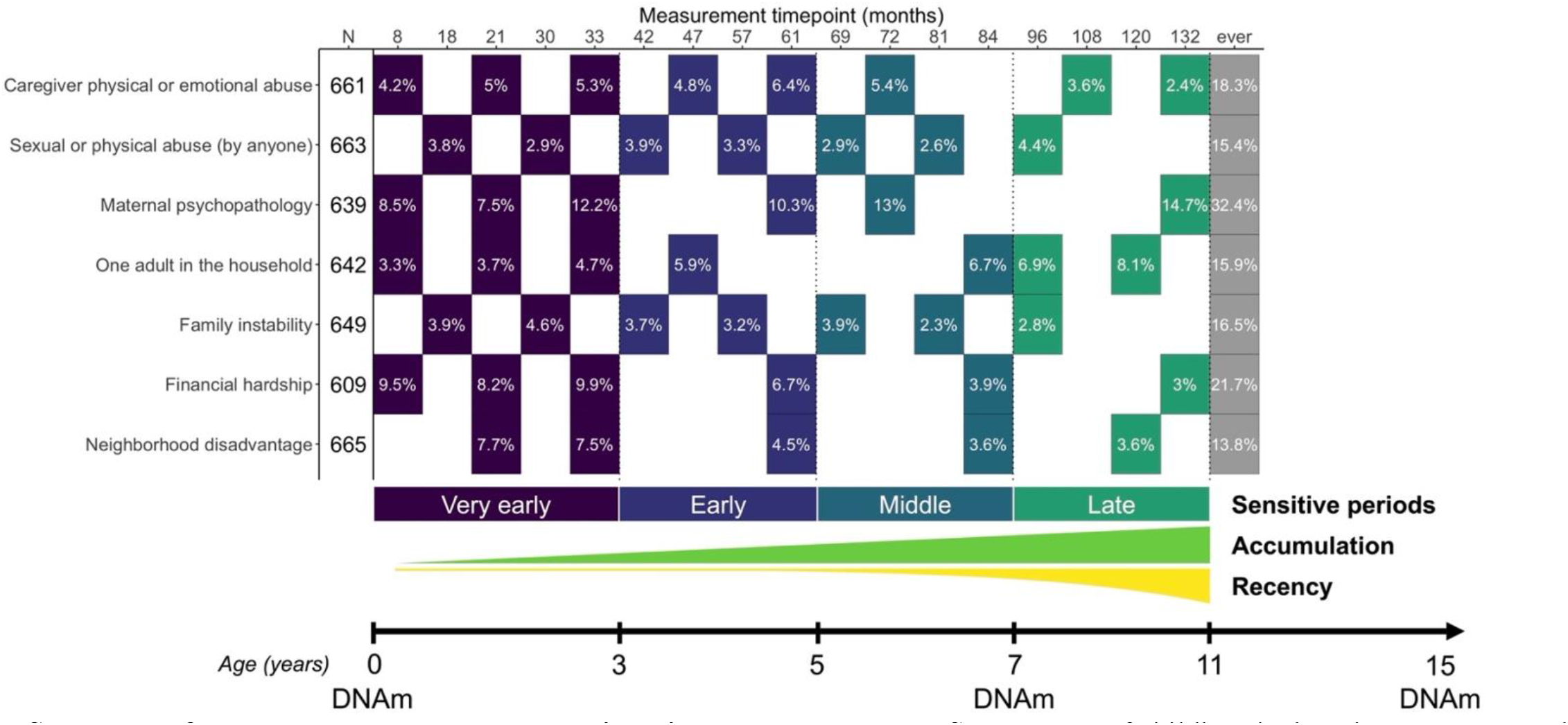
Summary of exposures and outcomes examined in the present study. Seven types of childhood adversity were assessed 5-8 times between the ages of 0 and 11. Children with complete cases across all time points and covariates were included in the present study (N=609-665). Each filled cell represents the time point when the adversity was measured, along with the prevalence of children exposed to adversity. Colors represent the four sensitive periods used to define time-dependent exposure to adversity: *very early childhood* (age 0-3), *early childhood* (age 3-5), *middle childhood* (age 5-7), and *late childhood* (age 8-11). The additional life course models tested were accumulation and recency, which reflect the total number of exposures across development and exposure to adversity weighted by time, respectively. Genome-wide DNA methylation (DNAm) data were collected at age 0, 7, and 15.

### DNAm data generation

DNAm was measured at 485,577 CpG sites using the Illumina Infinium HumanMethylation450 BeadChip microarray (Illumina, San Diego, CA). Laboratory procedures, preprocessing analyses, and quality control steps performed were described previously^33^. We removed non-variable CpGs (<5% DNAm difference between children in the 10^th^ and 90^th^ percentile), resulting in an analytic sample of 302,581 CpGs (details in **Supplemental materials**).

### Covariates

To adjust for potential confounders and be consistent with prior work^13^, we controlled for age of blood collection, sex, race/ethnicity, maternal age at birth, maternal education at birth, birthweight, number of previous pregnancies, maternal smoking during pregnancy, and cell type proportions estimated using the Houseman method^39^ (details in **Supplemental materials**).

### Analyses

#### Structured Life Course Modeling Approach (SLCMA)

Our primary analyses focused on identifying time-dependent associations between each type of childhood adversity and DNAm measured in adolescence (age 15). We used the structured life course modeling approach (SLCMA; pronounced “slick-mah”), a two-stage method that simultaneously compares *a priori* life course hypotheses explaining exposure-outcome relationships^40-44^. SLCMA first uses variable selection to identify the life course hypothesis explaining the greatest proportion of outcome variation. Effect estimates, confidence intervals, and p-values are then calculated for the selected life course hypothesis using post-selective inference. Importantly, SLCMA detects time-varying associations with more statistical power and less bias than traditional epigenome-wide association studies^13,19,45^.

We examined time-dependent associations for each adversity among children with complete data for the timepoints shown in **Figure 1**. We generated variables corresponding to six separate life course hypotheses, including four sensitive periods hypotheses encoding exposure to each childhood adversity during: 1) *very early childhood* (ages 0-2), 2) *early childhood* (ages 3-5), 3) *middle childhood* (ages 6-7), 4) *late childhood* (ages 8-11); and two additive hypotheses: 5) *accumulation of exposures* (total exposures across childhood), and 6) *recency of exposures* (total exposures weighted by age) to determine whether more recent exposures had a stronger impact than distal exposures.

We tested associations using selective inference^46^ and accounted for multiple-testing using 5% a false-discovery rate (FDR). Details on the SLCMA and functional analyses of top loci are in **Supplemental materials**.

To our knowledge, there are currently no comparable cohort studies with repeated measures of childhood adversity and DNAm in which to replicate our findings. Thus, we assessed the robustness of the SLCMA results through internal validation analyses of our associations using ordinary nonparametric bootstrapping (**Supplemental materials**). We also assessed the effects of potential confounders or alternate mediators of the association between childhood adversity and DNAm at age 15, including exposures to other types of childhood adversity in the same or different sensitive periods (**Supplemental materials**).

#### Trajectories of DNAm response to childhood adversity

The three waves of longitudinal DNAm data available in ALSPAC allowed us to investigate patterns of DNAm across development. Thus, we pursued three additional sets of analyses, building from the SLCMA findings.

##### Pre-existence of age 15 associations

We first determined whether DNAm differences identified at age 15 using the SCLMA emerged earlier in development. Using linear regression, we tested whether exposure to adversity was associated with DNAm at the same top loci at birth or age 7, adjusting for covariates.

##### Types of DNAm trajectories across development

We also investigated DNAm patterns beyond the cross-sectional age 15 time point, studying longitudinal change and stability patterns in our top loci among children from three distinct exposure groups: 1) children who had adversity exposure *during* the sensitive period identified from the SLCMA (labeled as exposed-SP); 2) children who had adversity exposure *outside* the sensitive period identified from the SLCMA (exposed-other); and 3) children who were never exposed to adversity. We performed an analysis of variance (ANOVA) of the statistical interaction between age at DNAm collection and exposure group, controlling for DNAm repeated measures as fixed effects. Loci showing differences between exposure groups across time (i.e., group-by-age interactions; FDR<0.05) were carried forward to subsequent analyses, to identify more granular temporal patterns between exposure groups.

From these loci, we identified different types of DNAm trajectories based on ANOVA and Tukey post-doc analyses of three main parameters: 1) mean exposure group differences *across ages*, 2) mean age differences *across exposure groups*, and 3) exposure group differences *within* each age. We then used hierarchical clustering of these three parameters to identify homogeneous subsets within these longitudinal patterns (**Supplemental Materials**).

##### Persistence of childhood DNAm differences to adolescence

We assessed whether DNAm alterations linked to childhood adversity, which we previously identified at age 7^19^, persisted to adolescence. Specifically, we performed linear models between adversity and age 15 DNAm data for these 46 childhood loci (**Supplemental materials**).

## RESULTS

### Sample characteristics and prevalence of exposure to adversity

Demographic characteristics did not differ between the ARIES sample and children exposed to any adversity between ages 0-11 (**Table S2**). The prevalence of exposure to a given adversity from age 0-11 ranged from 15.1% (sexual/physical abuse) to 34.8% (maternal psychopathology) (**Figure S1; Table S3**). The correlation of exposure within each adversity across development ranged from 0.36 (family instability) to 0.786 (one-adult households). Different types of adversity were weakly correlated (r_avg_=-0.04-0.16).

### Childhood adversity showed time-dependent associations with adolescent DNAm profiles

Across all types of childhood adversity, 41 loci showed significant associations between exposure to adversity and DNAm levels at age 15 (≥3.5% of DNAm variance explained by adversity; nominal p<1×10^−5^; **Table 1; Table S4**). Of these, 22 loci were significant after multiple-test correction (FDR<0.05). As we and others have recently shown that p-values are poor metrics of statistical inference when used on their own^47,48^, particularly in the context of time-varying effects^19^, we focus on downstream analyses on the subset of CpGs that met both the R^2^ threshold and nominal p-value.

**Table 1.**
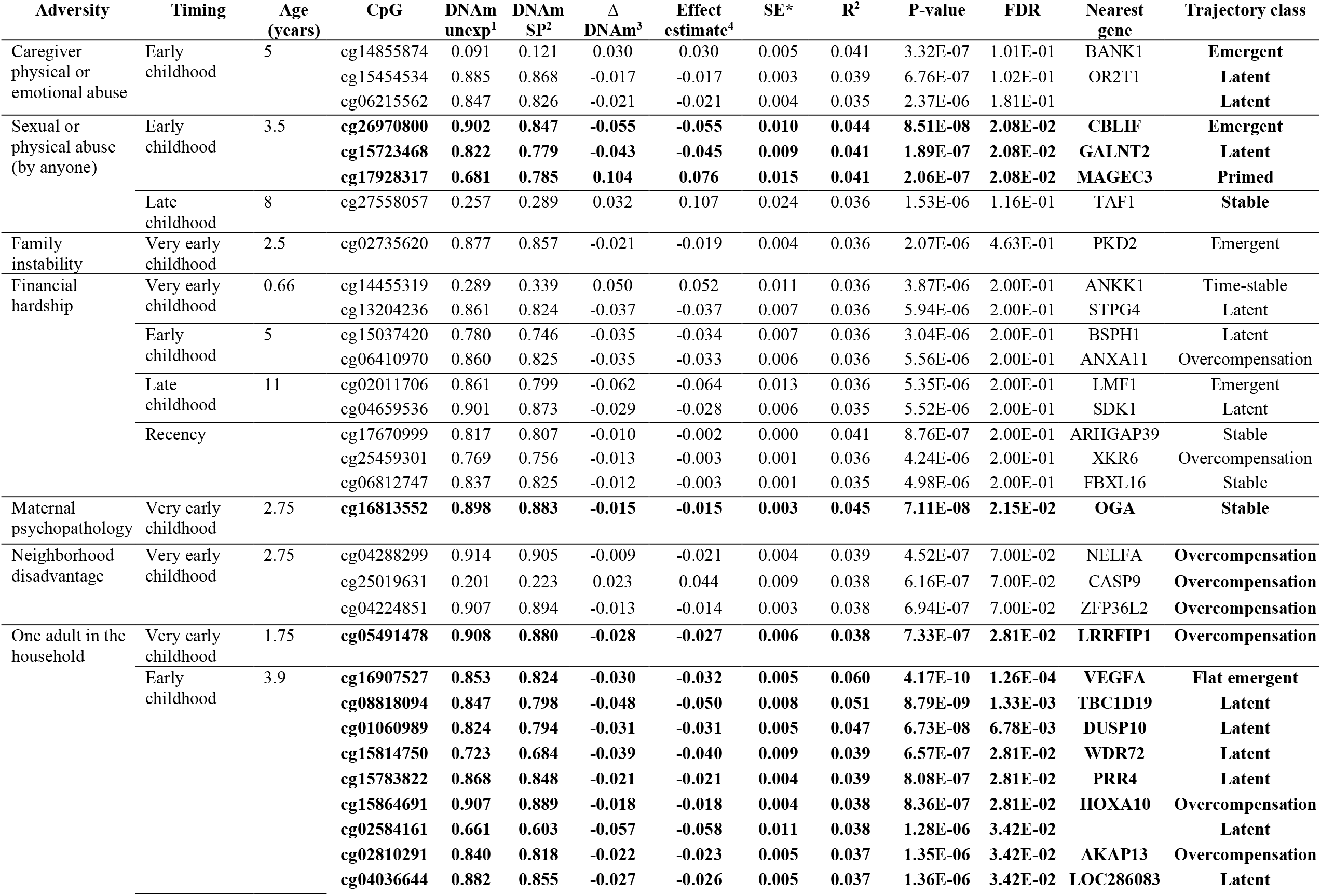
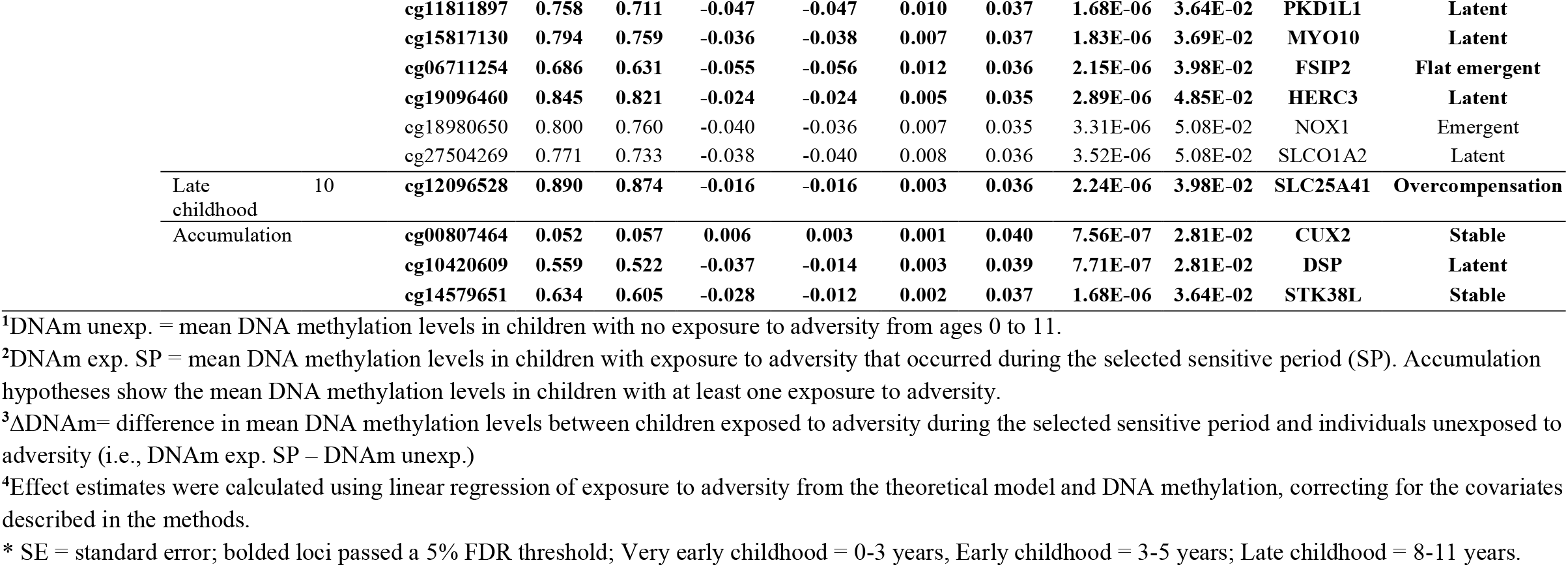
Top associations between time-dependent exposure to adversity and DNA methylation at age 15.

Sensitive periods were the most often selected life course hypothesis by the SLCMA, with 35 loci showing associations with childhood adversity that occurred during *very early childhood* (20% or 8 of 41), *early childhood* (56% or 23 of 41), or *late childhood* (10% or 4 of 41) (**Figure 2**). Only 3 loci (7%) showed associations with either the accumulation or recency of adversity.

**Figure 2.**
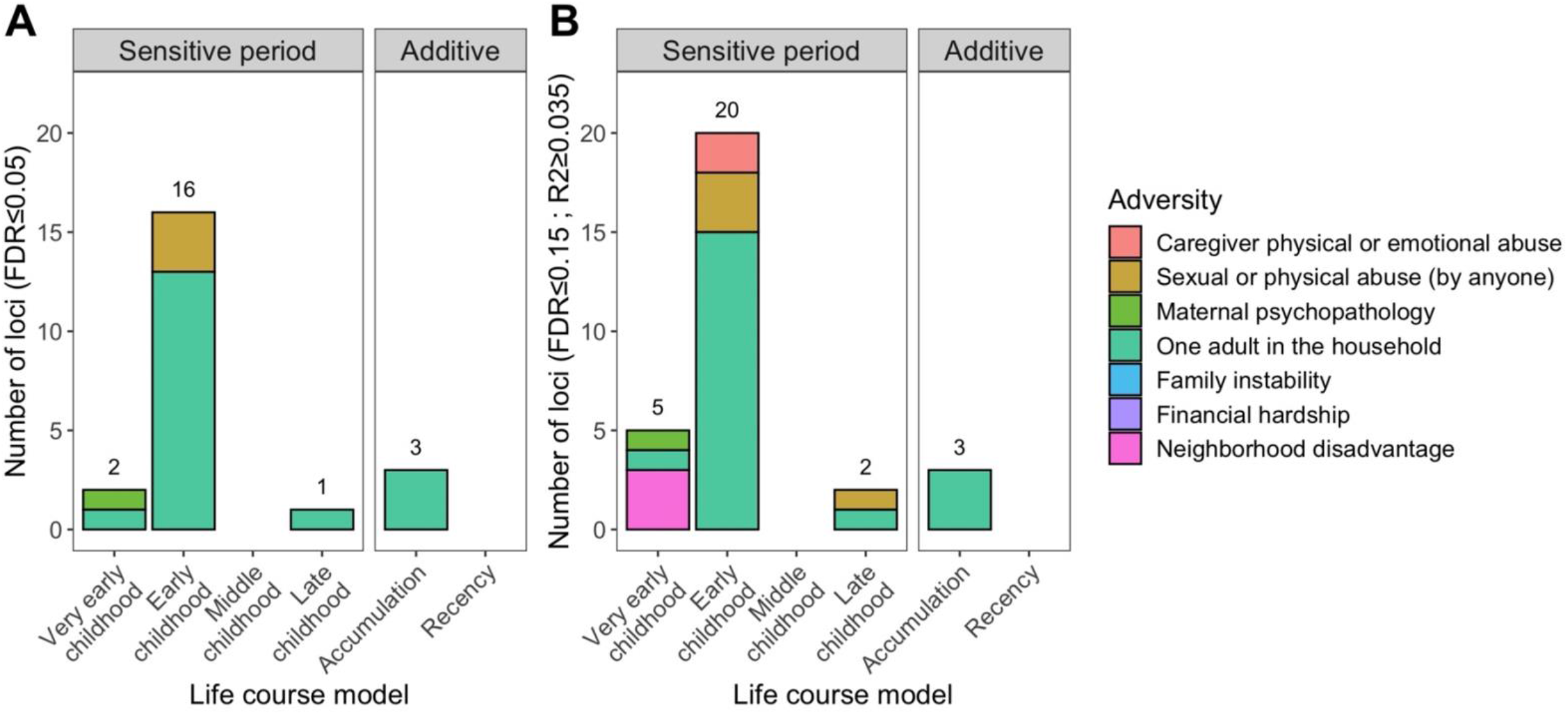
Life course theoretical models selected by the SLCMA for top loci at age 15. The life course theoretical models were split by sensitive periods (i.e., exposure to adversity during specific childhood periods) or additive models (i.e., accumulation or recency of exposures). Colors represent the different types of adversity. The distribution of theoretical models for top loci was significantly different than random chance, with exposure to adversity during sensitive periods more frequently predicting DNA methylation levels as compared to the additive models. **A)** 22 loci were identified at a false-discovery rate (FDR) <0.05. Most loci were associated with exposure to one adult households during early childhood. **B)** 41 loci were identified at an R2≥0.035 cutoff and p<1×10^−5^ threshold, which again mainly showed associations with adversity occurring during early childhood.

Most of these associations were for exposure to one-adult households (20 of 41 loci). We also identified associations with caregiver physical or emotional abuse (3 loci), sexual or physical abuse by anyone (4 loci), family instability (1 locus), financial hardship (9 loci), maternal psychopathology (1 locus), and neighborhood disadvantage (3 loci).

Childhood adversity was mainly associated with a decrease in DNAm (20 of 23 loci). On average, childhood adversity exposure was linked to a 3.5% absolute difference in DNAm (range 0.9-10.4%). For loci associated with accumulated time living in one-adult households, each additional timepoint of exposure was associated with a 1% difference in DNAm (range 0.3-1.4%). For loci associated with the recency of financial hardship, one additional exposure was linked to a −1.3% to 2.3% change in DNAm per year of age at exposure.

Top loci showed higher representation in regions of lower CpG density, such as enhancers (χ^2^=7.1, p=0.008) and Open Seas (χ^2^=13.6, p=0.018) (**Figure S2**). Most loci (28/41) also had weak, but positive correlations between brain and blood^49^ (**Table S5**; **Figure S3**), suggesting adversity-induced alterations to blood DNAm may reflect similar changes in the central nervous system. Although no biological processes were significantly enriched in top loci using the DAVID gene ontology tool^50,51^ (**Figure S4**), seven genes linked to sexual/physical abuse (*TAF1*), family instability (*PKD2*), financial hardship (*FBXL16, XKR6*), or one-adult households (*DSP, CUX2, STK38L*) showed evidence of strong evolutionary constraint (**Table S4; Figure S5**)^52^. Together, these findings suggest different types of childhood adversity may act through diverse biological processes and hunt at a potential evolutionary role for genes responsive to parental and social environments (**Supplemental materials**).

### Associations between childhood adversity and DNAm at age 15 were internally validated and specific to each adversity

Internal validation analysis of the top associations yielded nearly identical results to the initial analyses (largest difference in effect estimates=2.03%); all bootstrap effect estimates were significant at the 5% level (**Figure S6; Table S6**). Our results also remained stable when correcting for exposure to other adversities during the sensitive period or across childhood, suggesting they were not influenced by co-occurring incidences of adversity (**Supplemental materials; Figure S7-9**). Together, these results point to the robustness and specificity of associations between time-varying childhood adversity and DNAm at age 15.

### DNAm differences at age 15 were not present earlier in childhood

For the 41 loci identified in age 15 DNAm, none showed associations between adversity and DNAm at birth (**Table S7**) or age 7 (**Table S8**) (FDR<0.05). Notably, the age 7 effect estimates were *smaller* than the age 15 associations, with consistent directions-of-effect in about half (20 of 41) (**Figure 3A**). Importantly, the emergence these associations was not explained by early-life confounders or biological mediators during adolescence (**Supplemental materials; Figures S10-16**), suggesting some adolescent differences may emerge later in development and become stronger with time.

**Figure 3.**
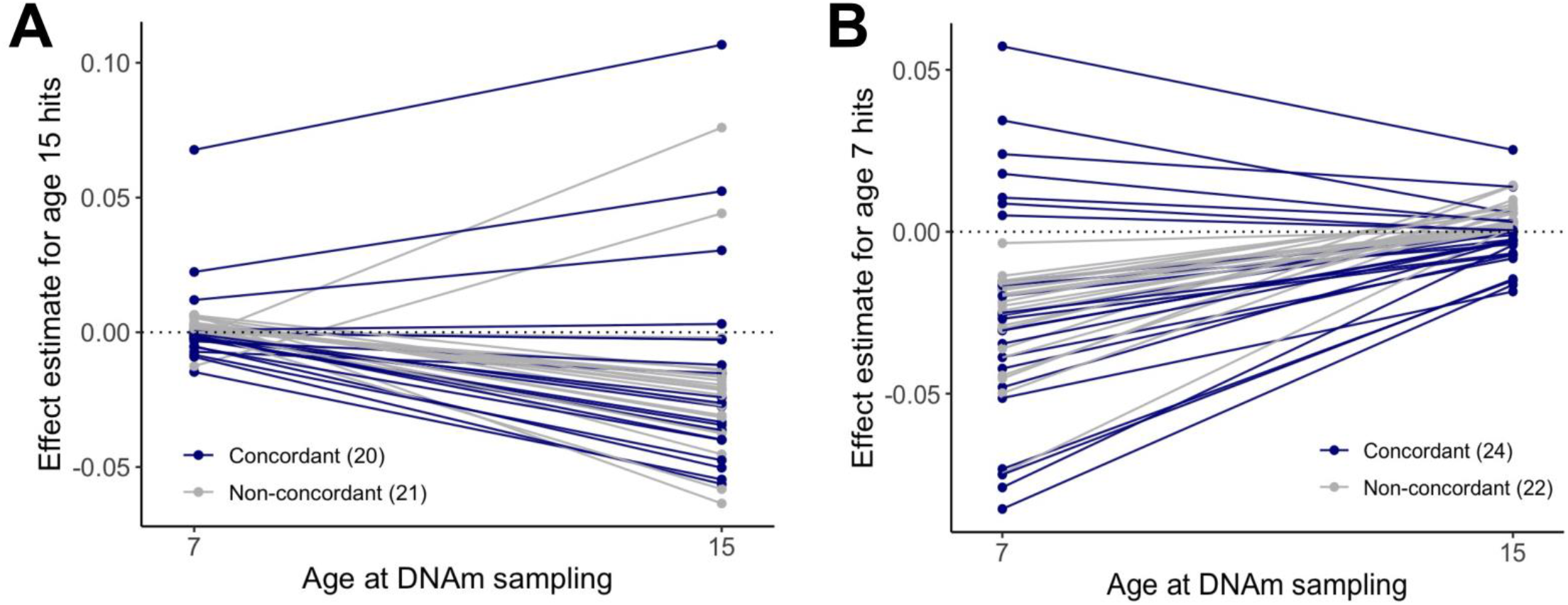
Persistence and stability of associations between childhood adversity and DNA methylation across development. **A)** The effect estimates of associations between childhood adversity and DNAm at age 7 or age 15 generally showed variable directions of effect for the significant loci identified from the SLCMA at age 15 (20 concordant and 21 non-concordant directionality). Effect estimates for age 7 DNAm data were also smaller than those at age 15, suggesting that these loci showed latent responses to adversity. **B)** The effect estimates of associations between childhood adversity and DNAm at age 7 or age 15 generally showed variable directions of effect for the significant loci identified in a previous study of age 7 DNAm (24 concordant and 22 non-concordant directionality). Effect estimates for age 15 DNAm data were also smaller than those at age 7, suggesting that these loci showed early responses to adversity that resolved by adolescence

### Childhood adversity was linked to distinct trajectories of DNAm across development

Moving beyond adolescent DNAm alone, 34 of the 41 loci had significant adversity exposure group-by-age interactions (FDR<0.05), suggestive of more complex patterns of change and stability across development. From these loci, we identified five additional types of longitudinal DNAm trajectories (**Table S9**), which showed distinct DNAm patterns across ages and adversity exposure groups (**Figures S17-20**), but not between the FDR and R^2^ subsets of CpGs (**Figure S21**). **Table 2** provides a full description and examples of the patterns distinguishing DNAm trajectories, which included differences that emerged earlier versus later in development, differences between children exposed during a sensitive period or at other developmental stages, and differences linked to age at DNAm measurement.

**Table 2.**
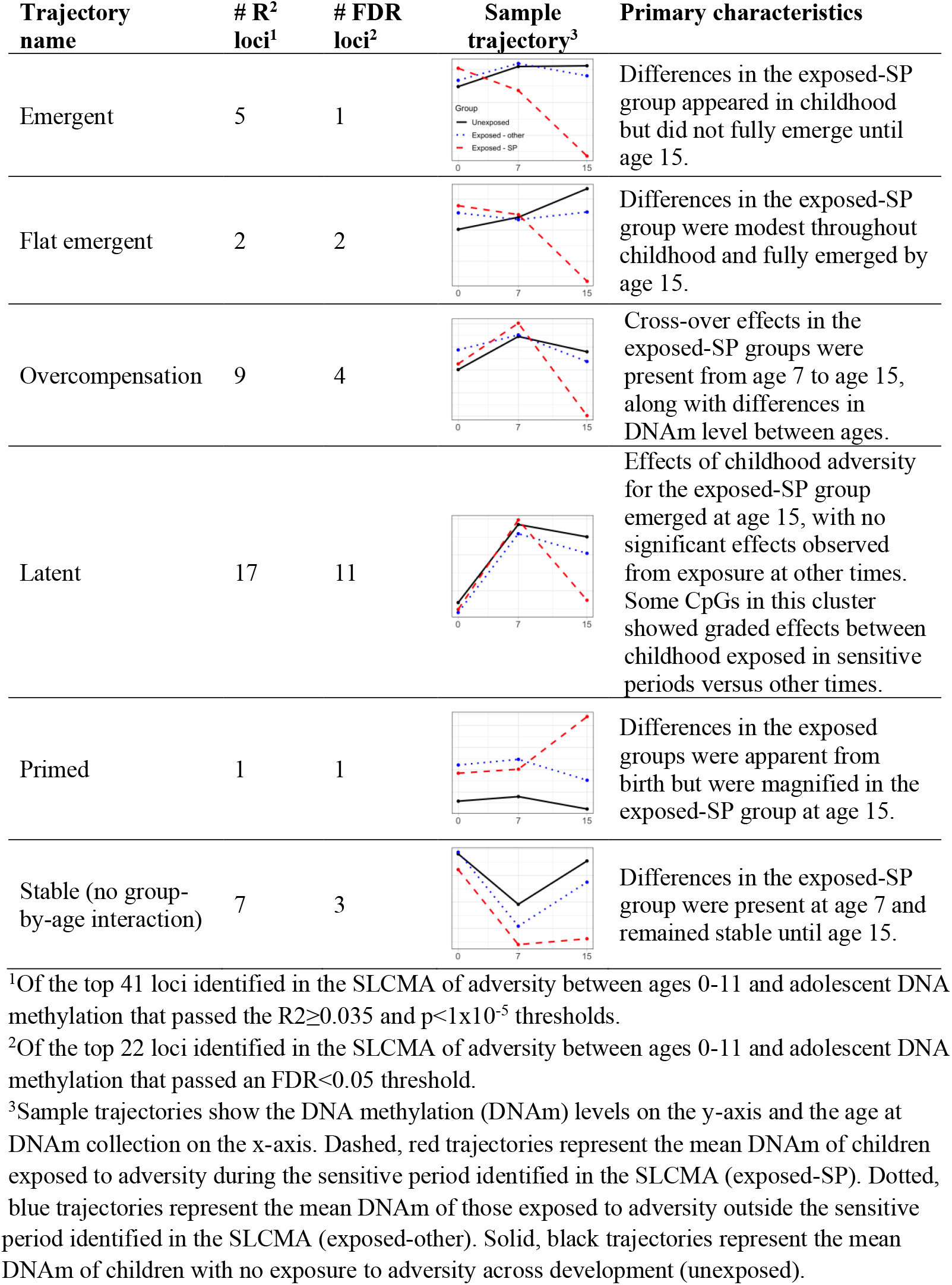
Types of DNAm trajectories and response to childhood adversity.

| Trajectory name | # R<sup>2</sup> loci<sup>1</sup> | # FDR loci<sup>2</sup> | Sample trajectory<sup>3</sup> | Primary characteristics |
| --- | --- | --- | --- | --- |
| Emergent | 5 | 1 |  | Differences in the exposed-SP group appeared in childhood but did not fully emerge until age 15. |
| Flat emergent | 2 | 2 |  | Differences in the exposed-SP group were modest throughout childhood and fully emerged by age 15. |
| Overcompensation | 9 | 4 |  | Cross-over effects in the exposed-SP groups were present from age 7 to age 15, along with differences in DNAm level between ages. |
| Latent | 17 | 11 |  | Effects of childhood adversity for the exposed-SP group emerged at age 15, with no significant effects observed from exposure at other times. Some CpGs in this cluster showed graded effects between childhood exposed in sensitive periods versus other times. |
| Primed | 1 | 1 |  | Differences in the exposed groups were apparent from birth but were magnified in the exposed-SP group at age 15. |
| Stable (no group-by-age interaction) | 7 | 3 |  | Differences in the exposed-SP group were present at age 7 and remained stable until age 15. |

<sup>1</sup>Of the top 41 loci identified in the SLCMA of adversity between ages 0-11 and adolescent DNA methylation that passed the $R^2 \geq 0.035$ and $p < 1 \times 10^{-5}$ thresholds.
<sup>2</sup>Of the top 22 loci identified in the SLCMA of adversity between ages 0-11 and adolescent DNA methylation that passed an $FDR < 0.05$ threshold.
<sup>3</sup>Sample trajectories show the DNA methylation (DNAm) levels on the y-axis and the age at DNAm collection on the x-axis. Dashed, red trajectories represent the mean DNAm of children exposed to adversity during the sensitive period identified in the SLCMA (exposed-SP). Dotted, blue trajectories represent the mean DNAm of those exposed to adversity outside the sensitive
period identified in the SLCMA (exposed-other). Solid, black trajectories represent the mean DNAm of children with no exposure to adversity across development (unexposed).

### Associations between adversity and childhood DNAm did not persist into adolescence

Of the 46 CpG sites previously showing time-varying associations between adversity and DNAm at age 7 ^19^, only one showed an association at age 15 (p<0.05; **Table S10**), which did not pass multiple-test correction. Again, approximately half of loci showed consistent direction-of-effect between age 7 and 15 (24/46) (**Figure 3B**). These findings suggest some childhood epigenetic responses to adversity may not persist into adolescence.

## DISCUSSION

The main finding from this study is that associations between childhood adversity and DNAm vary across the life course, manifesting at different developmental stages through distinct patterns of persistence and latency. To our knowledge, this is the first study to incorporate time-dependent measures of childhood adversity in the study of human longitudinal epigenetic patterns.

Our findings point to early childhood – meaning the period between ages 3 to 5 – as a possible sensitive period for the biological embedding of childhood adversity that manifests in adolescence. These findings are consistent with multiple prior human^11,13^ and animal studies^53,54^ showing that exposures earlier in life may have greater influence on epigenetic patterns^19^. As early childhood is a time of rapid cognitive, social, emotional, and regulatory development^55^, epigenetic processes may be more malleable during this period^24^, resulting in increased sensitivity to life experiences that shape DNAm levels and trajectories across development. Importantly, these findings suggest early childhood as a potential period for focused interventions to limit or prevent the long-term sequelae of childhood adversity.

Of the seven types of adversity examined, exposure to single parent households had the greatest number of associations to DNAm in adolescence. By contrast, previous research on DNAm from the same children at age 7 identified no associations with one-adult households^19^, suggesting these effects may be adolescent-specific. Prior studies have shown the effects of single parent households begin to emerge around puberty, manifesting through shifts in the timing of puberty^56,57^, poorer self-esteem^58^, and higher depressive symptoms^59^ and externalizing behaviors^59^. These findings suggest a latency to the effects of one adult households, which may not become apparent until the rapid biological changes that occur during puberty^60^.

Curiously, we observed fewer associations than expected for other adversities, such as maternal psychopathology and experiences of sexual, physical, or emotional abuse. These adversities may have subtler influences on the adolescent epigenome, requiring larger sample sizes or meta-analyses to uncover. Notably, none of our top loci overlapped between different types of childhood adversity, nor were they present among top loci from a twin study of adolescent victimization (N=118), where half of the children in the sample were exposed to severe victimization^22^. However, it remains unknown whether subtypes of adversity might have differential effects on downstream biological processes and vulnerability to disease. As has been discussed in ongoing debates surrounding the “lumping or splitting” of childhood adversities in clinical research^61^, it may be that different dimensions of adversity result in different epigenetic signatures, a hypothesis supported by the finding that adjusting for other types of adversity had modest influences on the strength of associations. To this end, we investigated potential differences in sensitive periods for exposures to adversities using a threat vs deprivation paradigm^62^, showing that exposures to deprivation-type adversities may have more influence on adolescent DNAm than threat-type adversities, particularly when they occur in early childhood (**Figure S22**).

Arguably the most novel set of findings from our study concerned the patterns of stability and change in the relationship between adversity and DNAm. Most DNAm trajectories showed primarily *latent* effects of adversity, meaning they did not emerge until age 15 in youth exposed to adversity. These findings align with previous longitudinal studies of genome-wide DNAm from ALSPAC and Project Viva, which have shown that early-life stressors, such as prenatal maternal smoking^26^ and socio-economic disadvantage during childhood^21,27^, can have both immediate and latent effects on DNAm during childhood and adolescence. Subtle desynchronization of DNAm levels may appear earlier in development, while evading immediate detection until later in life. These “sleeper” effects may explain why complex diseases unfold over years of development, rather than immediately after exposures or risk factors^20^. Future research should investigate whether these latent effects of childhood adversity on the epigenome persist into adulthood and whether they are more likely to influence physical and mental health than alterations arising earlier in development.

Similarly, the DNAm differences we previously observed at age 7 did not persist into adolescence^19^. Studies on early-life stressors^21,27^ and markers of prenatal environments, such as birthweight and gestational age^29^, or maternal weight before and during pregnancy^28^, parallel these findings, showing that DNAm differences linked to early-life environments do not generally persist across time. Whether these patterns resolved naturally or due to active intervention is unknown and should be investigated to determine whether interventions are needed to rescue the effects of early-life stressors. Nevertheless, even short-term alterations that eventually fade over time could still alter the developmental trajectories of cellular pathways or physiological systems, influencing downstream health and risk for disease^63^.

Although this study did not focus on causal relationships, several differentially methylated genes were implicated in key processes that could influence downstream disease. For instance, *CUX2* is transcription factor involved in dendrite and synapse formation^64^, alterations to which could influence neurodevelopment and vulnerability to mental disorders. Several top genes, including *DUSP10*^65^, *DSP*^66^, and *VEGFA*^67^, are also linked to cardiac function, and may partially reflect mechanisms linking childhood adversity to heart disease^68^. However, additional research is needed to identify the true health consequences of these differences and determine whether short- and/or long-term DNAm changes mediate the link between childhood adversity and health outcomes.

Our study had limitations. First, DNAm data were generated from slightly different tissue types at each wave. Although we corrected for cell type composition using established methods, differences in the stability of DNAm differences between waves may have been partially driven by tissue-based differences and variability. Second, we were unable to replicate our findings in an independent sample, due to the lack of data available from other cohort studies. We also cannot fully rule-out the possibility that our findings were influenced by unmeasured or technical factors specifically influencing age 15 associations. However, our results were robust in internal validation analyses and when controlling for 14 potential confounders. Nonetheless, future studies are needed to fully replicate and confirm these longitudinal epigenetic responses to childhood adversity. Third, our analytic subsample was mainly composed of children from European descent. This lack of diversity limited the generalizability of our findings. Given the disparities in childhood adversity exposure and health^69^, our findings should be replicated in more diverse cohorts.

## CONCLUSIONS

This study highlights developmental variability in the relationship between childhood adversity and DNAm trajectories and its potential role in adversity-related health outcomes. Future studies should continue to investigate longitudinal measures of DNAm to identify the potential role of latent and persistent epigenetic alterations in driving short- and long-term health outcomes. Ultimately, this research will help guide intervention strategies and identify individual who are at higher risk for physical and mental disorders arising from exposure to childhood adversity.

## Data Availability

All data are available by request from the ALSPAC Executive Committee for researchers who meet the criteria for access to confidential data (
http://www.bristol.ac.uk/alspac/researchers/access/).

## ACKNOWLEDGEMENTS

This work was supported by the National Institute of Mental Health of the National Institutes of Health (grant number R01MH113930 awarded to ECD). The content is solely the responsibility of the authors and does not necessarily represent the official views of the National Institutes of Health. Dr. Dunn and Dr. Lussier were also supported by a grant from One Mind. We are extremely grateful to all the families who took part in the ALSPAC study, the midwives for their help in recruiting them, and the whole ALSPAC team, which includes interviewers, computer and laboratory technicians, clerical workers, research scientists, volunteers, managers, receptionists, and nurses. The UK Medical Research Council and Wellcome (Grant ref: 217065/Z/19/Z) and the University of Bristol provide core support for ALSPAC. A comprehensive list of grants funding is available on the ALSPAC website (http://www.bristol.ac.uk/alspac/external/documents/grant-acknowledgements.pdf); This research was specifically funded by grants from the BBSRC (BBI025751/1; BB/I025263/1), MRC IEU (MC_UU_00011/5), National Institute of Child and Human Development (R01HD068437), NIH (5RO1AI121226-02), and CONTAMED EU (212502). This publication is the work of the authors, whom will serve as guarantors for the contents of this paper.

Dr. Walton is funded by CLOSER, whose mission is to maximize the use, value, and impact of longitudinal studies (www.closer.ac.uk). CLOSER was funded by the Economic and Social Research Council (ESRC) and the Medical Research Council (MRC) between 2012 and 2017. Its initial five-year grant has since been extended to March 2021 by the ESRC (grant reference: ES/K000357/1). The funders took no role in the design, execution, analysis, or interpretation of the data or in the writing up of the findings. Dr. Walton is also supported by the European Union’s Horizon 2020 research and innovation programme (grant nº 848158). Finally, we would also like to thank Dr. Garrett Fitzmaurice for his guidance in the characterization of DNAm trajectories across development. We also thank Elizabeth Leblanc for her assistance with the prediction of age at pubertal onset using superimposition by translation and rotation (SITAR).

## AUTHOR CONTRIBUTIONS

AAL designed the study, performed all analyses, interpreted the results, and wrote the manuscript. YZ, BJS, JC, AJS, ADACS, MJS, EW, CLR, and KJR assisted in the design and interpretation of the study and provided critical input in writing the manuscript. ECD obtained grant support for this work, designed the study, interpreted the results, and helped write the manuscript.

