## Supplemental materials for "A prospective study of time-dependent childhood adversity and DNA methylation across childhood and adolescence"

**Figure S22. Enrichment of top adolescent loci within the threat versus deprivation paradigm. ..70**

### SUPPLEMENTAL METHODS

#### Cohort description

Data came from the Avon Longitudinal Study of Parents and Children (ALSPAC), a longitudinal birth cohort of children born to mothers who were living in the county of Avon, England, with expected delivery dates between April 1991 and December 1992[1, 2]. The main goal of the ALSPAC study is to increase knowledge of the pathways influencing lifelong health, with a focus on the genetic and environmental determinants of health and disease. A total of 14,451 pregnant women participated in the study and of 14,062 of eligible live births who were alive at one year of age (n=13,988 children) were enrolled in the study. Please note that the study website contains details of all the data that is available through a fully searchable data dictionary and variable search tool: http://www.bristol.ac.uk/alspac/researchers/our-data/.

Ethical approval for the study was obtained from the ALSPAC Ethics and Law Committee and the Local Research Ethics Committees. Consent for biological samples has been collected in accordance with the Human Tissue Act (2004). Informed consent for the use of data collected via questionnaires and clinics was obtained from participants following the recommendations of the ALSPAC Ethics and Law Committee at the time. All data are available by request from the ALSPAC Executive Committee for researchers who meet the criteria for access to confidential data (http://www.bristol.ac.uk/alspac/researchers/access/). Secondary analyses of ALSPAC data were approved with oversight by the Mass General Brigham Institutional Review Boards (IRB) (Protocol 2017P001110).

**DNA methylation data**

The analytic sample came from a subsample of ALSPAC, the Accessible Resource for Integrated Epigenomics Studies (ARIES). The subsample consisted of 1,018 mother-child pairs from whom blood-based DNA methylation data were collected. Participants in the ARIES subsample were randomly selected from ALSPAC participants with complete data across at least five timepoints of data collection [3]. Three timepoints of DNAm were collected, including cord blood at birth (n=905), whole blood at age 7 (n=970), and peripheral blood mononuclear cells at age 15 (n=966). 846 individuals had DNAm collected at all three timepoints. Number of samples are based on the number of samples with available data after the pre-processing procedures described in the main text.

**DNA methylation pre-processing and normalization**

DNAm data were processed using the *meffil* package in R, which performs background correction and functional normalization of DNAm data [4]. Twins and samples with >10% of CpG sites with a detection p-value >0.01 or a bead count <3 were removed, as were cross-hybridizing probes and polymorphic probes. To remove possible outliers, we winsorized the beta values (i.e., values that represent the percent of methylation at each CpG site), setting the bottom 5% and top 5% of values to the 5th and 95th quantile, respectively [5]. Finally, we removed probes showing little variability across individuals, defined as CpGs with <5% difference in DNAm between the $10^{th}$ and $90^{th}$ percentile of values. The final analytic sample after pre-processing consisted of 966 youths and 302,581 CpGs with DNAm data measured at age 15. DNAm measured at age 0 and 7 were similarly pre-processed and normalized.

**Coding of covariates**

Across all analyses, we controlled for the following covariates, which were measured at birth and coded as follows:

1. *Sex* – coded as a binary variable, as reported at birth and confirmed from epigenetic data.
2. *Race/ethnicity* – coded as a binary variable corresponding to white or non-white, as our analytic sample was predominantly white and previous work in the ARIES subsample found no strong evidence of population stratification [6].
3. *Maternal age at birth* – coded as a categorical variable with three categories of response, ages 15-19, ages 20-35, and age 36+.
4. *Number of previous pregnancies* – coded as a categorical variable, with response categories of 1, 2, and 3+.
5. *Maternal smoking during pregnancy* – coded as an exposure if the mother smoked during at least two trimesters of pregnancy, as previously described [7].
6. *Child birthweight* – coded as a continuous variable.
7. *Maternal education* – coded as a categorical variable with four categories of response, less than O-level, O-level, A-level, and degree or above.

We also estimated cell type composition using the Houseman method for all three ages as part of the *meffil* pipeline [4, 8]. All estimated cell type proportions were included in downstream analyses and regressions.

**Structured Life Course Modeling Approach (SLCMA)**

We tested time-dependent associations for each adversity using the timepoints shown in **Fig. 1**. In the first step, the SLCMA selected the timepoint or additive hypothesis (accumulation; recency) that explained the most variation in a given CpG for each type of adversity (seven separate analyses of 302,581 CpGs). We interpreted the model selected by the SLCMA through six separate life course hypotheses, including four sensitive periods hypotheses that encoded exposure to each childhood adversity during:

1. *very early childhood* – hypothesis selected by the SLCMA fell within the ages of 0-2 (before 36 months);
2. *early childhood (ages 3-5)* – hypothesis selected by the SLCMA fell within the ages of 3-5 (61 months or before);
3. *middle childhood* – hypothesis selected by the SLCMA fell within the ages of 6-7 (84 months or before);
4. *late childhood* – hypothesis selected by the SLCMA fell within the ages of 8-11 (after 84 months);
5. *accumulation* – total number exposures across childhood, ranging from 0-8 total exposures, depending on the adversity analyzed;
6. *recency* – total number of exposures weighted by age when the adversity was measured.

In the second stage of the SCLMA, we used selective inference to perform post-selection inference[9] and adjusted for covariates using the Frisch-Waugh-Lovell theorem[10], shown to improve statistical power in penalized regression analyses[11, 12]. Only complete cases (i.e., individuals with non-missing covariate and exposure data from ages 0-11) were analyzed for each adversity (**Fig. 1**).

**Biological implications of loci associated with childhood adversity identified from SLCMA**

To further understand the biological implications of significant loci, we investigated the biological implications of findings from SLCMA in four different ways (**Table S4**).

First, we assessed the enrichment of regulatory elements in top loci compared to all analyzed loci using chi-squared tests. Both FDR-significant and $R^2$-threshold loci were overrepresented in enhancers (**FDR**: $\chi^2$=5.1, p=0.034; **$R^2$**: $\chi^2$=7.1, p=0.008), but not gene promoters (**FDR**: $\chi^2$=1.9, p=0.17; **$R^2$**: $\chi^2$=0.46, p=0.17; **Fig. S2A**). These loci were also enriched for regions away from CpG islands (‘Open Sea’), rather than CpG Islands, shores, or shelves (**FDR**: $\chi^2$=13.3, p=0.021; **$R^2$**: $\chi^2$=13.6, p=0.018; **Fig. S2B**). Overall, top loci showed higher representation in regions of lower CpG density, suggesting these genomic regions may be more responsive to childhood adversity.

Second, we examined the correlation of DNAm at the top loci in blood and four different brain regions using the Blood Brain DNA Methylation Comparison Tool[13]. Most FDR-significant loci (17/22) had weak, but positive correlations between brain and blood (prefrontal cortex $r_{avg}$=0.05, range=-0.19-0.65; entorhinal cortex $r_{avg}$=0.06, range=-0.24-0.60; superior temporal gyrus $r_{avg}$ =0.05, range=-0.18-0.61; cerebellum $r_{avg}$=0.06, range=-0.14-0.54)(**Table S5**; **Fig. S3**)[13]. Similarly, most $R^2$-threshold loci (28/41) also had weak, but positive correlations, which were, on average, larger than those for the FDR loci (prefrontal cortex $r_{avg}$=0.11, range= -0.19-0.95; entorhinal cortex $r_{avg}$=0.11, range=-0.24-0.95; superior temporal gyrus $r_{avg}$ =0.09, range=-0.21-0.94; cerebellum $r_{avg}$=0.09, range=-0.20-0.97). Thus, adversity-induced alterations to blood DNAm levels may reflect similar changes in the central nervous system.

Third, we analyzed the enrichment of biological processes in top loci using gene ontology (GO) terms from the DAVID tool [14, 15]. Although none reached significance, eight distinct clusters of biological processes were overrepresented in FDR-significant loci (n=21 genes)[14, 15]. These clusters were implicated in abiotic stimulus, development, ion transport, and cellular regulation of biosynthetic processes (**Fig. S4**). By contrast, 18 clusters were identified for $R^2$-threshold loci, which were involved in development, MAPK activity, muscle development, and immunity. These results suggest that different types of childhood adversity may act through diverse biological processes, rather than a concerted network of pathways.

Fourth, we assessed the evolutionary constraint of genes linked to top loci using data from the Exome Aggregation Consortium [16]. Genes linked to top loci showed little evidence of evolutionary conservation, as measured by the intolerance to loss-of-function estimates (pLI) from the Exome Aggregation Consortium (**Table S4**; **Fig. S5**). However, 3 FDR-significant genes linked to the accumulation of exposure to one-adult households showed evidence of strong evolutionary constraint (pLI>0.9; *DSP*, *CUX2*, and *STK38L*). Four additional genes with high evolutionary constraint were identified in the $R^2$-threshold loci (*FBXL16*, *PKD2*, *TAF1*, and *XKR6*). Together, these findings highlight a potential role for genes influenced by parental and social environment in human survival and evolution.

**Internal validation of age 15 loci using non-parametric bootstrapping**

The ALSPAC cohort is unique; no longitudinal birth cohorts at present have collected comparable measures of childhood adversity and DNAm. At best, other birth cohort studies with repeated measures of childhood adversity have only collected one timepoint of DNAm during childhood or adolescence, but not both. By contrast, studies with repeated DNAm measures do not have repeated and prospective measures of childhood adversity. As such, we could not complete external replication analyses of the associations we detected between time-varying childhood adversity and DNAm at age 15. In the absence of a cohort in which to replicate our findings, we performed internal validation analyses of our associations using ordinary nonparametric bootstrapping[17].

In brief, the bootstrap involves resampling data with replacement from a given sample[18]. Unlike parametric methods, such as t-test and linear regressions, the bootstrap does not require assumptions of normality nor rely on parameter estimation (e.g., regression coefficients) from the original sample. Rather, the bootstrap relies on the approximations of test statistics, generated by drawing repeated resamples from a given sample – at random – across thousands of iterations. By resampling with replacement, the original sample size is maintained, with some rows of data omitted and others repeated; this process creates multiple random (re)samples of data from the same underlying population. Since the original sample is drawn from the population of interest, each bootstrap resample can be thought of as a new sample of data drawn from the population. In other words, the bootstrap sample differs from the original sample in each iteration at random, while also remaining similar to the general population from which the original sample was collected. As such, bootstrapping can provide insight into whether findings might be replicated in an independent cohort sampled from the same general population.

Here, we performed a random-x bootstrap resampling using the *boot* package in R[19]. For each CpG identified in the analyses of childhood adversity and DNAm at age 15, we performed 10,000 bootstrapped linear regressions of the selected hypothesis (**Table 1**) and DNAm. We included the same covariates as the SLCMA analyses in the bootstrapped models. Effect estimates across the 10,000 bootstraps were averaged to obtain the "bootstrapped effect estimate". 95% confidence intervals were calculated using the normal-theory interval[19].

The results from the bootstrap analyses were nearly identical to those identified in the initial SLCMA analyses (**Table S6**), both in terms of average effect estimates and confidence intervals. The mean difference between effect estimates from the bootstrap and original analyses (i.e., bootstrap bias) across all top loci was 4.57 $x10^{-5}$ ($2.52x10^{-5}$ for FDR-significant loci), with the largest absolute magnitude of difference being 2.03% (comparing the bootstrap to the original effect estimate). In addition, all effect estimates were significant at the 5% level, judged by bootstrap confidence intervals (**Fig. S6**). Confidence intervals were narrower in all but two of the original analyses (linear regression) compared to the bootstrap, suggesting the bootstrap could more precisely assess the effect estimate.

Together, these findings show that our initial results were robust to different analytic subsamples and populations, as well as nonparametric approaches that make fewer distributional assumptions. Thus, our findings may be likely to replicate in independent cohorts.

**Adjusting for exposure to other childhood adversities**

To further determine the specificity of our associations between subtypes of childhood adversity and DNAm patterns at age 15, we performed a set of mutually-adjusted regression analyses. Specifically, we investigated the impact of correcting for exposure to the other six types of childhood adversity on the strength of association between a given measure of childhood adversity and DNAm.

Children in this analytic sample could have been exposed to adversity before, during, or after the sensitive periods we identified. We therefore coded exposure to other types of childhood adversity in five ways, as outlined below. We investigated these five different ways of coding co-occurring adversities to facilitate future replication of our work in datasets that may not be as fine-grained as ALSPAC, as well as narrow down the periods when co-occurring adversities may have the greatest impact on our results.

1. *Exposed to any other childhood adversity between age 1-11* – the full window of potential exposures to childhood adversity;
2. *Exposed to any other childhood adversity between age 1-7* – the window of potential exposures to childhood adversity that would influence age 7 and age 15 DNAm;
3. *Exposed to any other childhood adversity between age 8-11* – the window of potential exposures to childhood adversity that would only influence age 15 DNAm;
4. *Exposed to any other childhood adversity before the SLCMA-selected sensitive period;*
5. *Exposed to any other childhood adversity during the SLCMA-selected sensitive period;*

*NB: for loci with accumulation hypotheses* – #4 and #5 were calculated using the accumulation of all exposures to other adversities from age 1-11.

For each of the 41 adolescent-specific loci, we ran five separate regressions that included the base model (no mutual adjustment; i.e., the model we presented in primary text) and one of the five above variables. The strength of associations for the mutually-adjusted models were compared to the base model associations between the specific childhood adversity and DNAm at age 15. We found that all associations remained significant when correcting for other types of childhood adversity, no matter which mutual-adjustment strategy was employed (FDR<0.05 when correcting for testing 41 loci) (**Fig. 7**).

Associations between the accumulation of exposure to one-adult households and DNAm at age 15 were most attenuated in the mutually-adjusted model, showing between a 1 to 39% reduction in the size of the effect estimate per CpG; the average attenuation for these three CpGs was 9.0% (**Fig. S8**). Similarly, the three loci linked to the recency of exposures to financial hardship also showed stronger effect shifts in mutually adjusted models (range = -28% to 27%, mean = 2.4%). These results are perhaps unsurprising, given that accumulation and recency scores across childhood may be more highly correlated with other exposures to childhood adversity.

By contrast, we observed smaller alterations to the effect of exposures during sensitive period hypotheses when performing these mutual-adjustment analyses, suggesting our sensitive period findings were less prone to the influence of other types of childhood adversity. Of note, mutual-adjustment for other adversities reported during the same sensitive period identified by the SLCMA generally had the greatest effect on the strength of associations (**Fig. S9**). In particular, almost all associations between exposure to one adult households during early childhood, and DNAm at age 15 were attenuated when controlling for co-occurring adversities during the same sensitive period (mean = 8.6% reduction in effect estimate, range = -20.6% to

4.0%; **Fig. S8**). This finding suggests one-adult households may co-occur with other adversities more frequently, particularly during early childhood. Nevertheless, the strength of associations remained fairly stable even when controlling for these co-occurring exposures, indicating that associations remained specific to one-adult households.

Taken together, these results suggest our observed associations between childhood adversity and DNAm at age 15 were mostly specific to each type of childhood adversity and were not the result of other possible co-occurring exposures across childhood. Future studies should further investigate these findings in other cohorts to confirm their robustness and specificity to subtypes of childhood adversity, especially because ALSPAC is a sample where few children were simultaneously exposed to multiple types of adversity (see correlations in **Table S3** and **Fig. S1**).

**Testing for potential confounding effects of the relationship between childhood adversity and DNA methylation levels at age 7 and 15**

Given that our observed associations between childhood adversity and DNAm at age 15 were not present at age 7, we hypothesized that these emergent effects could be influenced by confounding structures of the data, whereby other factors might be driving these adolescent-specific associations. As such, we further investigated whether the associations we observed between time-varying childhood adversity and DNA methylation patterns across development were influenced by confounding factors or methodological artifacts that were not included in our models. We approached the issue of confounders using two approaches, outlined in **Fig. S10,** focusing on the 41 associations that were identified in age 15 DNAm.

*Early-life confounders of childhood adversity and DNAm at age 7 and 15*

First, we tested whether early-life factors could influence the strength of associations between childhood adversity and DNAm levels at age 7 and 15. To this end, we assessed the impact of removing covariates from our base model (described above) on the estimated effect from a linear regression of time-varying adversity and DNAm levels. When removing individual covariates from the base model, we did not observe any large changes in the effect estimates of the associations between childhood adversity and DNAm at age 15 (**Fig. S11**) or age 7 (**Fig. S12**), except for two CpGs (cg17928317: 37.5% increase; cg27558057: 72.8% decrease). The effect estimates of these two loci changed substantially upon removal of sex as a covariate (**cg17928317**: age 15 $\beta_{base}$=0.079, $\beta_{no\ sex}$=0.108; age 7 $\beta_{base}$ = 0.001, $\beta_{no\ sex}$=0.029; **cg27558057**: age 15 $\beta_{base}$=0.106, $\beta_{no\ sex}$=0.029; age 7 $\beta_{base}$ =0.066, $\beta_{no\ sex}$=-0.024), though we note that both

CpGs are located on chromosome X. As such, some amount of sex-dependent variability is expected due to differences in X chromosome dosage between males and females.

Beyond the covariates included in our base model, we also investigated whether other common confounders may have influenced our observed associations. Here, we assessed the impact of adding the following confounding factors known to influence childhood adversity or DNAm patterns to our base regression model: 1) parental socio-economic position measured at birth, 2) gestational age in weeks, and 3) maternal pre-pregnancy BMI. We investigated these potential confounding factors due to their influence on risk for childhood adversity, as well as their prior associations with longitudinal DNAm patterns[20, 21]. Of note, these factors were omitted from our initial analyses due to their high correlation with other covariates within our base model that are more robust predictors of longitudinal outcomes, such as maternal education, birthweight, maternal age, etc.

Once again, the inclusion of these potential confounders did not substantially influence the strength of associations between childhood adversity and DNAm levels at age 15 (**Fig. S11**) or age 7 (**Fig. S12**). Indeed, only four loci showed a >10% change in their effect estimates upon the inclusion of new covariates, all of which were influenced by the inclusion of maternal pre-pregnancy BMI (two from one-adult households and FDR-significant; two from financial hardship and passing the $R^2$-threshold). Changes less than 10% are generally thought to reflect factors that have little confounding effects [22], although more recent studies suggest that this threshold may be overly conservative [23].

Taken together, these findings suggest the specific associations between time-varying childhood adversity and DNAm at age 15 may not be due to the effects of common confounders or methodological artifacts arising from our current covariates. Furthermore, the associations between adversity and DNAm at age 7 remained null for these 41 loci, further suggesting that the latent effects we observed were unlikely due to common confounders. Nevertheless, it is possible that other unmeasured confounders may influence the relationship between childhood adversity and DNAm at age 15, and thus, our findings should be replicated in other longitudinal birth cohorts with repeated measured of childhood adversity and DNAm.

*Adolescent-specific factors mediating the relationship between childhood adversity and DNAm*

Second, we tested the influence of adolescent-specific factors that could have possibly explained our observed associations. These adolescent-specific factors occurred after childhood adversity and DNAm collection at age 7, but before DNAm collection at age 15 (**Fig. S10**). Because our associations maintained the temporal ordering of exposures preceding the outcome, adolescent-specific confounders should not influence associations with DNAm at age 7. Moreover, confounders are, by definition, linked to both the exposure (adversity) and outcome (DNAm levels at age 15). In the present situation, we could assume that adolescent-specific factors land in the causal path between adversity and DNAm, given that they would occur after adversity and before DNAm.  Given this causal path, potential adolescent-specific confounders could be considered mediators, rather than confounders that can be adjusted in a regression model. As such, we performed causal mediation analyses using the R package *mediation* (version 4.5.0) to determine whether our adolescent-specific association were explained, in part, by potential factors on the causal path. To this end, we assessed whether four biological outcomes previously linked to childhood adversity and/or DNAm patterns significantly mediated our observed associations; our rationale for testing these variables is described below. We corrected for the same covariates as previously described in mediation analyses.

Pubertal onset: Exposure to childhood adversity has been associated with earlier pubertal onset in some studies, including ALSPAC [24]. Puberty is a time of rapid change and development, with concomitant alterations in epigenetic pathways [25]. As such, age at pubertal onset is a plausible candidate to mediate the association between childhood adversity and DNAm levels in adolescence. To estimate pubertal timing, we analyzed the age at peak height velocity, calculated by a method called superimposition by translation and rotation (SITAR), which analyzes height measurements between age 5 and 16 (N=605-654) to identify the age at pubertal onset[26].

We did not identify significant mediation effects for pubertal onset for any of our top 41 loci (lowest p-value = 0.268, cg14455319; **Fig. S13**). Furthermore, when we contrasted our findings to a previous epigenome-wide association study of puberty and gonadal hormone levels, we did not find any overlaps with our 41 adolescent-specific loci[27]. These findings suggest pubertal onset was unlikely to explain adolescent-specific associations.

Body mass index (BMI): We next analyzed BMI measured at age 15 (N=569-618). Prior studies have shown that childhood adversity is linked to obesity and changes in metabolic function[28, 29]. In addition, a recent study of BMI in the ARIES cohort has shown a strong relationship between DNAm and BMI[30]. Although the majority of loci in our analysis showed no significant mediation through BMI at age 15 (**Fig. S14**), 2.67% of the association between exposure to a one adult household in early childhood and DNAm levels at cg16907527 was explained by BMI (p=0.050). Although this association did not survive multiple-test correction, we note this locus is located in *VEGFA*, a gene linked to hyperglycemia and diabetes[31]. Together, these finding suggest BMI was not likely to have substantial confounding effects on our findings.

C-reactive protein (CRP): Childhood adversity has been associated with alterations in inflammatory pathways [32], which, in turn, have been linked to genome-wide DNAm differences

[33, 34]. As such, we assessed the potential role of CRP levels, measured at age 15, as a mediator between childhood adversity and DNAm levels at age 15 (N=491-542). Again, we did not identify any significant mediation effects (**Fig. S15**). Two loci, located in *VEGFA* (cg16907527) and *SLC25A41 (*cg12096528), showed a causal mediation effect with $p<0.05$, suggesting that CRP levels may have slight effects on our associations. Again, these did not survive multiple-test correction for the analysis of 41 loci. Overall, these findings suggest that CRP may not have been an important confounding factor in our analyses.

Adolescent smoking: Smoking and exposure to cigarette smoke is one of the strongest and best-replicated associations with DNAm patterns[35]. In addition, smoking in early adolescence may reflect increased risk-taking behaviors, which are linked to a higher likelihood of exposure to some types of childhood adversity[36]. As such, we investigated daily smoking at age 15 (meaning whether the adolescent smoked every day or not) explained the relationship between childhood adversity and DNAm levels at age 15 (N=566-613). Again, we did not observe any significant mediation effects of smoking on the association between childhood adversity and DNAm at age 15 (**Fig. S16**), suggesting that smoking may not have confounded our findings.

All taken together, these results suggest that our findings were not influenced by these four biological and environmental factors linked to childhood adversity and known to influence DNAm levels. Although we cannot rule out that other pathways may be involved in our adolescent-specific associations, these analyses provide additional support for the direct and latent effects of childhood adversity on the adolescent epigenome.

**Types of DNAm trajectories across development for age 15 loci**

To further refine the patterns of change and stability in the DNAm response to childhood adversity, we characterized the different types of longitudinal DNAm trajectories present in the 41 $R^2$-threshold loci identified from the SCLMA of age 15 DNAm. We first split trajectory types based on the ANOVA of exposure group-by-age interactions, finding two sets of loci: 1) 7 loci that did not show any group-by-age interactions (i.e., stable cluster) and 2) 34 loci with significant group-by-age interactions (FDR<0.05).

Focusing on the second subset, we characterized the patterns of DNAm that could be used to distinguish between different types of DNAm trajectories across development. To this end, we applied a Tukey *post-hoc* test to identify the significant contrasts from the ANOVA of exposure group-by-age interactions for each locus, which included exposure group differences, mean age differences, and exposure group differences *within* and *between* each age. As we were interested in changes across time and age 15-specific patterns, we focused our analyses on a subset of these Tukey contrasts, which included:

1. *mean exposure group differences across all age* – meaning comparisons between individuals exposed during the period selected by the SLCMA (exposed-SP), individuals exposed outside the period selected by the SLCMA (exposed-other), and individuals with no exposure (unexposed);
    a. Exposed-SP versus Exposed-other
    b. Exposed-SP versus Unexposed
    c. Exposed-other versus Unexposed
2. *mean age differences across exposure groups for neighboring ages* – meaning mean differences between age 7 and 0, as well as mean differences between age 15 and 7;

    a. Age 7 versus Age 0
    b. Age 15 versus Age 7
3. *exposure group differences within each age* – meaning differences between exposure groups at age 0, age 7, or age 15.
    a. Age 0-specific differences
        i. Exposed-SP versus Exposed-other
        ii. Exposed-SP versus Unexposed
        iii. Exposed-other versus Unexposed
    b. Age 7- specific differences
        i. Exposed-SP versus Exposed-other
        ii. Exposed-SP versus Unexposed
        iii. Exposed-other versus Unexposed
    c. Age 15-specific differences
        i. Exposed-SP versus Exposed-other
        ii. Exposed-SP versus Unexposed
        iii. Exposed-other versus Unexposed

We recoded these contrasts as categorical variables to reflect whether the differences from the Tukey were significant (0 = $p>0.05$; 1 = $p<0.05$). We then performed divisive hierarchical clustering using a dissimilarity matrix for these categorical patterns (i.e., 0/1 based on significance) using the *cluster* package in R [37]. We selected the number of distinct types of trajectories based on the inflection point of the sum of squares (lowest without meaningful decrease), with no more than one trajectory type with one CpG (**Fig. S17**). This step resulted in six distinct types of DNAm trajectories (**Fig. S18**), which showed distinct profiles of age, group, and group-by-age differences (**Fig. S19**). Trajectories were plotted using cell-type corrected DNAm values and complete cases for covariates measured at birth (age 0: N = 559-616; age 7: N = 613-668; age 15: N= 609-665; sample sizes varied by adversity; **Fig. S20**).

For the seven loci without exposure group-by-age interactions, we identified slight differences between youths exposed during a sensitive period and those who were unexposed at age 7, which fully emerged by age 15 (i.e., stable).

Finally, we did not identify any differences in the enrichment of DNAm trajectories between loci in the FDR-significant and $R^2$-threshold subsets ($\chi^2$=1.92, p =0.86; **Fig. S21**). These findings further emphasize that p-values do not show the whole picture, though additional differences may emerge when thresholds are relaxed further.

**Investigating adversity-DNAm relationships within a threat and deprivation paradigm**

To investigate potential differences between in sensitive period enrichment among our top loci in the context of threat versus deprivation-type exposures[38-40], we used the following definitions to classify our adversities into this established paradigm:

A. *Threat:* Threat exposures are defined as "experiences that represent a threat to one's physical integrity"[38]. Based on this definition, exposures to 1) caregiver physical or emotional abuse, and/or 2) physical or sexual abuse (by anyone) were categorized as threat-type exposures.

B. *Deprivation:* Deprivation exposures are defined as the "absence of expected environmental inputs and complexity"[38]. Based on this definition, exposures to 1) family instability, 2) financial hardship, 3) maternal psychopathology, 4) neighborhood disadvantage, and/or 5) one adult households were categorized as deprivation-type exposures.

Following the classification of adversities into these paradigms, we investigated differential patterns of sensitive period enrichment for the 41 top loci identified at age 15 and 22 loci that passed an FDR<0.05 threshold (**Fig. S22**). Although there were differences in the number of adversities contributing to these two exposure paradigms, we observed more loci associated with a deprivation paradigm (34 loci) than a threat paradigm (7 loci). Furthermore, both exposure paradigms had more associations with exposure during early childhood than other exposure periods or models. However, loci associated with threat exposures were clustered mainly within early childhood, while loci associated with deprivation exposures were more distributed across time periods ($\chi^2$=7, p =0.32). Together, these findings suggest that deprivation-type exposures during early childhood may have greater impacts on adolescent DNAm profiles, but these effects can be further refined by investigating specific types of childhood adversity.

### SUPPLEMENTAL TABLES

**Table S1. Summary of the childhood adversity variables analyzed in the present study.**

| Adversity | Respondent | Questionnaire items | Exposure classification | Assessment timepoints |
|---|---|---|---|---|
| **Caregiver physical or emotional abuse** | Mother and partner | 1) your partner was physically cruel to your children,<br>2) you were physically cruel to your children,<br>3) your partner was emotionally cruel to your children,<br>4) you were emotionally cruel to your children. | Exposed: mother, the partner, or both, endorsed any of the items.<br><br>Unexposed: any negative response and no positive response.<br><br>Missing: all questions unanswered. | 8 months, 1.75 years, 2.75 years, 4 years, 5 years, 6 years, 9 years, and 11 years |
| **Sexual or physical abuse** | Mother | 1) an item asking if the child was exposed to either sexual or physical abuse from anyone. | Exposed: an affirmative response was provided to either item.<br><br>Unexposed: any negative response was available and no positive response was provided.<br><br>Missing: both questions unanswered. | 1.5 years, 2.5 years, 3.5 years, 4.75 years, 5.75 years, 6.75 years, and 8 years |
| **Maternal psychopathology** | Mother | 1) the Crown-Crisp Experiential Index (CCEI), assessing anxiety and depression,<br>2) the Edinburgh Postnatal Depression Scale (EPDS),<br>3) a question asking about suicide attempts in the past 1.5 years. | Exposed: one or more of the following criteria was met:<br>1) CCEI depression score > 9<br>2) CCEI anxiety score > 10<br>3) EPDS score > 12 | 8 months, 1.75 years, 2.75 years, 5 years, 6 years, and 11 years |

| | | | | |
|---|---|---|---|---|
| | | | 4) a suicide attempt since the time of the last interview<br><br>Unexposed: none of the above criteria above were met and none of the scores were missing.<br><br>Missing: Any of the prorated scales or questions were missing. | |
| **One adult in the household** | Mother | 1) an item asking about the number of adults (>18 years of age) living in the household. | Exposed: fewer than two adults were residing in the household.<br><br>Unexposed: two adults or more were residing in the household.<br><br>Missing: question unanswered. | 8 months, 1.75 years, 2.75 years, 4 years, 7 years, 8 years, and 10 years |
| **Family instability** | Mother | Child<br>1) taken into care,<br>2) separated from their mother for two or more weeks,<br>3) separated from their father for two or more weeks,<br>4) acquired a new parent. | Exposed: at least two of these events occurred at a single time point.<br><br>Unexposed: none of the events occurred at a single time point and no questions were missing.<br><br>Missing: any question was unanswered. | 1.5 years, 2.5 years, 3.5 years, 4.75 years, 5.75 years, 6.75 years, and 8 years |

| | | | | |
|---|---|---|---|---|
| **Financial hardship** | Mother | Family had difficulty affording the following items, coded on a Likert-type scale (1=not difficult; 2=slightly difficult; 3=fairly difficult; 4=very difficult):<br>1) items for the child,<br>2) rent or mortgage,<br>3) heating,<br>4) clothing,<br>5) food. | Exposed: mothers reported at least fair difficulty for three or more items at each time point.<br><br>Unexposed: mothers reported on all five items, but the above criterion was not met.<br><br>Missing: any question unanswered. | 8 months, 1.75 years, 2.75 years, 5 years, 7 years, and 11 years |
| **Neighborhood disadvantage** | Mother | The following problems happened in the neighborhood (2=serious problem, 1=minor problem, 0=not a problem or no opinion):<br>1) noise from other homes,<br>2) noise from the street,<br>3) garbage on the street,<br>4) dog dirt,<br>5) vandalism,<br>6) worry about burglary,<br>7) mugging,<br>8) disturbance from youth. | Exposed: scores >=8 of the total sum of questions, corresponding to the 95th percentile of exposure.<br><br>Unexposed: scores were <8 and no questions were missing.<br><br>Missing: any question unanswered. | 1.75 years, 2.75 years, 5 years, 7 years, and 10 years |

**Table S2. Distribution of covariates in the total ALSPAC sample, ARIES subsample, and among those exposed to any adversity between age 0-11.**

| | ALSPAC (N=15646) | ARIES* (N=966) | Exposed to any adversity (N=647) | ALSPAC vs. ARIES | ALSPAC vs. Exposed | ARIES vs. Exposed |
|---|---|---|---|---|---|---|
| | N (%) | N (%) | N (%) | | | χ2 test p-value |
| Sex | | | | 0.068 | 0.11 | 0.99 |
| Male | 7542 (51.3) | 466 (48.2) | 311 (48.1) | | | |
| Female | 7152 (48.7) | 500 (51.8) | 336 (51.9) | | | |
| Race/Ethnicity | | | | 0.007 | 0.38 | 0.28 |
| White | 11488 (94.9) | 900 (97) | 26 ( 4.2) | | | |
| Non-white | 611 (5.1) | 28 (3) | 596 (95.8) | | | |
| Maternal education | | | | <0.001 | <0.001 | 0.49 |
| less than O-level | 3735 (30) | 152 (16.1) | 118 (18.6) | | | |
| O-level | 4303 (34.6) | 321 (34) | 202 (31.8) | | | |
| A-level | 2795 (22.5) | 279 (29.5) | 194 (30.6) | | | |
| Degree or Above | 1603 (12.9) | 193 (20.4) | 121 (19.1) | | | |
| Maternal age at birth | | | | <0.001 | <0.001 | 0.68 |
| Ages 15-19 | 650 (4.6) | 9 (0.9) | 9 ( 1.4) | | | |
| Ages 20-35 | 12363 (88.4) | 858 (89.4) | 572 (88.7) | | | |
| Age 36+ | 968 (6.9) | 93 (9.7) | 64 ( 9.9) | | | |
| Smoking during pregnancy | | | | <0.001 | <0.001 | 0.19 |
| Smoker | 2577 (21.2) | 98 (10.7) | 80 (13.1) | | | |
| Non-smoker | 9565 (78.8) | 814 (89.3) | 532 (86.9) | | | |
| Previous pregnancies | | | | 0.004 | 0.1 | 0.96 |
| 0 | 5800 (44.7) | 439 (47.1) | 295 (47.0) | | | |
| 1 | 4550 (35) | 346 (37.1) | 229 (36.5) | | | |
| 2 | 1860 (14.3) | 113 (12.1) | 77 (12.3) | | | |
| 3+ | 772 (5.9) | 34 (3.6) | 26 ( 4.1) | | | |
| Birthweight | | | | <0.001 | <0.001 | 0.98 |
| < 3000 | 3649 (24.8) | 149 (15.4) | 101 (15.6) | | | |
| 3000 - 3499 | 4924 (33.5) | 339 (35.1) | 228 (35.2) | | | |
| 3500 - 3999 | 4382 (29.8) | 331 (34.3) | 216 (33.4) | | | |
| >= 4000 | 1735 (11.8) | 147 (15.2) | 102 (15.8) | | | |

*The ARIES subsample with DNA methylation data collected at age 15-17, without twins. P-values, used to evaluate whether distributions differed across each sample comparison, were determined by chi-square tests. Maternal education values are presented from lowest level of education (less than O-level) to highest (degree or above).

**Table S3. Prevalence and correlations between adversities occurring from age 0-11.**

| Adversity | Prevalence (% any exposure)[1] | Average within adversity correlation[2] | Average correlation with other adversities[3] |
|---|---|---|---|
| Caregiver physical or emotional abuse | 18.1 | 0.562 | 0.137 |
| Sexual or physical abuse (by anyone) | 15.1 | 0.402 | 0.090 |
| Family instability | 24.4 | 0.597 | 0.153 |
| Financial hardship | 15.9 | 0.357 | -0.035 |
| Maternal psychopathology | 34.8 | 0.611 | 0.161 |
| Neighborhood disadvantage | 16.1 | 0.741 | 0.112 |
| One adult in the household | 17.9 | 0.786 | 0.127 |

[1]Prevalence of any exposure to adversity between the ages of 0 and 11.
[2]Average tetrachoric correlation of exposure to adversity between different timepoints across development.
[3]Average tetrachoric correlation of exposure to different types of adversity across development.

### Table S4. Annotated loci identified at age 15.

| Adversity | Timing | Age (years) | CpG | Chr | Coordinate | Nearest Gene | Distance to gene | Relation to CGI | Enhancer | Promoter | pLI |
|---|---|---|---|---|---|---|---|---|---|---|---|
| Caregiver physical or emotional abuse | Early childhood | 5 | cg14855874 | 4 | 102712397 | BANK1 | 0 | S_Shore | 1 | 0 | 1.8E-10 |
| | | | cg15454534 | 1 | 248569605 | OR2T1 | 0 | OpenSea | 0 | 0 | 7.3E-07 |
| | | | cg06215562 | 13 | 82344645 | | | OpenSea | 1 | 0 | |
| Sexual or physical abuse (by anyone) | Early childhood | 3.5 | **cg26970800** | **11** | **59614212** | **CBLIF** | **1237** | **OpenSea** | **0** | **0** | |
| | | | **cg15723468** | **1** | **230387268** | **GALNT2** | **0** | **OpenSea** | **0** | **0** | **8.8E-01** |
| | | | **cg17928317** | **X** | **140982278** | **MAGEC3** | **0** | **OpenSea** | **0** | **0** | **3.2E-08** |
| | Late childhood | 8 | cg27558057 | X | 70712724 | TAF1 | 0 | Island | 0 | 1 | 1.00 |
| Family instability | Very early childhood | 2.5 | cg02735620 | 4 | 88950514 | PKD2 | 0 | OpenSea | 1 | 0 | 1.00 |
| Financial hardship | Very early childhood | 0.66 | cg14455319 | 11 | 113258908 | ANKK1 | 0 | S_Shore | 1 | 0 | 2.5E-08 |
| | | | cg13204236 | 2 | 47476732 | STPG4 | 72991 | OpenSea | 1 | 0 | |
| | Early childhood | 5 | cg15037420 | 19 | 48474386 | BSPH1 | 0 | OpenSea | 0 | 0 | |
| | | | cg06410970 | 10 | 81921424 | ANXA11 | 0 | OpenSea | 1 | 0 | 3.5E-06 |
| | Late childhood | 11 | cg02011706 | 16 | 891283 | LMF1 | 12350 | N_Shelf | 0 | 0 | 1.1E-14 |
| | | | cg04659536 | 7 | 4218154 | SDK1 | 0 | OpenSea | 0 | 0 | 5.0E-03 |
| | Recency | | cg17670999 | 8 | 145928398 | ARHGAP39 | 17203 | S_Shelf | 0 | 0 | 1.7E-03 |
| | | | cg25459301 | 8 | 10941183 | XKR6 | 0 | OpenSea | 1 | 0 | 9.6E-01 |
| | | | cg06812747 | 16 | 742426 | FBXL16 | 72 | N_Shore | 0 | 0 | 9.5E-01 |
| Maternal psychopathology | Very early childhood | 2.75 | **cg16813552** | **10** | **103544649** | **OGA** | **0** | **S_Shore** | **0** | **0** | |
| Neighborhood disadvantage | Very early childhood | 2.75 | cg04288299 | 4 | 1988825 | NELFA | 0 | S_Shore | 0 | 0 | 1.7E-01 |
| | | | cg25019631 | 1 | 15850977 | CASP9 | 0 | N_Shore | 0 | 1 | 3.2E-03 |
| | | | cg04224851 | 2 | 43304158 | ZFP36L2 | 145381 | OpenSea | 1 | 0 | 4.6E-01 |
| One adult in the household | Very early childhood | 1.75 | **cg05491478** | **2** | **238621313** | **LRRFIP1** | **0** | **OpenSea** | **0** | **0** | **3.7E-01** |
| | Early childhood | 3.9 | **cg16907527** | **6** | **43744388** | **VEGFA** | **0** | **OpenSea** | **1** | **0** | |
| | | | **cg08818094** | **4** | **26806047** | **TBC1D19** | **49128** | **OpenSea** | **1** | **0** | **1.6E-02** |
| | | | **cg01060989** | **1** | **221945814** | **DUSP10** | **30297** | **OpenSea** | **1** | **0** | **5.0E-01** |
| | | | **cg15814750** | **15** | **53880678** | **WDR72** | **0** | **OpenSea** | **1** | **0** | **1.6E-16** |
| | | | **cg15783822** | **12** | **10999279** | **PRR4** | **0** | **OpenSea** | **0** | **0** | **1.0E-05** |
| | | | **cg15864691** | **7** | **27217606** | **HOXA10** | **0** | **N_Shore** | **0** | **0** | **6.8E-01** |

| | | | | | | | | | | |
|---|---|---|---|---|---|---|---|---|---|---|
| | | **cg02584161** | **6** | **156086665** | | | **OpenSea** | **1** | **0** | |
| | | **cg02810291** | **15** | **85973746** | **AKAP13** | **0** | **OpenSea** | **1** | **0** | **8.5E-01** |
| | | **cg04036644** | **8** | **1200583** | **LOC286083** | **43709** | **OpenSea** | **0** | **0** | |
| | | **cg11811897** | **7** | **47811084** | **PKD1L1** | **3164** | **OpenSea** | **1** | **0** | **1.7E-23** |
| | | **cg15817130** | **5** | **16742179** | **MYO10** | **0** | **OpenSea** | **0** | **0** | **4.0E-03** |
| | | **cg06711254** | **2** | **186924071** | **FSIP2** | **226054** | **OpenSea** | **1** | **0** | **3.2E-08** |
| | | **cg19096460** | **4** | **89490818** | **HERC3** | **22754** | **OpenSea** | **1** | **0** | **7.2E-01** |
| | | cg18980650 | X | 100130547 | NOX1 | 1212 | OpenSea | 0 | 0 | 3.9E-04 |
| | | cg27504269 | 12 | 21524305 | SLCO1A2 | 0 | OpenSea | 0 | 0 | 5.6E-15 |
| Late childhood | 10 | **cg12096528** | **19** | **6427642** | **SLC25A41** | **0** | **S_Shore** | **0** | **0** | **6.0E-05** |
| Accumulation | | **cg00807464** | **12** | **111618977** | **CUX2** | **0** | **OpenSea** | **0** | **0** | **1.00** |
| | | **cg10420609** | **6** | **7538349** | **DSP** | **3519** | **N_Shelf** | **0** | **0** | **1.00** |
| | | **cg14579651** | **12** | **27429400** | **STK38L** | **0** | **OpenSea** | **1** | **0** | **9.7E-01** |

* CGI = CpG Island; Chr = chromosome; pLI = probability of intolerance to loss of function (Exome Aggregation Consortium). Bolded loci passed a 5% FDR threshold of in the original analysis.

**Table S5. Correlation of DNA methylation in brain and blood for age 15 loci (data from Hannon et al. 2015).**

| Adversity | Timing | Age (years) | CpG | PFC | EC | STG | CER |
|---|---|---|---|---|---|---|---|
| Caregiver physical or emotional abuse | Early childhood | 5 | cg14855874 | 0.213 | 0.269 | 0.444 | 0.239 |
| | | | cg15454534 | 0.059 | 0.072 | -0.033 | 0.145 |
| | | | cg06215562 | 0.068 | 0.014 | -0.023 | -0.067 |
| Sexual or physical abuse (by anyone) | Early childhood | 3.5 | **cg26970800** | **-0.097** | **0.016** | **0.067** | **-0.029** |
| | | | **cg15723468** | **0.035** | **-0.106** | **-0.024** | **0.103** |
| | | | **cg17928317** | **0.649** | **0.600** | **0.610** | **0.538** |
| | Late childhood | 8 | cg27558057 | 0.950 | 0.947 | 0.914 | 0.882 |
| Family instability | Very early childhood | 2.5 | cg02735620 | -0.061 | -0.027 | 0.119 | -0.071 |
| Financial hardship | Very early childhood | 0.66 | cg14455319 | 0.318 | 0.246 | 0.406 | 0.074 |
| | | | cg13204236 | -0.025 | 0.091 | 0.001 | -0.101 |
| | Early childhood | 5 | cg15037420 | 0.065 | -0.002 | -0.069 | 0.057 |
| | | | cg06410970 | -0.083 | -0.003 | 0.112 | -0.024 |
| | Late childhood | 11 | cg02011706 | 0.062 | 0.141 | 0.084 | 0.207 |
| | | | cg04659536 | 0.952 | 0.953 | 0.935 | 0.968 |
| | Recency | | cg17670999 | -0.039 | 0.089 | 0.139 | -0.199 |
| | | | cg25459301 | 0.390 | 0.228 | 0.059 | 0.211 |
| | | | cg06812747 | 0.075 | -0.107 | -0.037 | -0.158 |
| Maternal psychopathology | Very early childhood | 2.75 | **cg16813552** | **0.020** | **-0.035** | **0.019** | **0.173** |
| Neighborhood disadvantage | Very early childhood | 2.75 | cg04288299 | -0.137 | -0.007 | -0.213 | -0.192 |
| | | | cg25019631 | -0.043 | 0.017 | -0.037 | -0.047 |
| | | | cg04224851 | 0.201 | 0.092 | -0.147 | 0.050 |
| One adult in the household | Very early childhood | 1.75 | **cg05491478** | **-0.085** | **0.112** | **-0.058** | **0.057** |
| | Early childhood | 3.9 | **cg16907527** | **0.066** | **-0.008** | **-0.062** | **0.051** |
| | | | **cg08818094** | **0.088** | **0.041** | **0.151** | **-0.086** |
| | | | **cg01060989** | **0.034** | **-0.038** | **0.076** | **-0.033** |
| | | | **cg15814750** | **-0.020** | **-0.241** | **0.004** | **-0.029** |
| | | | **cg15783822** | **0.041** | **-0.005** | **0.151** | **0.027** |
| | | | **cg15864691** | **0.034** | **0.085** | **-0.074** | **-0.138** |

| | | | | | | |
|---|---|---|---|---|---|---|
| | | **cg02584161** | **0.166** | **-0.024** | **0.115** | **0.065** |
| | | **cg02810291** | **-0.185** | **0.187** | **0.134** | **0.058** |
| | | **cg04036644** | **-0.081** | **0.353** | **0.140** | **0.069** |
| | | **cg11811897** | **0.054** | **-0.034** | **0.031** | **0.106** |
| | | **cg15817130** | **0.090** | **0.064** | **-0.036** | **0.167** |
| | | **cg06711254** | **0.081** | **0.058** | **-0.133** | **0.152** |
| | | **cg19096460** | **0.044** | **0.139** | **-0.036** | **0.002** |
| | | cg18980650 | 0.375 | 0.352 | 0.177 | 0.255 |
| | | cg27504269 | -0.001 | -0.066 | -0.072 | 0.072 |
| Late childhood | 10 | **cg12096528** | **0.135** | **0.100** | **-0.181** | **-0.118** |
| Accumulation | | **cg00807464** | **-0.070** | **0.270** | **0.008** | **0.093** |
| | | **cg10420609** | **0.064** | **-0.095** | **0.039** | **-0.023** |
| | | **cg14579651** | **0.032** | **-0.117** | **0.097** | **0.043** |

PFC = prefrontal cortex; EC = entorhinal cortex; STG = superior temporal gyrus; CER = cerebellum. Values represent the correlation between DNA methylation levels in blood and the specified brain regions, as reported by Hannon et al., 2015. Bolded loci passed a 5% FDR in the original analysis.

**Table S6. Associations between childhood adversity and age 15 DNAm using non-parametric bootstrap analyses**

| Adversity | Timing | Age (years) | CpG | Original effect estimate[1] | Bootstrap effect estimate[2] | Bootstrap bias[3] | % difference[4] |
|---|---|---|---|---|---|---|---|
| Caregiver physical or emotional abuse | Early childhood | 5 | cg14855874 | 3.01E-02 | 3.02E-02 | 6.81E-05 | -0.23% |
| | | | cg15454534 | -1.64E-02 | -1.64E-02 | 3.21E-06 | 0.02% |
| | | | cg06215562 | -2.11E-02 | -2.10E-02 | 2.27E-05 | 0.11% |
| Sexual or physical abuse (by anyone) | Early childhood | 3.5 | **cg26970800** | **-5.47E-02** | **-5.45E-02** | **1.90E-04** | **0.35%** |
| | | | **cg15723468** | **-4.52E-02** | **-4.51E-02** | **1.10E-04** | **0.24%** |
| | | | **cg17928317** | **7.56E-02** | **7.54E-02** | **-2.07E-04** | **0.27%** |
| | Late childhood | 8 | cg27558057 | 1.07E-01 | 1.05E-01 | -1.86E-03 | 1.74% |
| Family instability | Very early childhood | 2.5 | cg02735620 | -1.97E-02 | -1.97E-02 | -2.86E-05 | -0.14% |
| Financial hardship | Very early childhood | 0.66 | cg14455319 | 5.29E-02 | 5.26E-02 | -3.25E-04 | 0.61% |
| | | | cg13204236 | -3.73E-02 | -3.73E-02 | -1.96E-05 | -0.05% |
| | Early childhood | 5 | cg15037420 | -3.50E-02 | -3.50E-02 | 6.23E-05 | 0.18% |
| | | | cg06410970 | -3.41E-02 | -3.41E-02 | -2.45E-05 | -0.07% |
| | Late childhood | 11 | cg02011706 | -6.39E-02 | -6.44E-02 | -4.63E-04 | -0.72% |
| | | | cg04659536 | -2.78E-02 | -2.78E-02 | 3.09E-06 | 0.01% |
| | Recency | | cg17670999 | -2.10E-03 | -2.06E-03 | 4.27E-05 | 2.03% |
| | | | cg25459301 | -2.81E-03 | -2.76E-03 | 5.18E-05 | 1.84% |
| | | | cg06812747 | -2.75E-03 | -2.75E-03 | 3.38E-06 | 0.12% |
| Maternal psychopathology | Very early childhood | 2.75 | **cg16813552** | **-1.52E-02** | **-1.52E-02** | **6.59E-06** | **0.04%** |
| Neighborhood disadvantage | Very early childhood | 2.75 | cg04288299 | -2.06E-02 | -2.07E-02 | -5.12E-05 | -0.25% |
| | | | cg25019631 | 4.43E-02 | 4.44E-02 | 1.29E-04 | -0.29% |
| | | | cg04224851 | -1.43E-02 | -1.43E-02 | -2.02E-05 | -0.14% |
| One adult in the household | Very early childhood | 1.75 | **cg05491478** | **-2.76E-02** | **-2.75E-02** | **5.17E-05** | **0.19%** |
| | Early childhood | 3.9 | **cg16907527** | **-3.16E-02** | **-3.17E-02** | **-1.40E-04** | **-0.44%** |
| | | | **cg08818094** | **-5.03E-02** | **-5.02E-02** | **4.75E-05** | **0.09%** |
| | | | **cg01060989** | **-3.15E-02** | **-3.15E-02** | **-3.71E-05** | **-0.12%** |
| | | | **cg15814750** | **-4.14E-02** | **-4.13E-02** | **8.80E-05** | **0.21%** |
| | | | **cg15783822** | **-2.21E-02** | **-2.21E-02** | **3.63E-05** | **0.16%** |

| | | | | | | |
|---|---|---|---|---|---|---|
| | | **cg15864691** | **-1.81E-02** | **-1.80E-02** | **5.27E-05** | **0.29%** |
| | | **cg02584161** | **-5.93E-02** | **-5.93E-02** | **-4.89E-05** | **-0.08%** |
| | | **cg02810291** | **-2.34E-02** | **-2.33E-02** | **7.06E-05** | **0.30%** |
| | | **cg04036644** | **-2.62E-02** | **-2.61E-02** | **6.66E-05** | **0.25%** |
| | | **cg11811897** | **-4.83E-02** | **-4.81E-02** | **2.23E-04** | **0.46%** |
| | | **cg15817130** | **-3.81E-02** | **-3.81E-02** | **-3.42E-06** | **-0.01%** |
| | | **cg06711254** | **-5.80E-02** | **-5.80E-02** | **4.05E-05** | **0.07%** |
| | | **cg19096460** | **-2.49E-02** | **-2.50E-02** | **-8.09E-05** | **-0.32%** |
| | | cg18980650 | -3.68E-02 | -3.69E-02 | -1.77E-04 | -0.48% |
| | | cg27504269 | -4.06E-02 | -4.05E-02 | 1.55E-04 | 0.38% |
| Late childhood | 10 | **cg12096528** | **-1.66E-02** | **-1.65E-02** | **5.63E-05** | **0.34%** |
| Accumulation | | **cg00807464** | **3.21E-03** | **3.18E-03** | **-3.37E-05** | **1.05%** |
| | | **cg10420609** | **-1.45E-02** | **-1.45E-02** | **1.36E-05** | **0.09%** |
| | | **cg14579651** | **-1.29E-02** | **-1.28E-02** | **5.16E-05** | **0.40%** |

[1] Effect estimate from the original linear regression of childhood adversity and DNAm at age 15 in the full ALSPAC sample.
[2] Average of effect estimates from the 10,000 bootstrapped analyses of childhood adversity and DNAm at age 15.
[3] Difference in effect estimates between the bootstrapped and original sample.
[4] Percent change in absolute effect estimate between the original and bootstrapped analyses.
*Bolded loci passed a 5% FDR threshold in the original analysis.

**Table S7. Sensitivity analysis of DNA methylation at birth (cord blood) for loci identified at age 15.**

| Adversity | Timing | Age (years) | CpG | DNAm unexposed[1] | DNAm exp. SP[2] | Δ DNAm[3] | Effect estimate[4] | SE* | P-value | FDR |
|---|---|---|---|---|---|---|---|---|---|---|
| Caregiver physical or emotional abuse | Early childhood | 5 | cg14855874 | 0.099 | 0.112 | 0.013 | 0.014 | 0.007 | 5.60E-02 | 3.95E-01 |
| | | | cg15454534 | 0.866 | 0.864 | -0.003 | -0.003 | 0.005 | 6.06E-01 | 8.78E-01 |
| | | | cg06215562 | 0.830 | 0.825 | -0.005 | -0.005 | 0.005 | 3.52E-01 | 7.10E-01 |
| Sexual or physical abuse (by anyone) | Early childhood | 3.5 | **cg26970800** | **0.890** | **0.901** | **0.011** | **0.012** | **0.013** | **3.65E-01** | **7.10E-01** |
| | | | **cg15723468** | **0.849** | **0.835** | **-0.014** | **-0.015** | **0.008** | **5.80E-02** | **3.95E-01** |
| | | | **cg17928317** | **0.690** | **0.721** | **0.032** | **-0.019** | **0.020** | **3.49E-01** | **7.10E-01** |
| | Late childhood | 8 | cg27558057 | 0.242 | 0.231 | -0.012 | 0.076 | 0.024 | 2.09E-03 | 8.56E-02 |
| Family instability | Very early childhood | 2.5 | cg02735620 | 0.880 | 0.881 | 0.001 | 0.000 | 0.005 | 9.86E-01 | 9.86E-01 |
| Financial hardship | Very early childhood | 0.66 | cg14455319 | 0.254 | 0.281 | 0.027 | 0.028 | 0.012 | 1.54E-02 | 3.15E-01 |
| | | | cg13204236 | 0.858 | 0.866 | 0.007 | 0.008 | 0.007 | 2.83E-01 | 7.10E-01 |
| | Early childhood | 5 | cg15037420 | 0.774 | 0.763 | -0.012 | -0.012 | 0.008 | 1.39E-01 | 5.39E-01 |
| | | | cg06410970 | 0.843 | 0.857 | 0.015 | 0.015 | 0.009 | 9.08E-02 | 4.65E-01 |
| | Late childhood | 11 | cg02011706 | 0.837 | 0.822 | -0.014 | -0.016 | 0.019 | 3.99E-01 | 7.11E-01 |
| | | | cg04659536 | 0.898 | 0.892 | -0.005 | -0.007 | 0.007 | 3.53E-01 | 7.10E-01 |
| | Recency | | cg17670999 | 0.807 | 0.807 | 0.000 | 0.000 | 0.000 | 6.21E-01 | 8.78E-01 |
| | | | cg25459301 | 0.757 | 0.765 | 0.009 | 0.001 | 0.001 | 1.27E-01 | 5.39E-01 |
| | | | cg06812747 | 0.819 | 0.817 | -0.003 | -0.001 | 0.001 | 3.01E-01 | 7.10E-01 |
| Maternal psychopathology | Very early childhood | 2.75 | **cg16813552** | **0.899** | **0.896** | **-0.003** | **-0.004** | **0.003** | **1.83E-01** | **6.25E-01** |
| Neighborhood disadvantage | Very early childhood | 2.75 | cg04288299 | 0.912 | 0.921 | 0.010 | 0.002 | 0.005 | 7.23E-01 | 8.84E-01 |
| | | | cg25019631 | 0.227 | 0.228 | 0.001 | 0.004 | 0.011 | 7.28E-01 | 8.84E-01 |
| | | | cg04224851 | 0.905 | 0.903 | -0.002 | -0.001 | 0.003 | 7.58E-01 | 8.88E-01 |
| One adult in the household | Very early childhood | 1.75 | **cg05491478** | **0.900** | **0.903** | **0.003** | **0.002** | **0.008** | **8.16E-01** | **9.24E-01** |
| | Early childhood | 3.9 | **cg16907527** | **0.840** | **0.848** | **0.008** | **0.006** | **0.006** | **3.68E-01** | **7.10E-01** |
| | | | **cg08818094** | **0.832** | **0.834** | **0.001** | **-0.001** | **0.011** | **9.50E-01** | **9.86E-01** |
| | | | **cg01060989** | **0.809** | **0.814** | **0.005** | **0.005** | **0.007** | **4.64E-01** | **7.61E-01** |
| | | | **cg15814750** | **0.738** | **0.755** | **0.018** | **0.016** | **0.008** | **4.25E-02** | **3.95E-01** |
| | | | **cg15783822** | **0.859** | **0.858** | **-0.001** | **0.001** | **0.005** | **8.77E-01** | **9.46E-01** |
| | | | **cg15864691** | **0.899** | **0.903** | **0.004** | **0.004** | **0.005** | **3.81E-01** | **7.10E-01** |

| | | | | | | | | | |
|---|---|---|---|---|---|---|---|---|---|
| | | **cg02584161** | **0.650** | **0.654** | **0.004** | **0.003** | **0.014** | **8.34E-01** | **9.24E-01** |
| | | **cg02810291** | **0.849** | **0.858** | **0.009** | **0.010** | **0.005** | **3.38E-02** | **3.95E-01** |
| | | **cg04036644** | **0.889** | **0.889** | **0.001** | **-0.002** | **0.006** | **7.31E-01** | **8.84E-01** |
| | | **cg11811897** | **0.737** | **0.728** | **-0.010** | **-0.011** | **0.011** | **3.14E-01** | **7.10E-01** |
| | | **cg15817130** | **0.787** | **0.782** | **-0.004** | **-0.006** | **0.007** | **4.23E-01** | **7.22E-01** |
| | | **cg06711254** | **0.711** | **0.698** | **-0.013** | **-0.015** | **0.010** | **1.45E-01** | **5.39E-01** |
| | | **cg19096460** | **0.843** | **0.841** | **-0.003** | **-0.003** | **0.006** | **6.16E-01** | **8.78E-01** |
| | | cg18980650 | 0.795 | 0.791 | -0.004 | 0.003 | 0.008 | 7.21E-01 | 8.84E-01 |
| | | cg27504269 | 0.748 | 0.752 | 0.004 | 0.003 | 0.008 | 7.33E-01 | 8.84E-01 |
| Late childhood | 10 | **cg12096528** | **0.877** | **0.886** | **0.009** | **0.009** | **0.005** | **6.74E-02** | **3.95E-01** |
| Accumulation | | **cg00807464** | **0.052** | **0.052** | **0.001** | **0.000** | **0.001** | **9.86E-01** | **9.86E-01** |
| | | **cg10420609** | **0.555** | **0.559** | **0.004** | **0.001** | **0.002** | **5.81E-01** | **8.78E-01** |
| | | **cg14579651** | **0.615** | **0.611** | **-0.004** | **-0.002** | **0.002** | **2.85E-01** | **7.10E-01** |

[1]DNAm unexp. = mean DNA methylation levels in individuals with no exposure to adversity from age 0 to 11.
[2]DNAm exp. SP = mean DNA methylation levels in individuals with exposure to adversity that occurred during the selected sensitive period (SP).
[3]ΔDNAm= difference in mean DNA methylation levels between individuals exposed to adversity during the selected sensitive period and individuals unexposed to adversity (i.e., DNAm exp. SP – DNAm unexp.)
[4]Effect estimates were calculated using linear regression of exposure to adversity from the theoretical model and DNA methylation, correcting for the covariates described in the methods.
* SE = standard error; bolded loci passed a 5% FDR threshold in the original age 15 analysis.

**Table S8. Associations between adversity and DNA methylation at age 7 (whole blood) for loci identified at age 15.**

| Adversity | Timing | Age (years) | CpG | DNAm unexposed[1] | DNAm exp. SP[2] | Δ DNAm[3] | Effect estimate[4] | SE* | P-value | FDR |
|---|---|---|---|---|---|---|---|---|---|---|
| Caregiver physical or emotional abuse | Early childhood | 5 | cg14855874 | 0.089 | 0.102 | 0.013 | 0.012 | 0.006 | 3.06E-02 | 2.51E-01 |
| | | | cg15454534 | 0.888 | 0.889 | 0.001 | 0.001 | 0.003 | 6.51E-01 | 9.60E-01 |
| | | | cg06215562 | 0.839 | 0.843 | 0.004 | 0.004 | 0.005 | 4.58E-01 | 9.60E-01 |
| Sexual or physical abuse (by anyone) | Early childhood | 3.5 | **cg26970800** | **0.902** | **0.887** | **-0.015** | **-0.015** | **0.010** | **1.27E-01** | **6.51E-01** |
| | | | **cg15723468** | **0.799** | **0.807** | **0.008** | **0.006** | **0.009** | **4.63E-01** | **9.60E-01** |
| | | | **cg17928317** | **0.695** | **0.726** | **0.031** | **-0.002** | **0.016** | **8.97E-01** | **9.60E-01** |
| | Late childhood | 8 | cg27558057 | 0.248 | 0.224 | -0.024 | 0.068 | 0.021 | 1.63E-03 | 6.67E-02 |
| Family instability | Very early childhood | 2.5 | cg02735620 | 0.877 | 0.880 | 0.002 | 0.003 | 0.004 | 4.72E-01 | 9.60E-01 |
| Financial hardship | Very early childhood | 0.66 | cg14455319 | 0.266 | 0.288 | 0.021 | 0.022 | 0.009 | 1.43E-02 | 2.27E-01 |
| | | | cg13204236 | 0.867 | 0.868 | 0.001 | 0.002 | 0.006 | 7.44E-01 | 9.60E-01 |
| | Early childhood | 5 | cg15037420 | 0.795 | 0.792 | -0.003 | -0.003 | 0.007 | 7.06E-01 | 9.60E-01 |
| | | | cg06410970 | 0.870 | 0.868 | -0.003 | -0.002 | 0.006 | 6.89E-01 | 9.60E-01 |
| | Late childhood | 11 | cg02011706 | 0.860 | 0.863 | 0.003 | 0.006 | 0.012 | 6.05E-01 | 9.60E-01 |
| | | | cg04659536 | 0.906 | 0.905 | -0.001 | -0.002 | 0.005 | 7.38E-01 | 9.60E-01 |
| | Recency | | cg17670999 | 0.836 | 0.836 | 0.000 | 0.000 | 0.000 | 4.15E-01 | 9.60E-01 |
| | | | cg25459301 | 0.791 | 0.788 | -0.002 | 0.000 | 0.000 | 6.66E-01 | 9.60E-01 |
| | | | cg06812747 | 0.847 | 0.843 | -0.004 | 0.000 | 0.000 | 8.49E-01 | 9.60E-01 |
| Maternal psychopathology | Very early childhood | 2.75 | **cg16813552** | **0.890** | **0.882** | **-0.008** | **-0.007** | **0.003** | **2.47E-02** | **2.51E-01** |
| Neighborhood disadvantage | Very early childhood | 2.75 | cg04288299 | 0.932 | 0.935 | 0.003 | 0.006 | 0.003 | 7.41E-02 | 5.06E-01 |
| | | | cg25019631 | 0.194 | 0.173 | -0.021 | -0.013 | 0.009 | 1.46E-01 | 6.66E-01 |
| | | | cg04224851 | 0.903 | 0.915 | 0.012 | 0.006 | 0.003 | 1.66E-02 | 2.27E-01 |
| One adult in the household | Very early childhood | 1.75 | **cg05491478** | **0.915** | **0.920** | **0.006** | **0.005** | **0.005** | **2.94E-01** | **9.60E-01** |
| | Early childhood | 3.9 | **cg16907527** | **0.844** | **0.845** | **0.001** | **0.001** | **0.005** | **7.86E-01** | **9.60E-01** |
| | | | **cg08818094** | **0.858** | **0.851** | **-0.007** | **-0.005** | **0.007** | **4.68E-01** | **9.60E-01** |
| | | | **cg01060989** | **0.834** | **0.835** | **0.001** | **0.001** | **0.006** | **9.13E-01** | **9.60E-01** |
| | | | **cg15814750** | **0.752** | **0.747** | **-0.006** | **-0.005** | **0.008** | **4.81E-01** | **9.60E-01** |
| | | | **cg15783822** | **0.878** | **0.880** | **0.002** | **0.002** | **0.004** | **5.46E-01** | **9.60E-01** |
| | | | **cg15864691** | **0.911** | **0.913** | **0.002** | **0.002** | **0.003** | **4.11E-01** | **9.60E-01** |

| | | | | | | | | | |
|---|---|---|---|---|---|---|---|---|---|
| | | **cg02584161** | **0.688** | **0.690** | **0.002** | **0.000** | **0.013** | **9.88E-01** | **9.88E-01** |
| | | **cg02810291** | **0.833** | **0.836** | **0.003** | **0.003** | **0.005** | **5.66E-01** | **9.60E-01** |
| | | **cg04036644** | **0.903** | **0.903** | **0.000** | **-0.001** | **0.005** | **8.99E-01** | **9.60E-01** |
| | | **cg11811897** | **0.778** | **0.772** | **-0.006** | **-0.008** | **0.009** | **3.43E-01** | **9.60E-01** |
| | | **cg15817130** | **0.822** | **0.824** | **0.002** | **0.000** | **0.006** | **9.57E-01** | **9.80E-01** |
| | | **cg06711254** | **0.713** | **0.704** | **-0.008** | **-0.009** | **0.011** | **4.27E-01** | **9.60E-01** |
| | | **cg19096460** | **0.853** | **0.850** | **-0.003** | **-0.002** | **0.005** | **6.05E-01** | **9.60E-01** |
| | | cg18980650 | 0.795 | 0.788 | -0.007 | -0.002 | 0.007 | 7.62E-01 | 9.60E-01 |
| | | cg27504269 | 0.783 | 0.781 | -0.001 | -0.001 | 0.008 | 8.90E-01 | 9.60E-01 |
| Late childhood | 10 | **cg12096528** | **0.885** | **0.886** | **0.001** | **0.002** | **0.004** | **6.85E-01** | **9.60E-01** |
| Accumulation | | **cg00807464** | **0.050** | **0.051** | **0.001** | **0.001** | **0.001** | **1.12E-01** | **6.51E-01** |
| | | **cg10420609** | **0.603** | **0.602** | **-0.001** | **0.001** | **0.003** | **8.34E-01** | **9.60E-01** |
| | | **cg14579651** | **0.663** | **0.653** | **-0.010** | **-0.003** | **0.003** | **2.03E-01** | **8.30E-01** |

[1]DNAm unexp. = mean DNA methylation levels at age 7 in individuals with no exposure to adversity from age 0 to 11.
[2]DNAm exp. SP = mean DNA methylation levels at age 7 in individuals with exposure to adversity that occurred during the selected sensitive period (SP).
[3]ΔDNAm= difference in mean DNA methylation levels between individuals exposed to adversity during the selected sensitive period and individuals unexposed to adversity (i.e., DNAm exp. SP – DNAm unexp.)
[4]Effect estimates were calculated using linear regression of exposure to adversity from the theoretical model and DNA methylation, correcting for the covariates described in the methods.
* SE = standard error; bolded loci passed a 5% FDR threshold in the original age 15 analysis.

**Table S9. Types of longitudinal DNAm trajectories in response to childhood adversity for top adolescent loci.**

| Adversity | Timing | Age (years) | CpG | Trajectory name |
|---|---|---|---|---|
| Caregiver physical or emotional abuse | Early childhood | 5 | cg14855874 | **Emergent** |
| | | | cg15454534 | **Latent** |
| | | | cg06215562 | **Latent** |
| Sexual or physical abuse (by anyone) | Early childhood | 3.5 | **cg26970800** | **Emergent** |
| | | | **cg15723468** | **Latent** |
| | | | **cg17928317** | **Primed** |
| | Late childhood | 8 | cg27558057 | **Stable** |
| Family instability | Very early childhood | 2.5 | cg02735620 | Emergent |
| Financial hardship | Very early childhood | 0.66 | cg14455319 | Time-stable |
| | | | cg13204236 | Latent |
| | Early childhood | 5 | cg15037420 | Latent |
| | | | cg06410970 | Overcompensation |
| | Late childhood | 11 | cg02011706 | Emergent |
| | | | cg04659536 | Latent |
| | Recency | | cg17670999 | Stable |
| | | | cg25459301 | Overcompensation |
| | | | cg06812747 | Stable |
| Maternal psychopathology | Very early childhood | 2.75 | **cg16813552** | **Stable** |
| Neighborhood disadvantage | Very early childhood | 2.75 | cg04288299 | **Overcompensation** |
| | | | cg25019631 | **Overcompensation** |
| | | | cg04224851 | **Overcompensation** |
| One adult in the household | Very early childhood | 1.75 | **cg05491478** | **Overcompensation** |
| | Early childhood | 3.9 | **cg16907527** | **Flat emergent** |
| | | | **cg08818094** | **Latent** |
| | | | **cg01060989** | **Latent** |
| | | | **cg15814750** | **Latent** |
| | | | **cg15783822** | **Latent** |
| | | | **cg15864691** | **Overcompensation** |
| | | | **cg02584161** | **Latent** |
| | | | **cg02810291** | **Overcompensation** |

| | | | |
|---|---|---|---|
| | | **cg04036644** | **Latent** |
| | | **cg11811897** | **Latent** |
| | | **cg15817130** | **Latent** |
| | | **cg06711254** | **Flat emergent** |
| | | **cg19096460** | **Latent** |
| | | cg18980650 | Emergent |
| | | cg27504269 | Latent |
| Late childhood | 10 | **cg12096528** | **Overcompensation** |
| Accumulation | | **cg00807464** | **Stable** |
| | | **cg10420609** | **Latent** |
| | | **cg14579651** | **Stable** |

*Bolded loci passed a 5% FDR threshold in the original analysis.

**Table S10. Persistence of differential DNA methylation patterns identified at age 7 into adolescence (age 15).**

| Adversity | Timing | Age (years) | CpG | DNAm unexposed[1] | DNAm exp. SP[2] | ΔDNAm[3] | Effect estimate[4] | SE* | P-value | FDR |
|---|---|---|---|---|---|---|---|---|---|---|
| Caregiver physical or emotional abuse | Middle childhood | 6 | cg12023170 | 0.098 | 0.105 | 0.008 | 0.006 | 0.007 | 4.02E-01 | 8.56E-01 |
| Sexual or physical abuse (by anyone) | Early childhood | 4.75 | cg20369299 | 0.682 | 0.662 | -0.02 | -0.016 | 0.018 | 3.72E-01 | 8.56E-01 |
| | | | cg13817046 | 0.425 | 0.424 | -0.001 | 0.001 | 0.014 | 9.18E-01 | 9.61E-01 |
| Family instability | Very early childhood | 2.5 | cg04079399 | 0.885 | 0.883 | -0.002 | -0.002 | 0.004 | 5.90E-01 | 8.75E-01 |
| | Early childhood | 4.75 | cg01407460 | 0.023 | 0.024 | 0.000 | 0.001 | 0.001 | 4.22E-01 | 8.56E-01 |
| | | | cg17134302 | 0.835 | 0.836 | 0.001 | 0.001 | 0.006 | 8.63E-01 | 9.61E-01 |
| | | | cg13706680 | 0.875 | 0.883 | 0.008 | 0.008 | 0.005 | 6.30E-02 | 7.16E-01 |
| | | | cg27457457 | 0.664 | 0.646 | -0.017 | -0.015 | 0.016 | 3.74E-01 | 8.56E-01 |
| | | | cg01504589 | 0.836 | 0.828 | -0.008 | -0.007 | 0.009 | 4.28E-01 | 8.56E-01 |
| | | | cg13876553 | 0.801 | 0.805 | 0.004 | 0.006 | 0.009 | 5.43E-01 | 8.75E-01 |
| | | | cg01841772 | 0.810 | 0.825 | 0.014 | 0.014 | 0.009 | 1.02E-01 | 7.16E-01 |
| | | | cg16231917 | 0.214 | 0.242 | 0.028 | 0.025 | 0.015 | 8.66E-02 | 7.16E-01 |
| | | | cg26997966 | 0.860 | 0.854 | -0.006 | -0.007 | 0.005 | 2.24E-01 | 8.56E-01 |
| | | | cg14401897 | 0.799 | 0.808 | 0.009 | 0.010 | 0.010 | 3.37E-01 | 8.56E-01 |
| | | | cg27639644 | 0.854 | 0.851 | -0.003 | -0.003 | 0.006 | 6.78E-01 | 8.75E-01 |
| | | | cg02886132 | 0.878 | 0.885 | 0.007 | 0.007 | 0.004 | 9.28E-02 | 7.16E-01 |
| | | | cg27061903 | 0.051 | 0.054 | 0.003 | 0.003 | 0.003 | 2.84E-01 | 8.56E-01 |
| | | | cg10571837 | 0.897 | 0.903 | 0.006 | 0.006 | 0.004 | 1.15E-01 | 7.16E-01 |
| | | | cg12188526 | 0.883 | 0.885 | 0.001 | 0.002 | 0.004 | 6.95E-01 | 8.75E-01 |
| | | | cg21172807 | 0.109 | 0.124 | 0.014 | 0.014 | 0.005 | 9.90E-03 | 4.55E-01 |
| | | | cg01267076 | 0.846 | 0.847 | 0.002 | 0.003 | 0.007 | 6.36E-01 | 8.75E-01 |
| | | | cg22346081 | 0.858 | 0.860 | 0.002 | 0.002 | 0.005 | 6.40E-01 | 8.75E-01 |
| | | | cg16338178 | 0.825 | 0.821 | -0.004 | -0.003 | 0.007 | 6.75E-01 | 8.75E-01 |
| | | | cg08971940 | 0.772 | 0.785 | 0.013 | 0.014 | 0.011 | 1.97E-01 | 8.56E-01 |
| | | | cg14948379 | 0.851 | 0.848 | -0.003 | -0.003 | 0.007 | 6.79E-01 | 8.75E-01 |

| | | | | | | | | | |
|---|---|---|---|---|---|---|---|---|---|
| | | cg01654242 | 0.810 | 0.817 | 0.007 | 0.007 | 0.010 | 4.88E-01 | 8.75E-01 |
| | | cg11438065 | 0.901 | 0.902 | 0.002 | 0.002 | 0.004 | 6.12E-01 | 8.75E-01 |
| | | cg22011436 | 0.840 | 0.846 | 0.006 | 0.007 | 0.008 | 3.75E-01 | 8.56E-01 |
| | | cg01587190 | 0.058 | 0.061 | 0.003 | 0.003 | 0.002 | 7.05E-02 | 7.16E-01 |
| | | cg01023798 | 0.854 | 0.853 | -0.002 | 0.000 | 0.006 | 9.92E-01 | 9.92E-01 |
| | | cg09305491 | 0.910 | 0.909 | -0.001 | -0.001 | 0.004 | 7.80E-01 | 9.38E-01 |
| | | cg22060367 | 0.880 | 0.880 | 0.000 | 0.000 | 0.005 | 9.20E-01 | 9.61E-01 |
| | | cg05353659 | 0.892 | 0.888 | -0.004 | -0.004 | 0.004 | 3.41E-01 | 8.56E-01 |
| | | cg27567416 | 0.882 | 0.887 | 0.005 | 0.006 | 0.004 | 1.42E-01 | 7.24E-01 |
| | | cg07206497 | 0.876 | 0.876 | 0.001 | 0.002 | 0.005 | 7.04E-01 | 8.75E-01 |
| | | cg05886789 | 0.839 | 0.841 | 0.002 | 0.003 | 0.006 | 5.77E-01 | 8.75E-01 |
| | | cg14637285 | 0.858 | 0.851 | -0.007 | -0.007 | 0.006 | 2.23E-01 | 8.56E-01 |
| | | cg00967695 | 0.883 | 0.875 | -0.008 | -0.008 | 0.007 | 2.62E-01 | 8.56E-01 |
| | | cg01100868 | 0.892 | 0.894 | 0.002 | 0.003 | 0.004 | 5.20E-01 | 8.75E-01 |
| | | cg23184756 | 0.834 | 0.835 | 0.001 | 0.000 | 0.006 | 9.41E-01 | 9.61E-01 |
| | | cg00943585 | 0.828 | 0.824 | -0.005 | -0.003 | 0.011 | 8.16E-01 | 9.38E-01 |
| Middle childhood | 5.75 | cg17719337 | 0.040 | 0.040 | 0.000 | 0.000 | 0.002 | 9.39E-01 | 9.61E-01 |
| | | cg26848593 | 0.027 | 0.028 | 0.001 | 0.000 | 0.001 | 6.23E-01 | 8.75E-01 |
| | | cg06770536 | 0.733 | 0.718 | -0.015 | -0.018 | 0.012 | 1.24E-01 | 7.16E-01 |
| | 6.75 | cg19569074 | 0.677 | 0.668 | -0.009 | -0.004 | 0.016 | 7.98E-01 | 9.38E-01 |
| | | cg10940545 | 0.807 | 0.796 | -0.011 | -0.015 | 0.015 | 3.14E-01 | 8.56E-01 |

[1]DNAm unexp. = mean DNA methylation levels at age 15 in individuals with no exposure to adversity from age 0 to 11.
[2]DNAm exp. SP = mean DNA methylation levels at age 15 in individuals with exposure to adversity that occurred during the selected sensitive period (SP).
[3]ΔDNAm= difference in mean DNA methylation levels between individuals exposed to adversity during the selected sensitive period and individuals unexposed to adversity (i.e., DNAm exp. SP – DNAm unexp.)
[4]Effect estimates were calculated using linear regression of exposure to adversity during the selected sensitive period from the theoretical model and DNA methylation, correcting for the covariates described in the methods.
* SE = standard error.

### SUPPLEMENTAL FIGURES

**Figure S1. Summary of prevalence and correlations between adversities from age 0-11.**

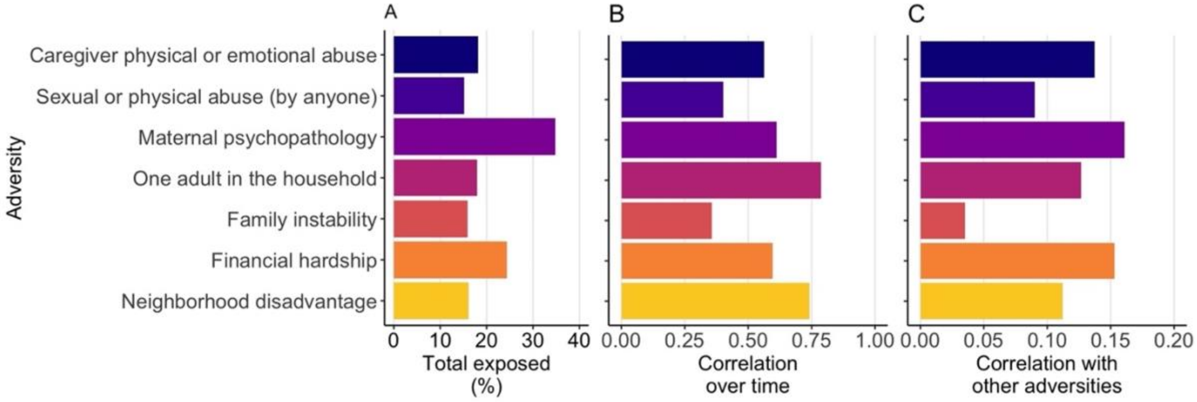

**A)** The prevalence of each adversity from age 0-11 varied by type, ranging from 15.1% (sexual or physical abuse by anyone) to 34.8% (maternal psychopathology).
**B)** Exposures within each type of adversity were generally correlated over time, ranging from 0.357 (family instability) to 0.786 (one adult in the household). Closer timepoints tended to be more related than more distant timepoints.
**C)** On average, the absolute correlation of exposures to different adversities was modest, ranging from -0.035 (family instability; shown here on absolute scale) to 0.161 (maternal psychopathology), which may reflect various dimensions of childhood adversity.

**Figure S2. Genomic locations of top age 15 loci compared to all sites tested (n=302,581).**

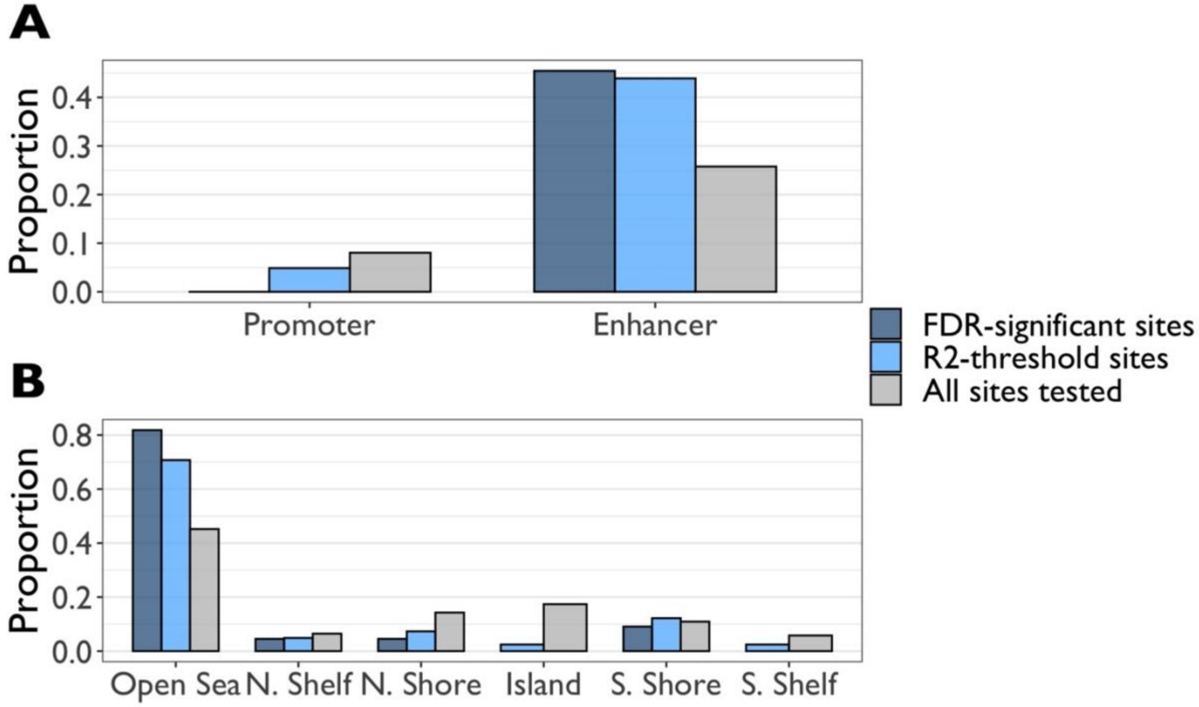

**A)** Compared to all tested sites, FDR-significant loci showed more enrichment in enhancer regions ($\chi^2$=4.5; p=0.034) and no presence in promoter regions ($\chi^2$=1.9; p=0.17). $R^2$-threshold loci also showed higher enrichment in enhancers ($\chi^2$=7.1; p=0.0079) and no differences in promoter enrichment ($\chi^2$=0.55; p=0.46).

**B)** FDR-significant loci also differed in terms of their location relation to CpG islands, showing higher enrichment in Open Sea regions and decreased enrichment in CpG islands compared to all sites ($\chi^2$=13.3; p=0.021). $R^2$-threshold loci also higher enrichment in Open Sea regions and decreased enrichment in CpG islands compared to all sites ($\chi^2$=13.6; p=0.018).

**Figure S3. Brain-blood correlations for top loci identified at age 15.**

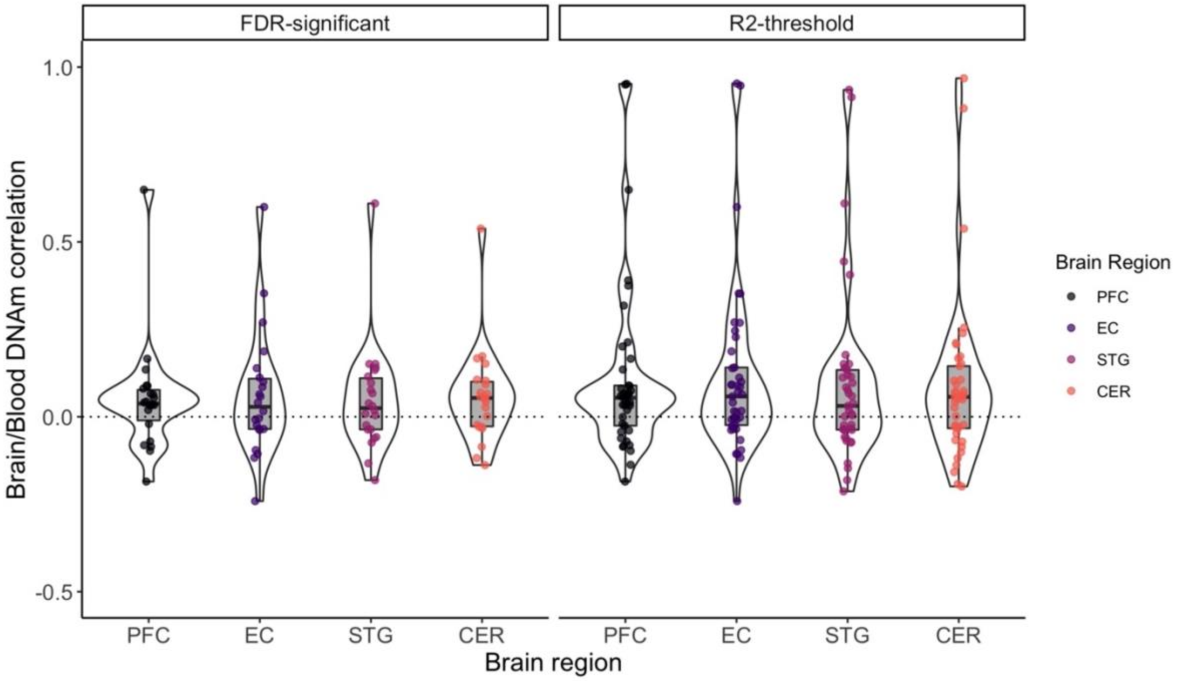

Correlations between DNA methylation measured in blood and specific brain regions are shown for the 22 FDR-significant loci identified at age 15, as well as the 41 loci that passed an $R^2$ threshold of 0.035. Data were obtained from Hannon et al., 2015. PFC = prefrontal cortex; EC = entorhinal cortex; STG = superior temporal gyrus; CER = cerebellum.

**Figure S4. Enrichment of Gene Ontology (GO) term clusters for top loci at age 15.**

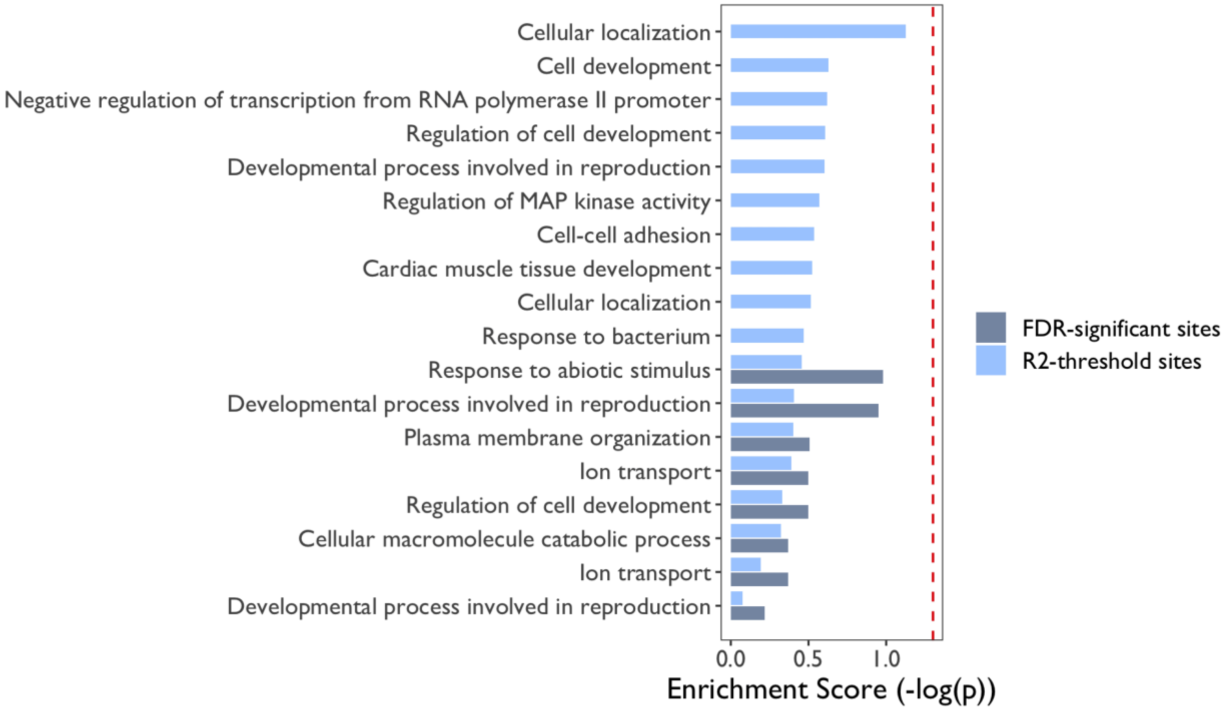

The 22 FDR-significant loci were annotated to 21 unique genes, while the 41 $R^2$-threshold loci were annotated to 39 genes. The plot shows the clusters of GO biological processes that emerged from these genes, as analyzed using DAVID (4,5). No clusters were significant at p<0.05, shown here as the dotted red line corresponding to an enrichment score of 1.3.

**Figure S5. Genes annotated to top age 15 loci were no more highly constrained than all sites.**

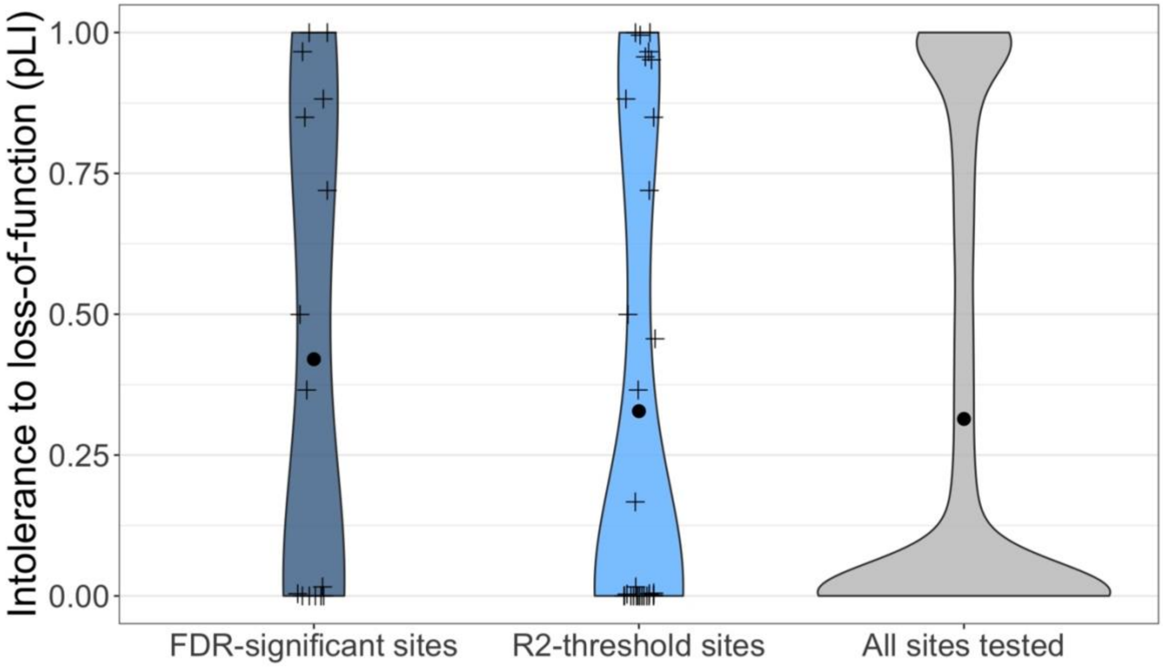

Violin plots show the distribution of gene constraint scores (pLI) for FDR-significant (n=17 annotated genes from 22 loci), $R^2$-threshold loci (n=33 annotated genes from 41 loci), and genome-wide loci (n=16,114), where higher values represent increased probability of a gene being intolerant to Loss-of-Function variation. Genes annotated to FDR-significant sites were no more highly constrained than the rest of genes tested (permutation p=0.27 for FDR-significant subset; p=0.51 for $R^2$-threshold subset). Black points represent mean pLI values for the two sets of genes. Three genes in the set of FDR-significant loci showed a pLI>0.9 (*DSP*, *CUX2*, and *STK38L*), with four more in the $R^2$-threshold subset (*FBXL16*, *PKD2*, *TAF1*, and *XKR6*).

**Figure S6. Non-parametric bootstrapping of associations between childhood adversity and DNA methylation at age 15.**

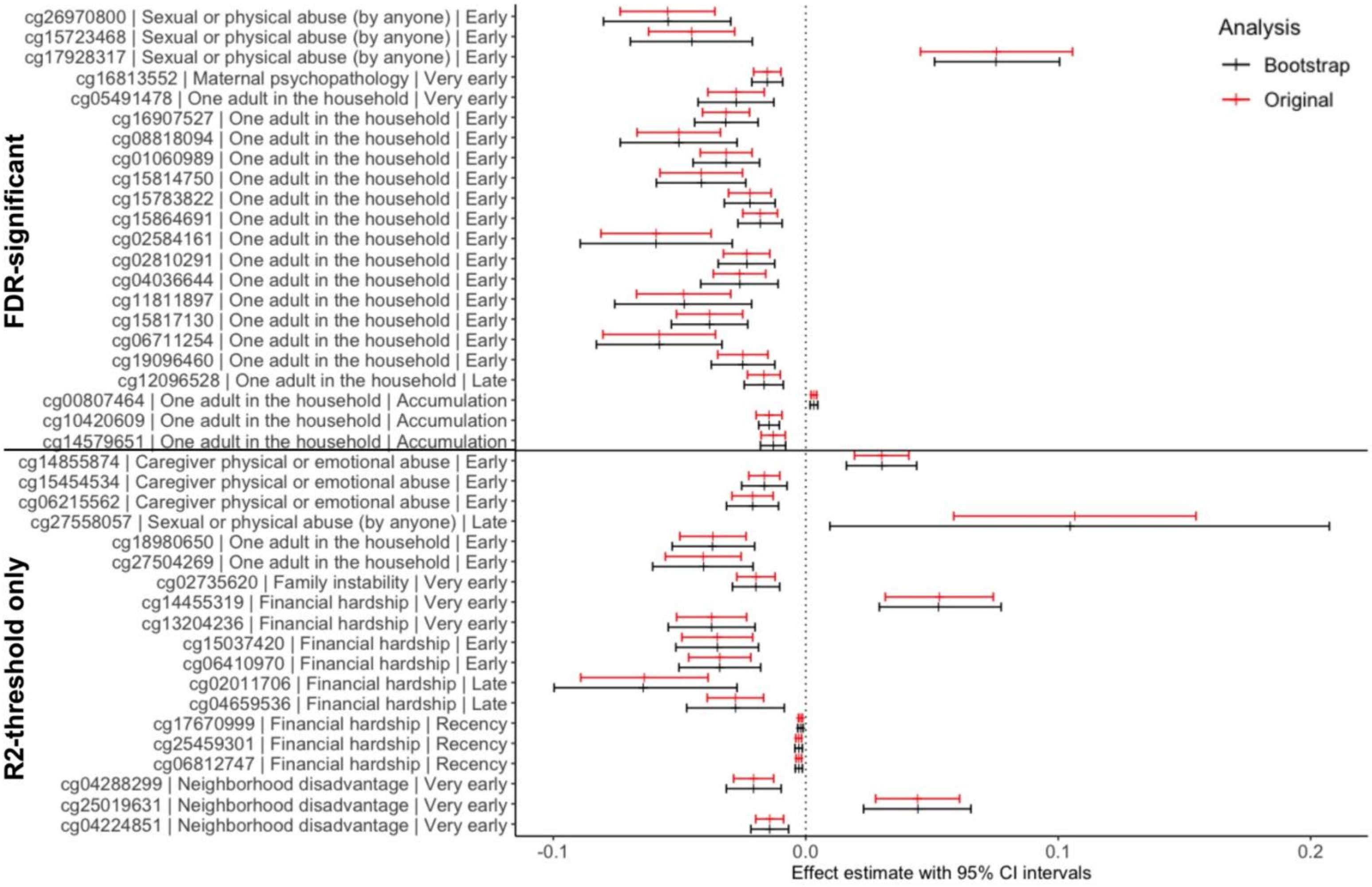

The 41 $R^2$-threshold associations (of which 22 passed a 5% FDR cutoff) between childhood adversity and DNA methylation at age 15 were internally validated using non-parametric bootstrap analyses. The average effect estimates for the 10,000 bootstraps (black) showed only minor differences from the effects estimates generated in the original analyses of childhood adversity and DNAm (red). 95% confidence intervals are shown.

**Figure S7. Significance levels of associations between childhood adversity and DNA methylation at age 15 for mutually-adjusted regression models.**

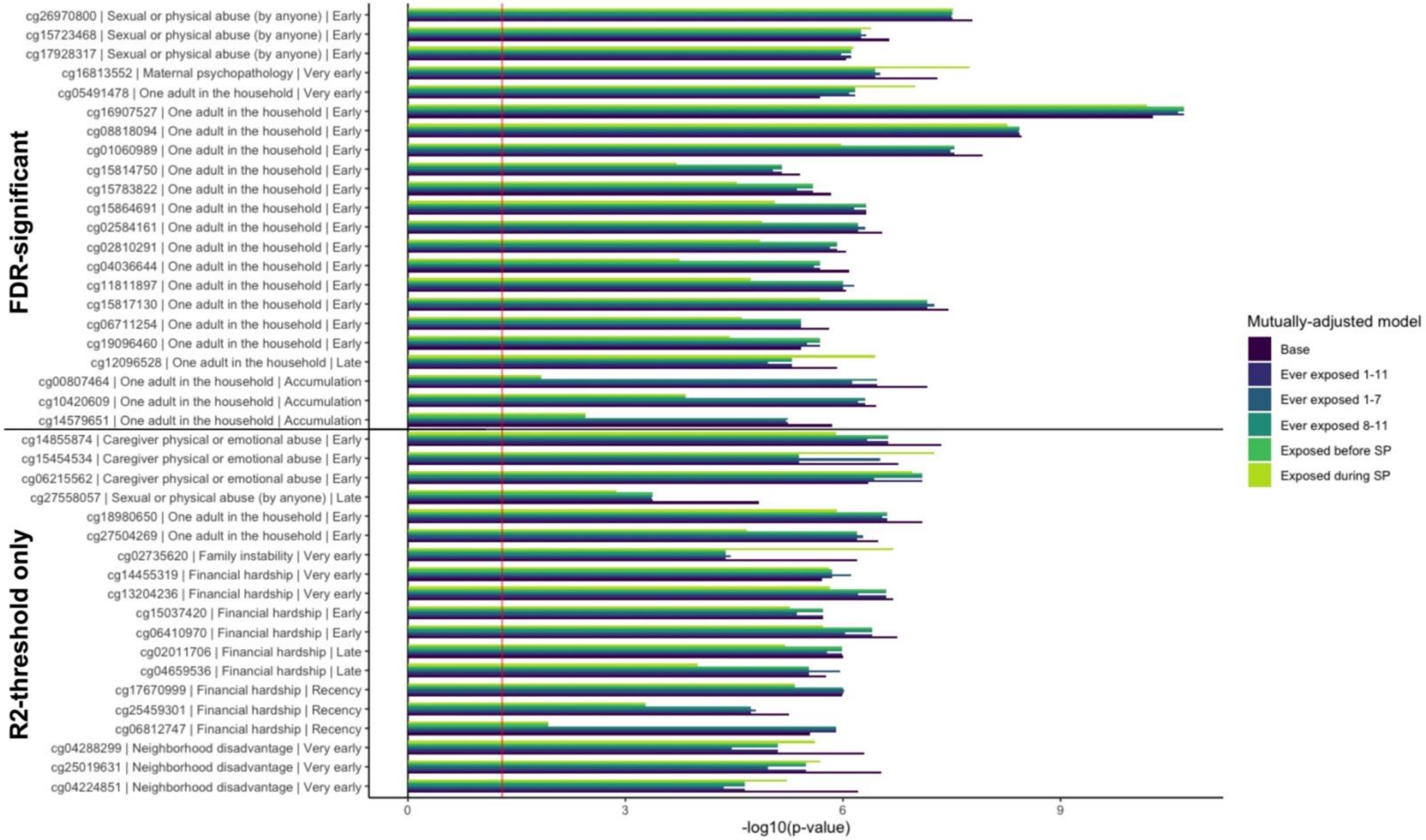

We compared the significance of associations between childhood adversity and DNA methylation (DNAm) at age 15 between the base model and "mutually-adjusted" models, which additionally included other types of childhood adversity. These five mutually-adjusted models included a variable of exposure to any other adversity between age 1-11, age 1-7, or age 8-11. We also tested the effects of exposures to adversity before or during the SLCMA-selected sensitive period; accumulation hypotheses were corrected using the total number of exposures from age 1-11. Significance levels are represented by the $-\log_{10}$ of p-values, whereby larger values represent smaller p-values (higher significance) and smaller values represent larger p-values (lower significance). The red line shows the $-\log_{10}$ of p=0.05. All associated passed a false-discovery rate of 0.05 when correcting for the testing of 22 FDR-significant loci.

**Figure S8. Change in effects estimates for mutually-adjusted regression models of adversity and DNA methylation at age 15.**

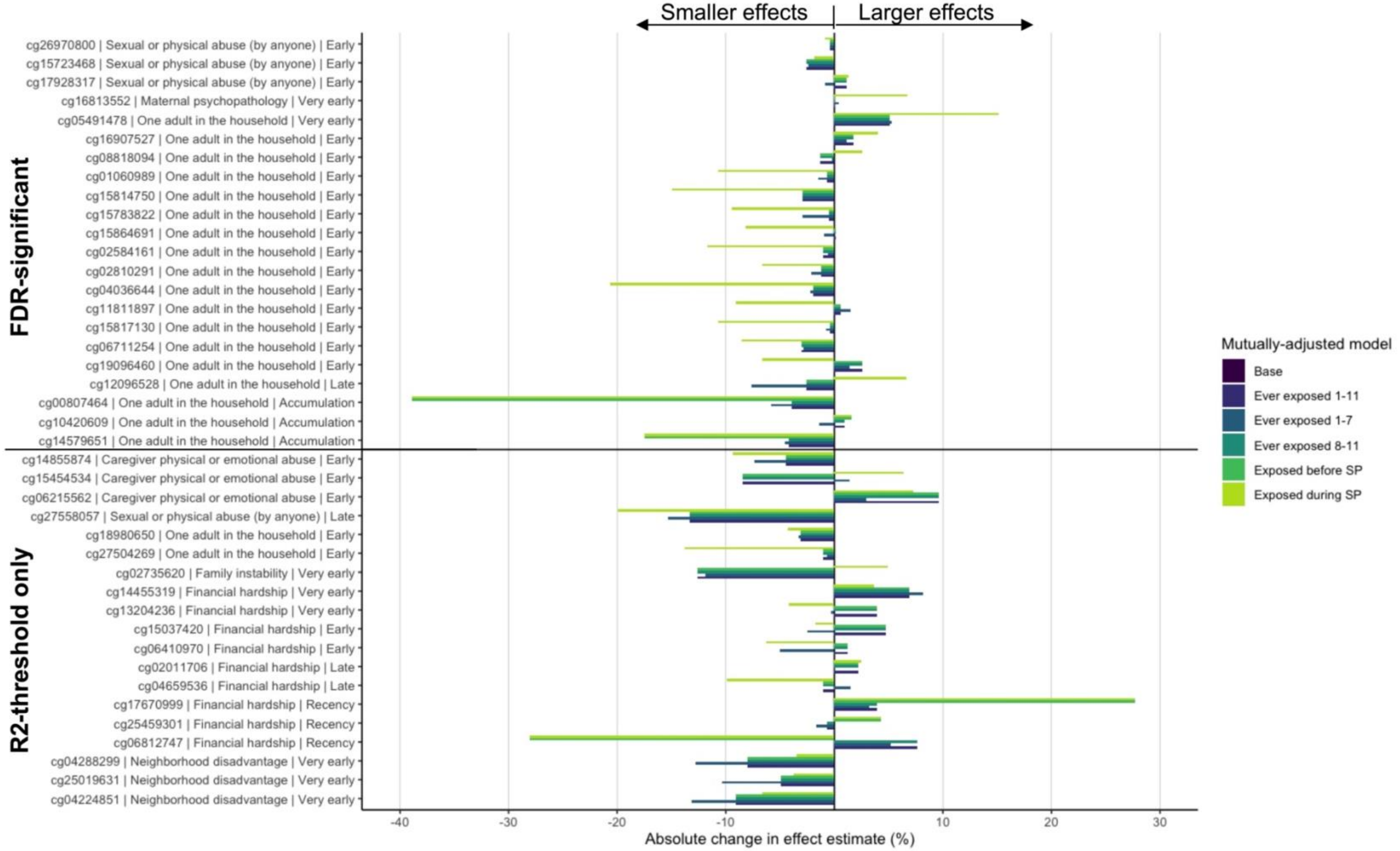

The strength of associations between childhood adversity and DNA methylation (DNAm) at age 15 from the base model were compared to mutually-adjusted models, which additionally included other types of childhood adversity. These five “mutually-adjusted” models included a variable of exposure to any other adversity between age 1-11, age 1-7, or age 8-11. We also tested the effects of exposures to adversity before or during the SLCMA-selected sensitive period (SP); accumulation hypotheses were corrected using the total number of exposures from age 1-11. The majority of associations showed little change in the strength of associations between a given childhood adversity and DNAm when accounting for other exposures, shown as the absolute percent change in effect estimate. However, associations between the accumulation of exposures to one-adult households and DNAm at age 15 were most attenuated in the mutually-adjusted models, showing a 1-40% reduction in the size of the effect estimate. Accounting for exposure that co-occurred during the SLCMA-selected sensitive period also resulted in smaller effect estimates for exposures to one-adult households during early childhood.

**Figure S9. Average differences across mutually-adjusted models of exposure to childhood adversity and DNA methylation at age 15.**

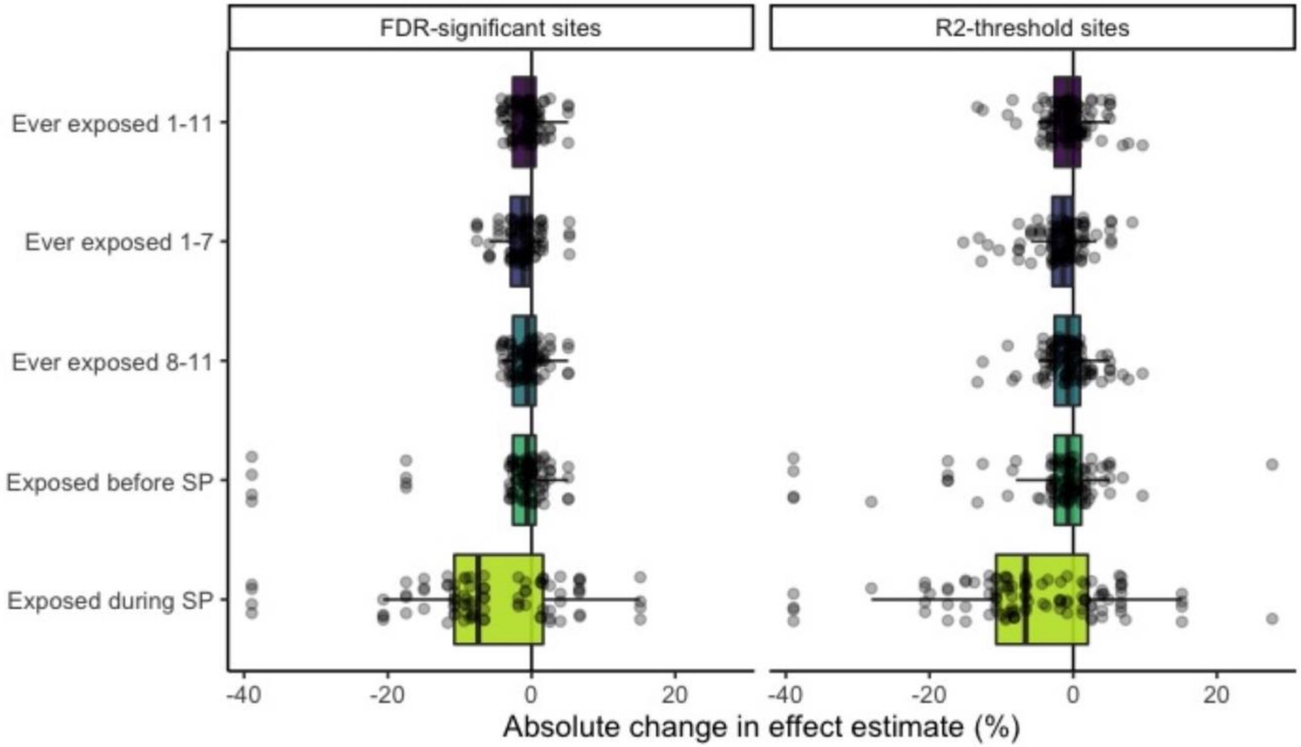

The strength of the associations between childhood adversity and DNA methylation (DNAm) at age 15 from the base model were compared to mutually-adjusted models that accounted for the potential effects of other types of childhood adversity. These "mutually-adjusted" models included a variable of exposure to any other adversity between age 1-11, age 1-7, or age 8-11. We also tested the effects of exposures to adversity before or during the SLCMA-selected sensitive period (SP); accumulation hypotheses were corrected using the total number of exposures from age 1-11. Across all 22 loci FDR-significant, the effects of mutual adjustment were most pronounced when correcting for exposures that occurred during the same sensitive period (mean = -6.3%, range = -38.9% to 15.1%). These effects were similar in the 41 $R^2$-treshold loci (mean = -4.7%, range = -38.9% to 27.7%).

**Figure S10. Approaches to account for potential confounders.**

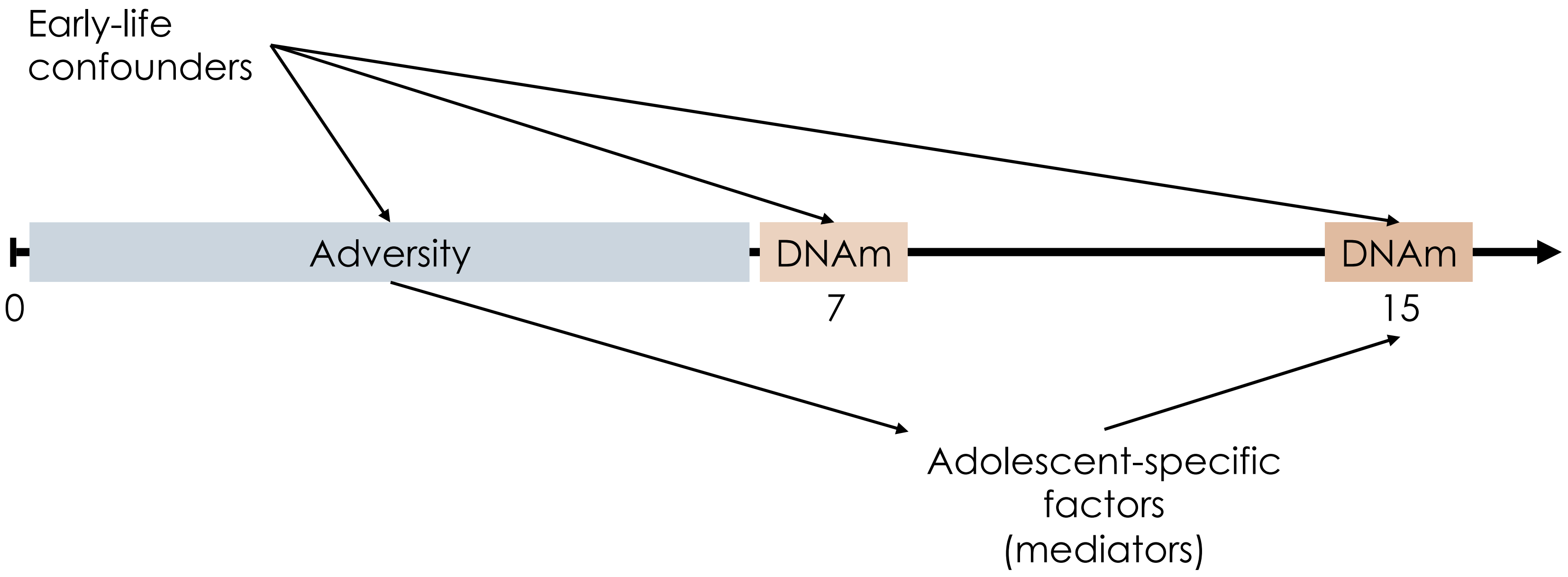

We identified two main types of confounders that may have influenced the results of our analyses between time-varying childhood adversity and DNA methylation (DNAm) patterns at age 7 and 15.
First, early-life confounders could have influenced the results of analyses of both age 7 and age 15 DNA methylation levels. These early-life confounders were investigated by including or removing covariates from the regression analyses of the 23 adolescent-specific loci to determine whether they influenced the strength of associations.
Second, adolescent-specific factors, meaning those that occurred after age 7, could only influence associations with age 15 DNA methylation for temporal reasons. Given that confounders must be associated with the exposure (adversity) and outcome (DNAm at age 15), adolescent-specific confounders were considered as potential mediators of this relationship. In this case, any factors that significantly mediated this relationship would explain why associations between adversity and DNAm were not present at age 7.

**Figure S11. Effects of early-life confounders on strength of associations between time-varying adversity and DNA methylation at age 15.**

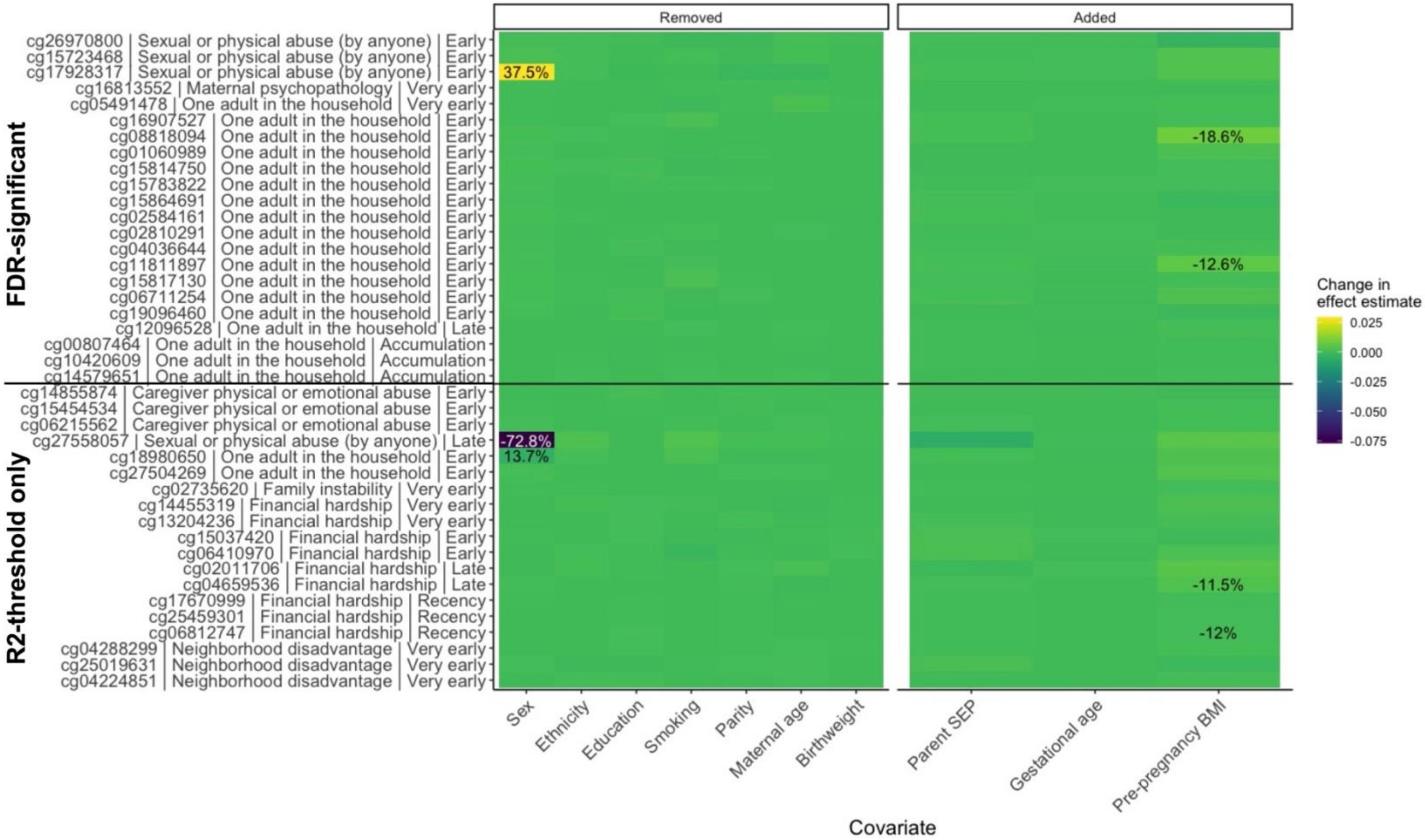

We investigated the impact of removing or adding confounders to our regression analyses of the CpGs that showed associations between childhood adversity and age 15 DNA methylation, compared to a base model that included the following covariates: sex, ethnicity, maternal education at birth, maternal smoking during pregnancy (smoking), parity, maternal age at birth, and birthweight. Removing potential confounders from our analyses resulted in small changes to the effect estimate from the regression model, except for two CpGs on chromosome X (cg17928317; cg27558057), which showed large changes in effect when sex was not included in the model. When adding potential confounders to the regression model, we again found small changes in effect estimates, with only four CpGs showing a >10% change in effect upon including of maternal pre-pregnancy body mass index (BMI). Parental socio-economic status at birth (SES parent) and gestational age in weeks did not influence the strength of associations.

**Figure S12. Effects of early-life confounders on strength of associations between time-varying adversity and DNA methylation at age 7.**

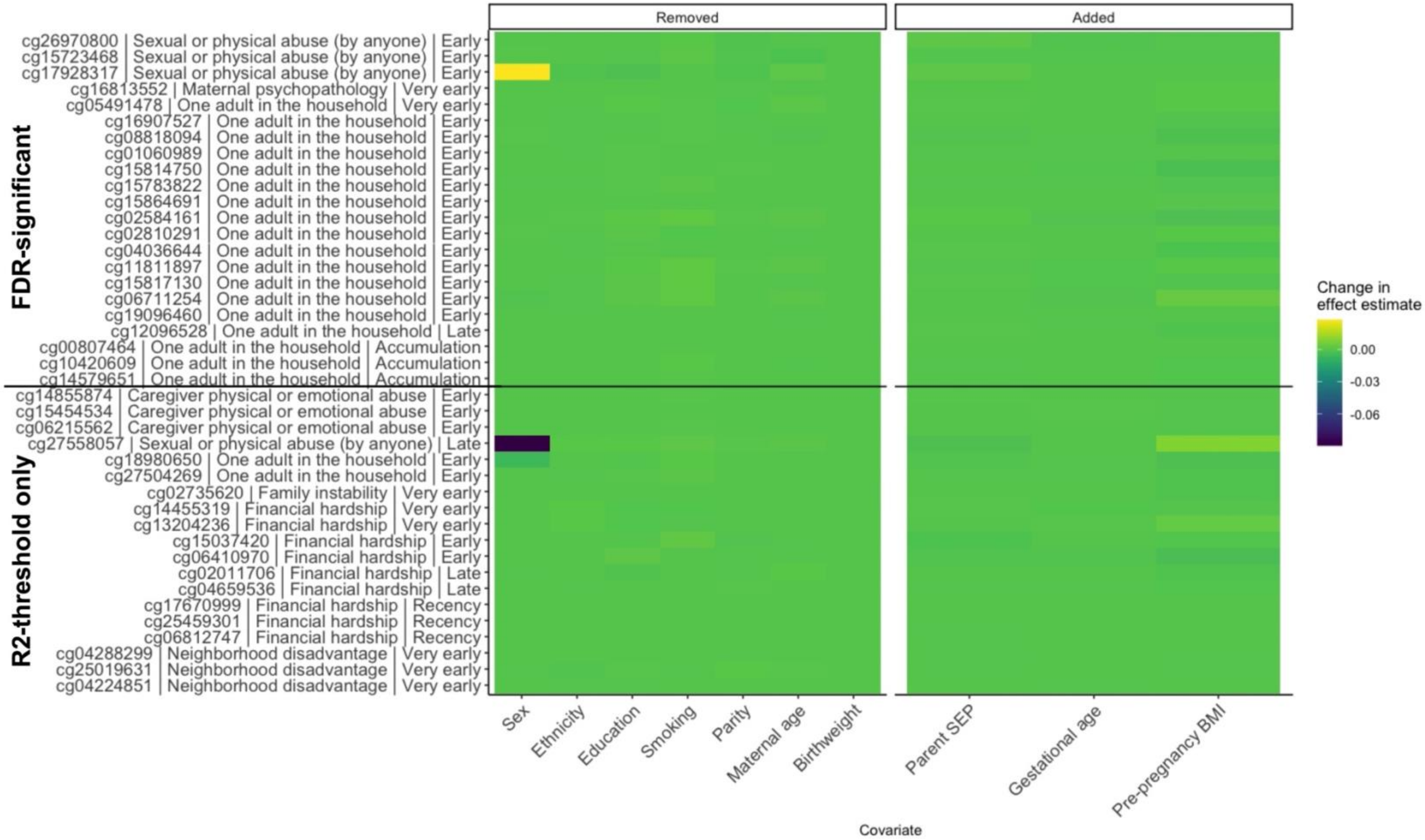

We investigated the impact of removing or adding confounders to our regression analyses of the 23 CpGs that showed associations between childhood adversity and age 15 DNA methylation, compared to a base model that included the following covariates: sex, ethnicity, maternal education at birth, maternal smoking during pregnancy (smoking), parity, maternal age at birth, and birthweight. With this base model, none of the loci showed significant associations between childhood adversity and DNA methylation at age 7.

Removing potential confounders from our analyses resulted in small changes to the effect estimate from the regression model, except for two CpGs on chromosome X (cg17928317; cg27558057), which showed a larger change in effect when sex was not included in the model. However, there were no changes in the significance level of these associations, as the effect estimates remained very small. When adding parental socio-economic status at birth (SES parent), gestational age in weeks, or maternal pre-pregnancy body mass index (BMI) to the base model, we again found minor fluctuations in the strength of associations, suggestive of little confounding effects on these associations.

**Figure S13. Age at pubertal onset did not mediate the effects of childhood adversity on age 15 DNA methylation.**

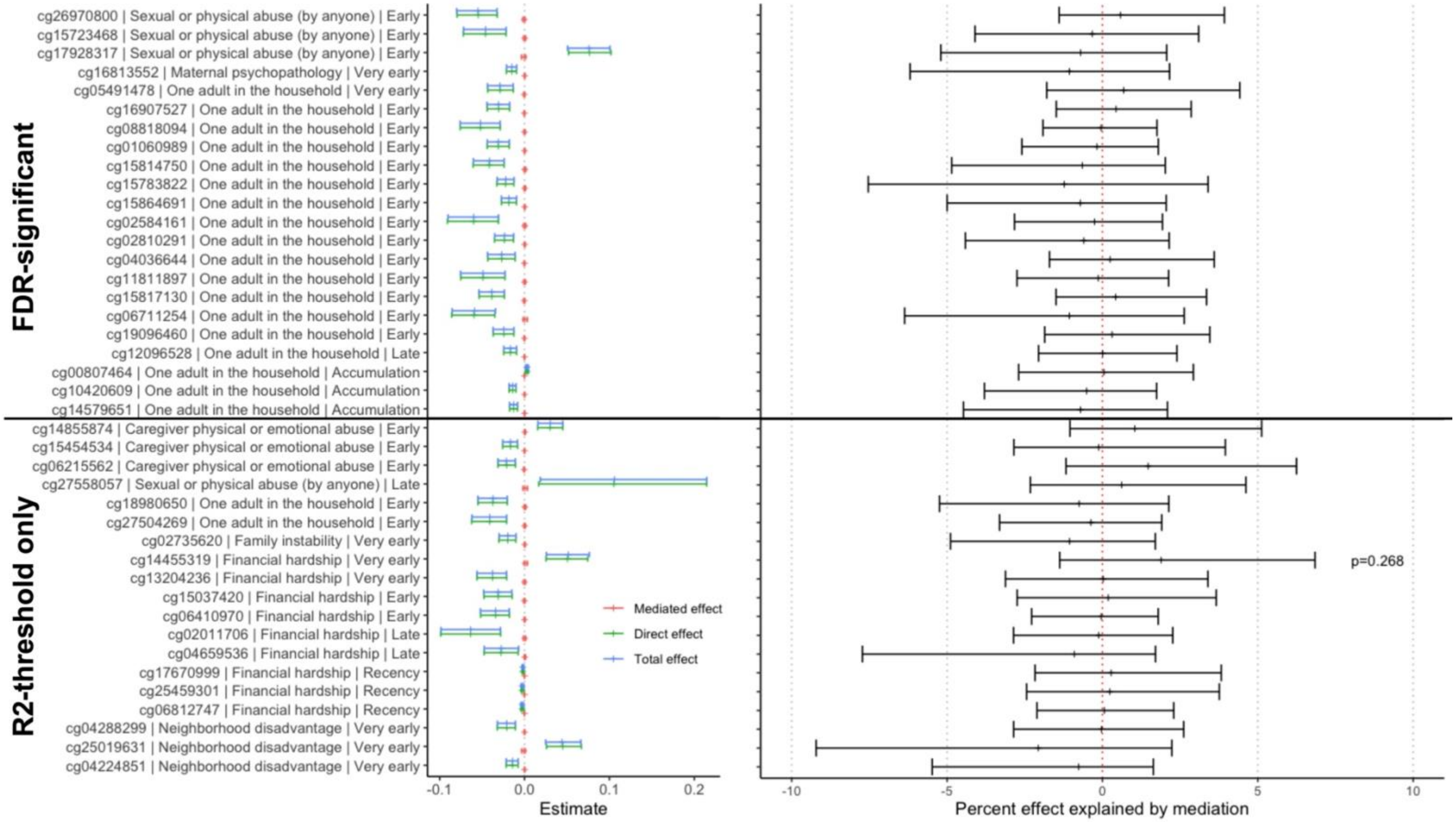

Mediation by the age of pubertal onset, estimated using peak height velocity, was tested for the loci associated with childhood adversity and DNA methylation at age 15. The average causal mediation effect (mediated effect, red; left panel) was close to zero for all CpGs, explaining very little of the association between childhood adversity and DNA methylation levels. None of the estimated mediated effects were significant ($p>0.05$). The lowest p-value belong to cg14455319 ($p=0.268$). Y-axis is noted as “CpG | childhood adversity | SLCMA hypothesis”.

**Figure S14. Body mass index at age 15 putatively mediated the effects of childhood adversity on age 15 DNA methylation.**

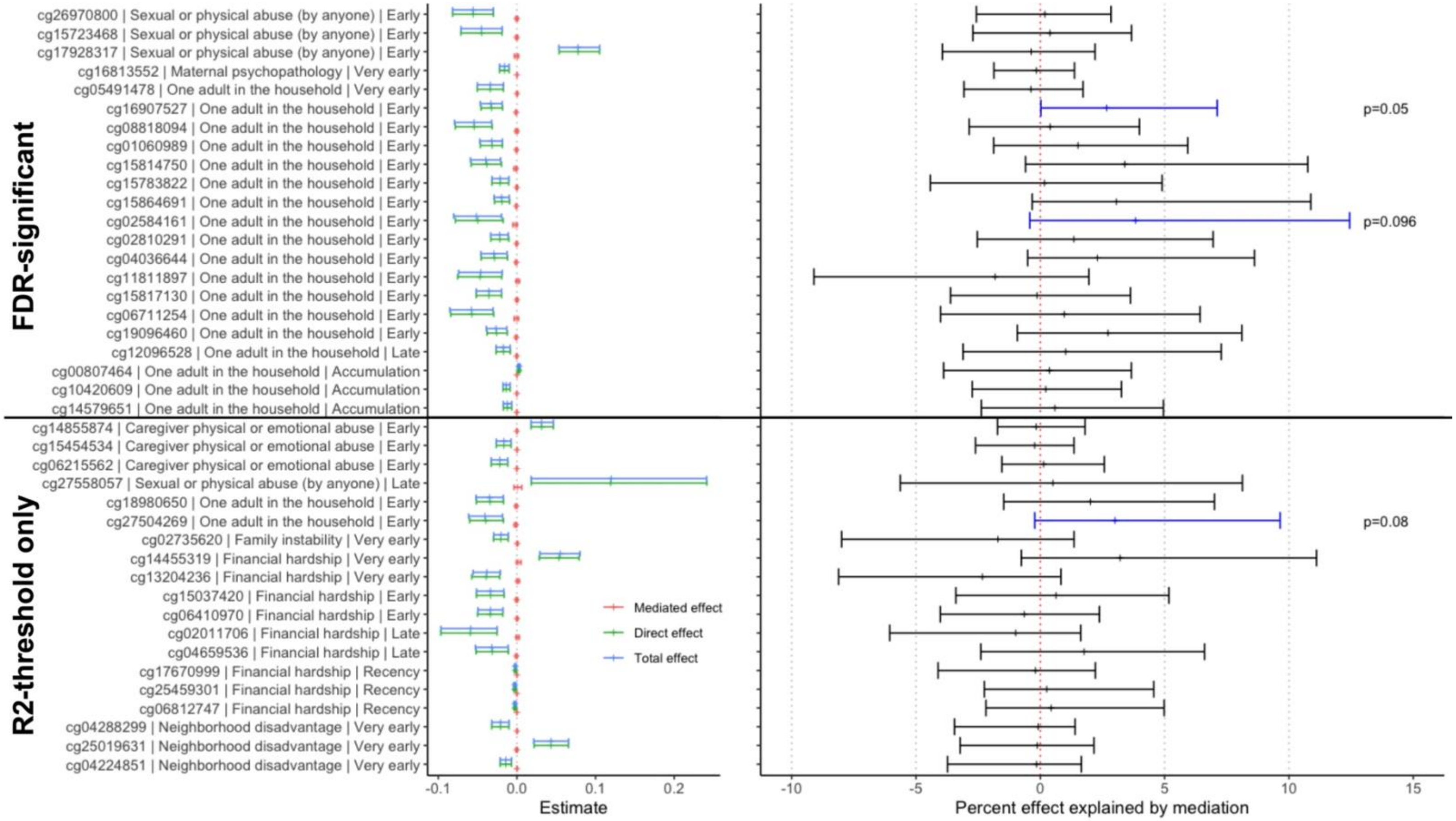

Mediation by body mass index, measured at age 15, was tested for the loci associated with childhood adversity and DNA methylation at age 15. The average causal mediation effect (mediated effect, red; left panel) was near zero for all CpGs, explaining very little of the association between childhood adversity and DNA methylation levels. However, one locus (cg16907527) showed nearly significant mediated effects, explaining 2.67% of the relationship between childhood adversity and DNA methylation (p=0.050). Two other loci showed causal mediation with p<0.1, shown in blue (right panel). No associations were significant after correction for multiple-testing at a false-discovery rate <0.05. Y-axis is noted as “CpG | childhood adversity | SLCMA hypothesis”.

**Figure S15. C-reactive protein levels at age 15 putatively mediated the effects of childhood adversity on age 15 DNA methylation.**

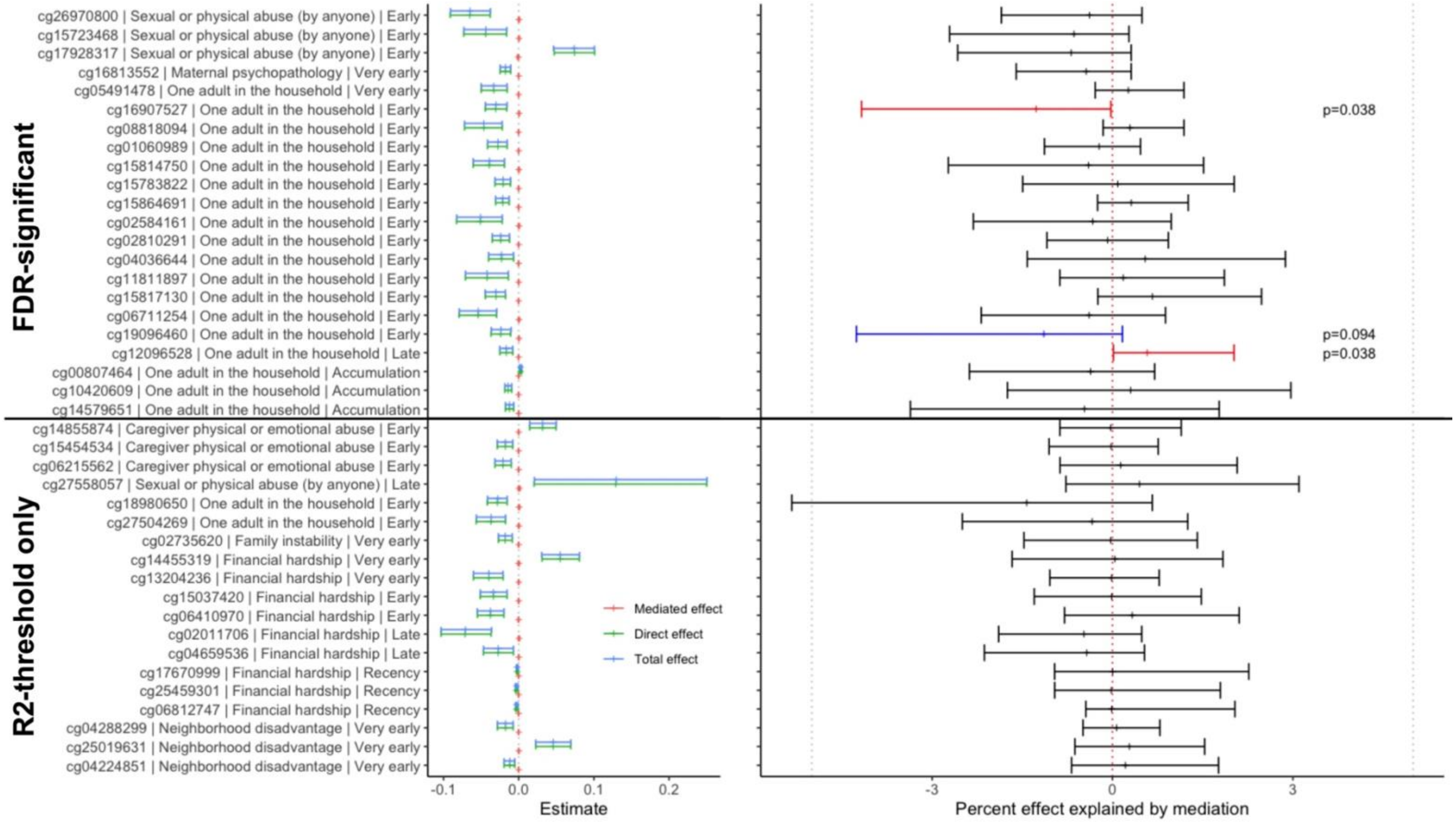

Mediation by the levels of C-reactive protein, measured at age 15, was tested for the loci associated with childhood adversity and DNA methylation at age 15. **A)** The average causal mediation effect (mediated effect, red; left panel) was close to zero for all CpGs, explaining very little of the association between childhood adversity and DNA methylation levels. **B)** Two of the estimated mediated effects were significant (p<0.05, red; cg16907527, *VEGFA*, -1.27% relationship explained; cg12096528, *SLC25A41*, -1.14% relationship explained) and one locus showed a putative causal mediation effect with (p<0.1, blue; cg19096460, *HERC3*). However, none of these passed multiple-test correction. Y-axis is noted as “CpG | childhood adversity | SLCMA hypothesis”.

**Figure S16. The adolescent's daily smoking at age 15 did not mediate the effects of childhood adversity age 15 DNA methylation.**

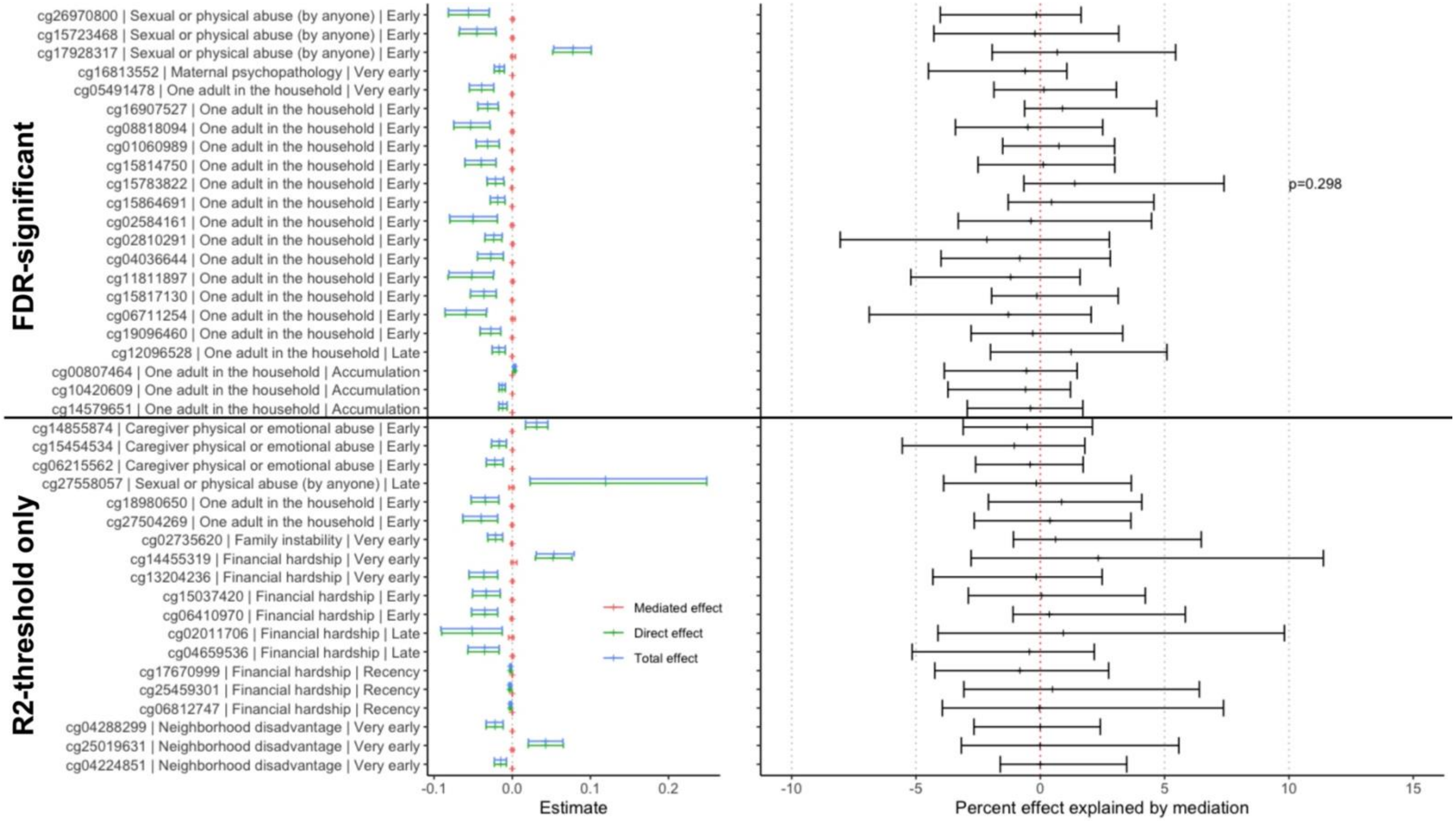

Mediation by smoking behavior at age 15, categorized as the adolescent smoking cigarettes on a daily basis, was tested for the 23 loci significantly associated with childhood adversity and DNA methylation at age 15. The average causal mediation effect (mediated effect, red; left panel) was close to zero for all CpGs, explaining very little of the association between childhood adversity and DNA methylation levels. None of the estimated mediated effects were significant ($p>0.05$). The lowest p-value belonged to cg15783822 ($p=0.298$). Y-axis is noted as "CpG | childhood adversity | SLCMA hypothesis".

**Figure S17. Selection metrics for the number of types of DNAm trajectories across development.**

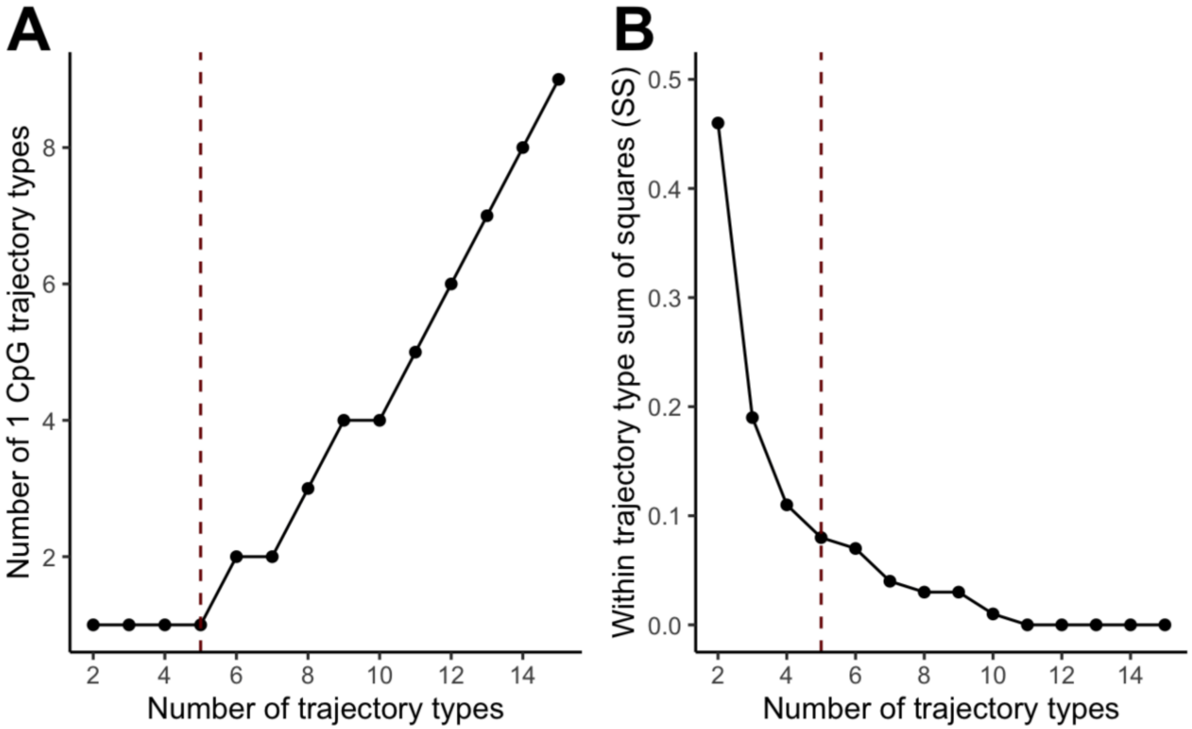

**A)** Number of trajectory types that were composed of a single CpG, with the x-axis showing the total number of different trajectory types. From the 2 to 5 trajectory solutions, only one trajectory type was composed of a single CpG.

**B)** The mean within trajectory type sum of squares shown by number of total trajectories, where lower values reflect closer observations within clusters (i.e., more homogenous clusters). This metric showed an almost complete drop-off by the model with 5 trajectory types, suggesting that the good model fit was achieved.

The red dashed line represents the number of total trajectory types selected for final analyses (5), based on the number of trajectory types with single loci and elbow of the minimal sum of squares plot.

**Figure S18. Hierarchical clustering of CpGs based on a five-trajectory model.**

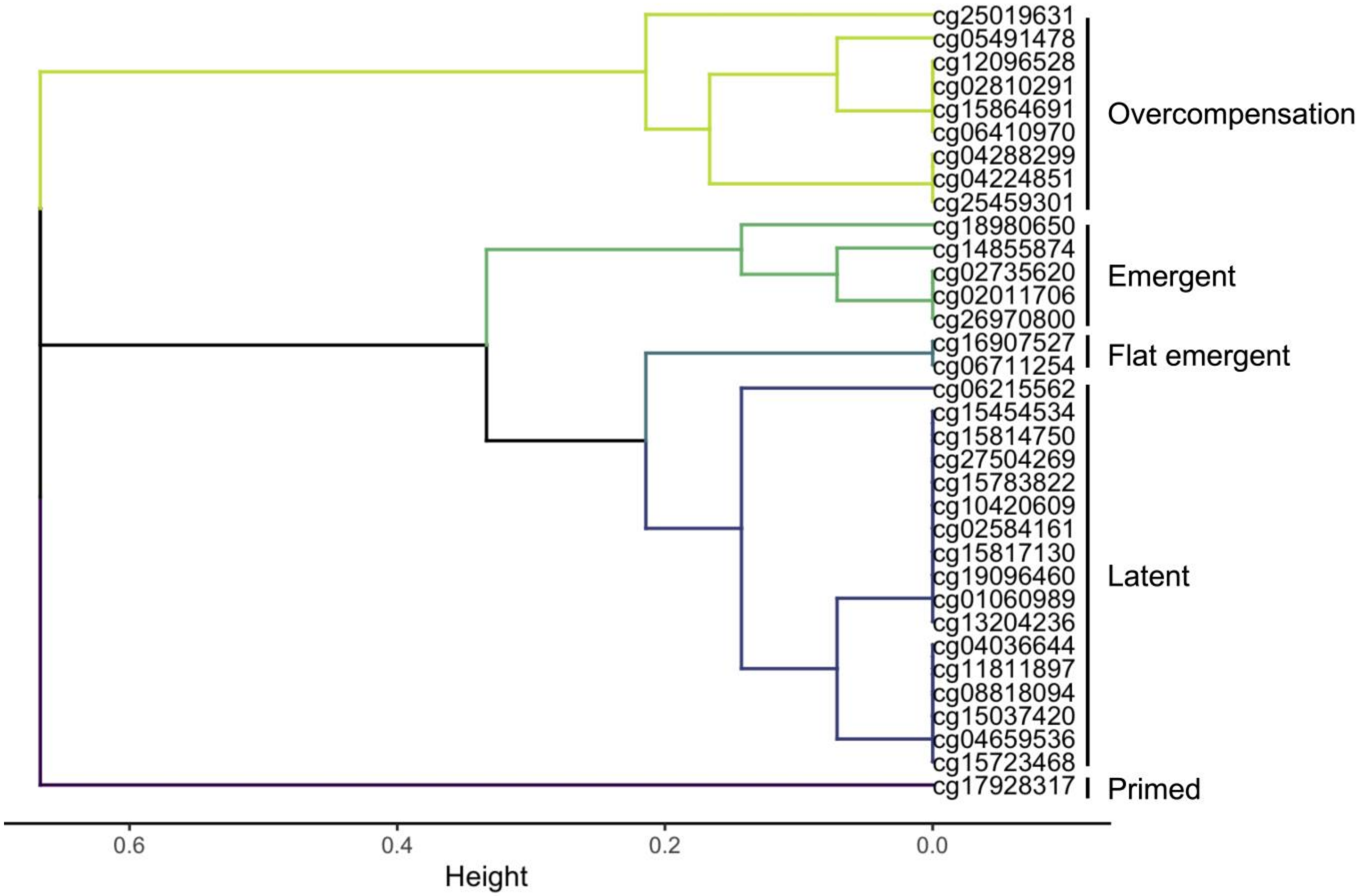

Hierarchical clustering of age 15 loci using Tukey summary statistics for group-by-age interactions revealed five additional types of longitudinal DNAm patterns beyond those that did not show significant group-by-age interactions. These types of trajectories ranged in size from 1 (primed) to 17 CpGs (latent).

**Figure S19. Distinguishing features between the six types of DNA methylation trajectories.**

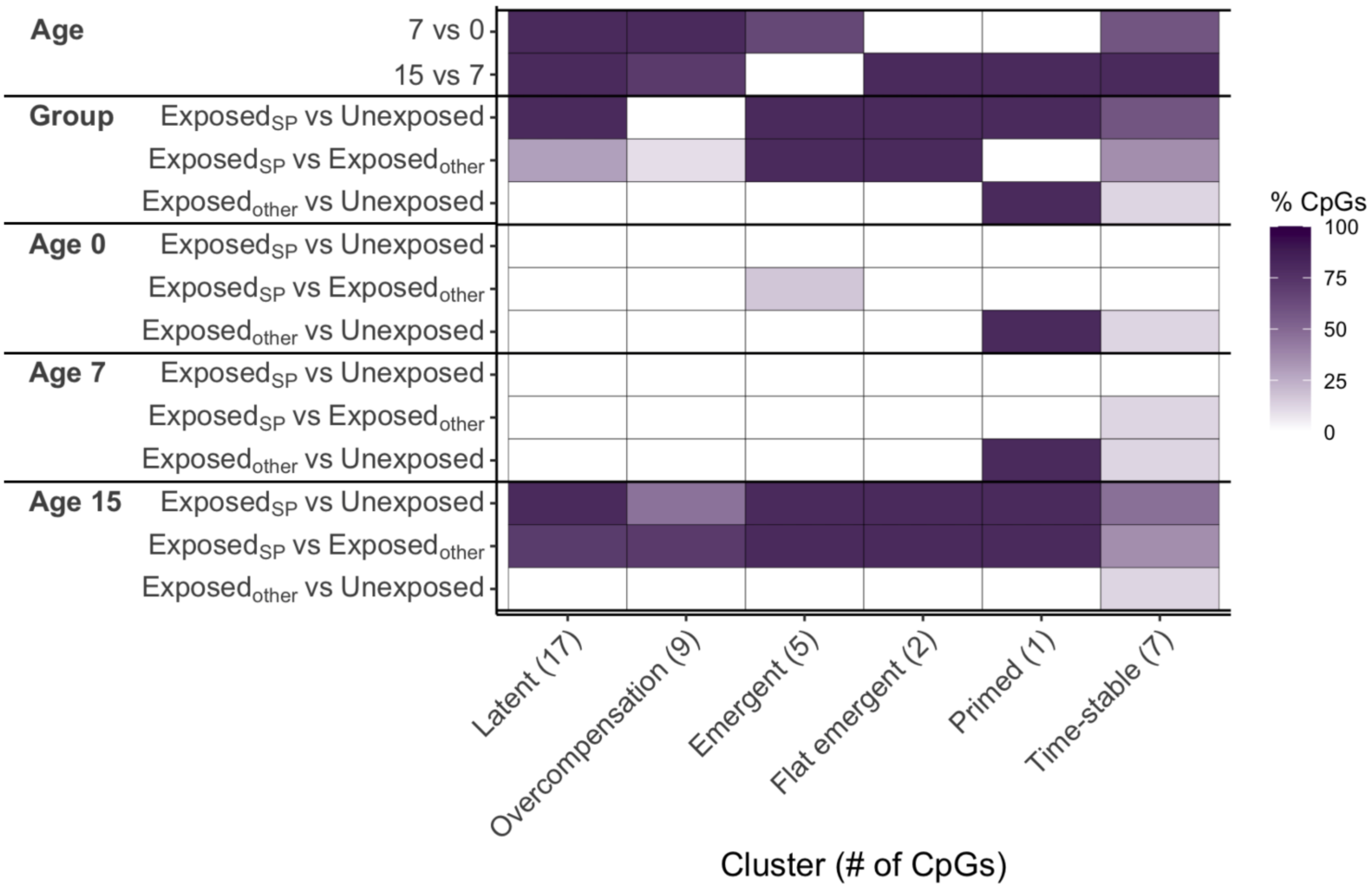

Summary of the significant Tukey summary statistics used to differentiate the six types of DNA methylation trajectories. The fraction of loci with a significant contrast for each type of trajectory is shown (lighter color indicates more loci, or a greater fraction of trajectories). The summary statistics on the y-axis show whether the contrast was significant for: 1) mean differences between ages (age 0, age 7, age 15), 2) mean exposure group differences *across* all ages (exposed during the period identified from the SLCMA [exposed$_{SP}$]; exposed during other period [exposed$_{other}$], or unexposed), and 3) exposure group differences *within* each age.

**Figure S20. Types of DNAm trajectories for the 41 loci identified at age 15.**

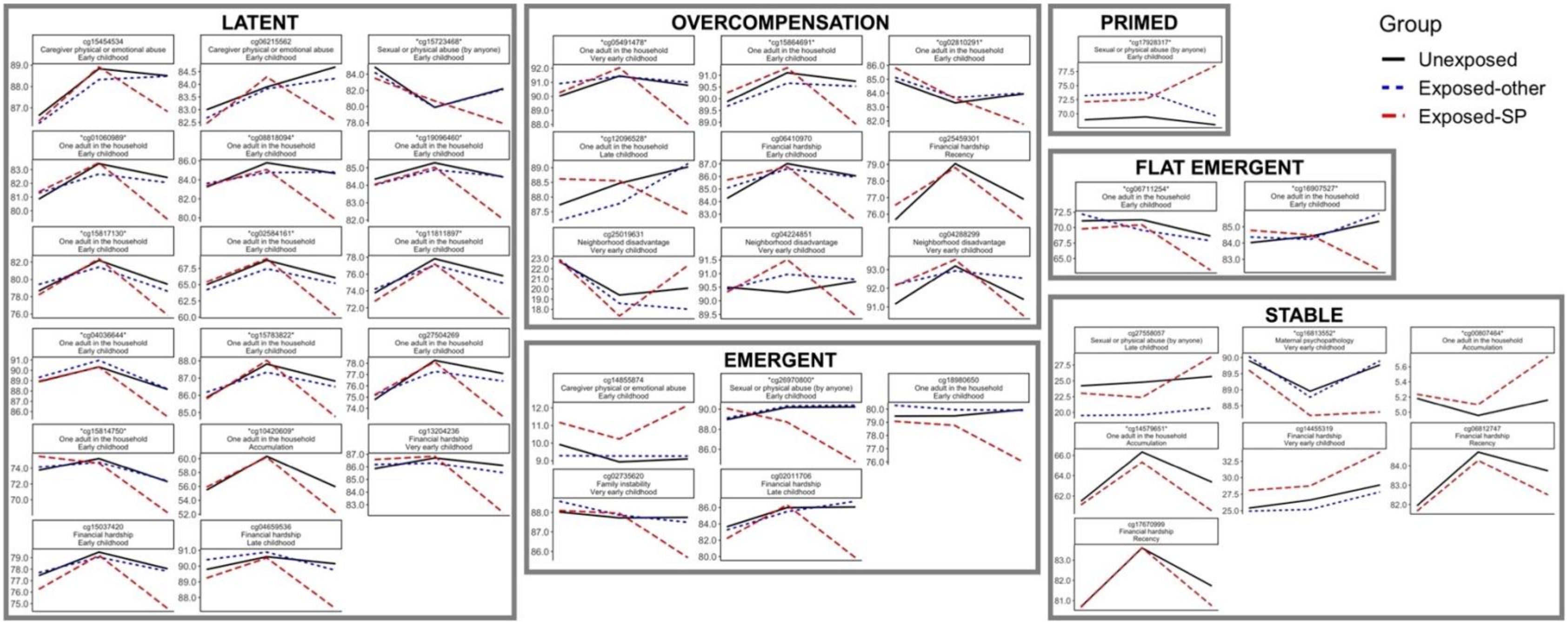

Shown here are the cell-type corrected DNA methylation (DNAm) values on the y-axis and the age at DNAm collection on the x-axis for the 41 loci identified from the SLCMA analyses of age 15 DNAm. Of the 41 loci, seven did not show significant exposure group by age effects (group-by-age effects) and are shown as "Stable". From the 34 loci with significant group by age effects, we identified five distinct types of DNAm trajectories and responses to childhood adversity across development. These DNAm trajectories were identified based on mean exposure group differences *across* ages, mean age differences *across* exposure groups, and exposure group differences at specific ages. Exposure groups were as follows: 1) exposed to adversity *during* the period identified from the SLCMA (exposed-SP; red); 2) exposed to adversity *outside* the period identified from the SCLMA (exposed-other; blue); or 3) unexposed to adversity across development (black). The childhood adversity and hypothesis selected in the SLCMA are shown in the header of each individual plot. Waves of DNAm collection are shown on the x-axis (age 0, 7, and 15 at the inflection points) and percent DNAm is shown on the y-axis.

**Figure S21. Types of trajectories based on the significance threshold of top loci.**

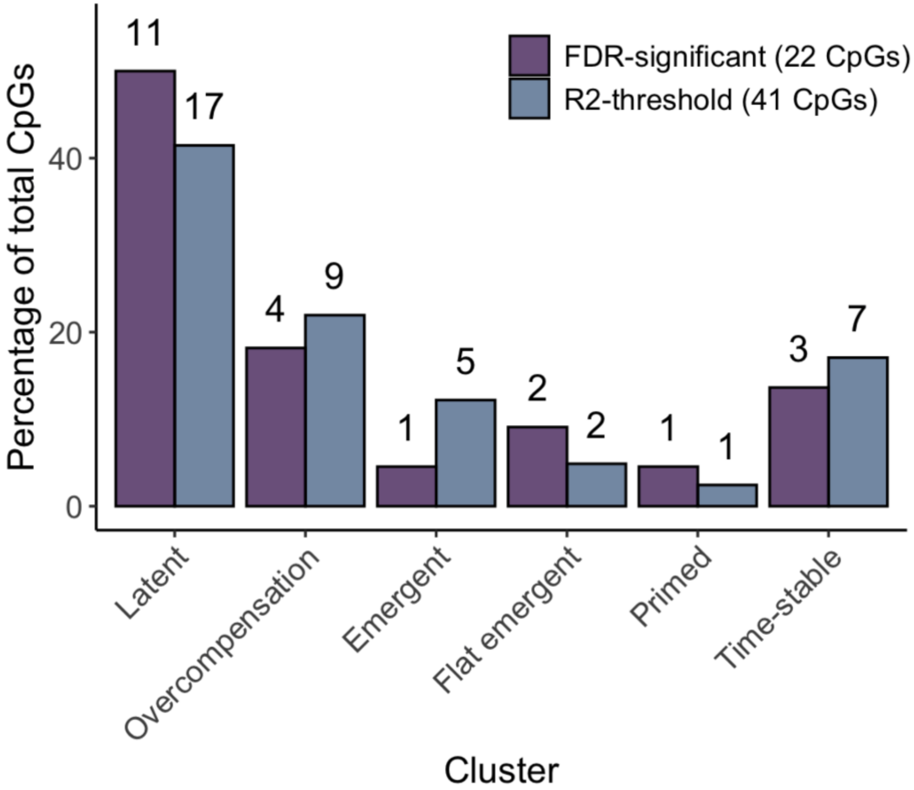

The fraction of CpGs falling within different types of DNA methylation trajectories across development did both vary based on selection thresholds based on and FDR<0.05 or and $R^2 \geq 0.035$ ($\chi^2=1.92$, p =0.86). However, the were generally more CpGs in the latent class and fewer in the emergent class for the FDR-significant loci compared to the $R^2$-threshold loci.

**Figure S22. Enrichment of top adolescent loci within the threat versus deprivation paradigm.**

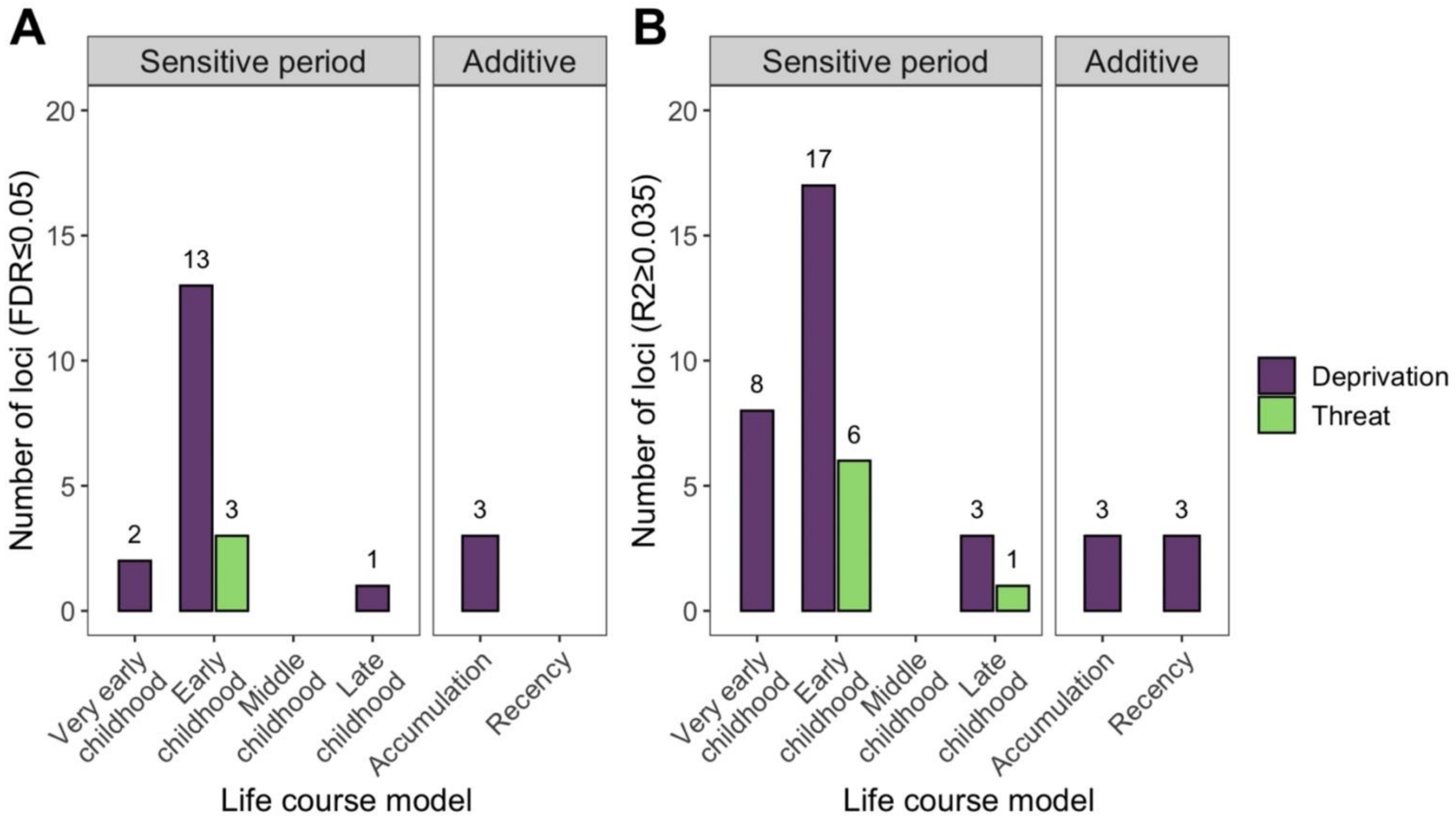

The life course theoretical models were split by sensitive periods (i.e., exposure to adversity during specific childhood periods) or additive models (i.e., accumulation or recency of exposures). Colors represent the two adversity paradigms, threat versus deprivation. **A)** Of the 22 loci identified at a false-discovery rate (FDR) <0.05, most loci were associated with exposure to deprivation during early childhood. **B)** Of the 41 loci identified at an $R2 \geq 0.035$ cutoff and $p<1x10^{-5}$ threshold, most associations were again linked to a deprivation exposure, particular during very early and early childhood. Exposures to threat-type adversities were mainly linked to DNAm when they occurred during early childhood.
